# NOTCH1 fusions in pediatric T-cell lymphoblastic lymphoma: a high-risk subgroup with CCL17 (TARC) levels as diagnostic biomarker

**DOI:** 10.1101/2024.01.31.24301517

**Authors:** Emma Kroeze, Michelle M Kleisman, Lennart A Kester, Marijn A Scheijde-Vermeulen, Edwin Sonneveld, Jessica GC Buijs-Gladdines, Melanie M Hagleitner, Friederike AG Meyer-Wentrup, Margreet A Veening, Auke Beishuizen, Jules PP Meijerink, Jan LC Loeffen, Roland P Kuiper

## Abstract

Twenty percent of children with T-cell lymphoblastic lymphoma (T-LBL) will relapse and have an extremely poor outcome. Currently, we can identify a genetically low-risk subgroup in pediatric T-LBL, yet these high-risk patients who need intensified or alternative treatment options remain undetected. Therefore, there is an urgent need to recognize these high-risk T-LBL patients through identification of molecular characteristics and biomarkers. By using RNA sequencing which was performed in 29/49 T-LBL patients who were diagnosed in the Princess Maxima Center for Pediatric Oncology between 2018-2023, we discovered a previously unknown high-risk biological subgroup of children with T-LBL. This subgroup is characterized by *NOTCH1* gene fusions, found in 21% of our T-LBL cohort (6/29). All patients presented with a large mediastinal mass, pleural/pericardial effusions, and absence of blasts in the bone marrow, blood, and central nervous system. Blood CCL17 (C-C Motif Chemokine Ligand 17, TARC) levels were measured at diagnosis in 26/29 patients, and all six patients with *NOTCH1* gene fusions patients exclusively expressed highly elevated blood CCL17 levels, defining a novel and previously not known clinically relevant biomarker for T-cell lymphoblastic lymphoma. Four out of these six patients relapsed during therapy, a fifth developed a therapy related acute myeloid leukemia during maintenance therapy. These data indicate that T-LBL patients with a *NOTCH1* fusion have a high risk of relapse which can be easily identified using a blood CCL17 screening at diagnosis. Further molecular characterization through *NOTCH1* gene fusion analysis offers these patients the opportunity for treatment intensification or new treatment strategies.

## Introduction

T-cell lymphoblastic lymphoma (T-LBL) is a common pediatric malignancy accounting for approximately 20% of the non-Hodgkin lymphomas during childhood^1^. Survival rates of T-LBL are ∼80%, but outcome after relapse is dismal, with salvage rates reaching only ∼15%^2,3^. Considering the extremely poor prognosis after relapse and absence of clinically relevant high-risk genetics, there is an urgent need for the identification of molecular risk factors and new prognostic biomarkers in T-LBL, as well as identification of new therapeutic strategies.

Pediatric T-LBL is typically characterized by infiltration of blasts in the mediastinum (thymus) and lymph nodes. Approximately half of the cases present with pleural effusion at diagnosis and, by definition, T-LBL patients have less than 25% blasts in the bone marrow (BM)^4,5^. Based on morphology and immunophenotype, T-LBL is indistinguishable from its leukemic counterpart, T-cell acute lymphoblastic leukemia (T-ALL). T-ALL presents as leukemic disease with ≥25% blasts in the BM and presence of blasts in the peripheral blood (PB), but usually with mild mediastinal enlargement. Even though the clinical presentation of T-LBL and T-ALL largely differs, there has been no evidence so far that there are major differences in molecular genetics of T-LBL and T-ALL^6,7^. Therefore, T-LBL and T-ALL are regarded as different manifestations of the same disease^8^. Pediatric T-ALL has been extensively studied and great advances in treatment protocols have been made, including minimal residual disease (MRD) measurements as a useable stratification strategy. In contrast, molecular genetics underlying T-LBL are still poorly understood and T-LBL patients are currently mainly treated according to ALL-based protocols. Thus far, MRD assessment has not proven to be usable in T-LBL and diagnostic biomarkers to identify high-risk patients are lacking.

Diagnostic biomarkers have been successfully introduced in other types of lymphomas. For example, CCL17, also known as blood *thymus and activation-regulated chemokine* (TARC) has proven to be a useful diagnostic biomarker in children with classical Hodgkin lymphoma^9,10^. It has never been studied whether CCL17 can serve as a biomarker in non-Hodgkin lymphoma and T-LBL in particular. CCL17 is constitutively produced in the thymus, acting as a powerful T-cell chemoattractant. In classical Hodgkin lymphoma patients, CCL17 production can be induced by NOTCH1 and CCL17 is highly expressed by the Reed-Sternberg cells, thereby creating a specific supporting tumor microenvironment that recruits T-cells^10^.

Considering the importance of NOTCH1 and strong preference of malignant T-cells in T-LBL for the thymus, we hypothesize that CCL17 is of importance in the pathophysiology of T-LBL and creating a thymic microenvironment that favors the T-LBL cells.

Recent studies show that both T-LBL and T-ALL patients with *NOTCH1* and/or *FBXW7* DNA mutations have a better outcome compared to *NOTCH1* and *FBXW7* wildtype patients^11,12^. The pathogenic molecular mechanisms of T-LBL patients without *NOTCH1* and/or *FBXW7* mutations remain largely unknown, and this group probably contains both high-risk and low-risk patients. Considering the extremely poor prognosis after relapse, it is essential to identify these high-risk patients. Additionally, there is an urgent need for the identification of new prognostic biomarkers in T-LBL. In this study we present a novel entity of pediatric T-LBL patients characterized by previously unknown *NOTCH1* gene fusions, high risk of relapse, and highly elevated blood CCL17 (TARC) levels

## Methods

### Patients

We included a complete cohort of all pediatric T-LBL patients (n=49) that were diagnosed in the Princess Máxima Center for Pediatric Oncology between 2018-2024. RNA sequencing data at diagnosis were available for 29/49 patients and at relapse for two additional patients. Clinical information and hematological values at diagnosis were retrieved from patient files. All patients were treated according to the EURO-LB02 protocol^3^ or its successor, the LBL2018 protocol (NCT04043494). *NOTCH1*/*FBXW7* mutational status was determined for most patients and retrieved from patient files. Pediatric T-ALL patients (n=39) diagnosed between 2019-2022 at the Princess Máxima Center for Pediatric Oncology were included as reference cohort. All sample IDs are completely anonymized. Written informed consent was obtained from all patients and/or their legal guardians included in this study. The study was performed in accordance with the Declaration of Helsinki. The Medical Research Ethical Committee Utrecht declared that the Medical Research Involving Human Subjects Act (WMO) does not apply, and has approved the study (19-140/C).

### RNA sequencing analysis

RNA sequencing data was obtained from the in-house diagnostics department in the Princess Máxima Center. The source of biological material is given in **Supplementary Table 1**. Pre-processing of the data was done with standardized and in-house pipelines and guidelines^13^. Fusion detection was performed using STAR fusion^14^. In addition, we analyzed the entire T-LBL and T-ALL cohort for exon-specific *NOTCH1* coverage as an indication for fusions, by using DepthOfCoverage of GATK v3.8.0. Whole genome sequencing data (WGS) were also obtained from the in-house diagnostics department and used for validation of genomic breakpoints.

### Gene expression analyses

For expression analyses, samples with less than 30 million unique reads were excluded due to lower quality (TLBL042, TLBL046 and TLBL058). For TLBL042, expression data from the relapse were used (TLBL042_R). Gene expression alterations were assessed with log2 transformed gene length normalized read counts (transcript per million mapped reads, TPM) using R v4.4.0. Gene expression variance was determined by calculating standard deviations and the 200 most variable genes were taken in unsupervised clustering using Euclidean distance as a measure of similarity. The R package pheatmap v1.0.12 was implemented for visualizations.

Differential expression analyses were performed with DESeq2 v1.36.0^15^ between NOTCH1-rearranged- and WT cases and between NOTCH1-mutated- and WT cases. Differentially expressed genes were identified after adjustment for false discovery rate (FDR-adjusted p-value ≤ 0.05). Visualisations were generated using R packages EnhancedVolcano v1.12.0 and pheatmap v1.0.12. Subsequently, gene set enrichment analysis (GSEA) was conducted for biological interpretations with clusterProfiler v4.12.0^16,17^. Genes were ranked according to direction-signed log10-transformed p-values, as determined by DESeq2 v1.36.0, and annotations were provided by implementing the R package AnnotationHub v3.2.2. The ranked gene list was used for Kyoto Encyclopedia of Genes and Genomes (KEGG) pathway enrichment analysis. Additionally, z-scores of log2-transformed TPM values were calculated within the complete T-LBL cohort for genes associated with the NOTCH signaling pathway. ClusterProfiler v4.12.0 and ComplexHeatmap v2.10.0 were implemented for GSEA-result visualizations.

### CCL17 (TARC) measurements and immunohistochemistry

CCL17 measurements at diagnosis were performed in serum or plasma of 26 out of the 29 patients for whom RNAseq was available. Measurements were performed in triplo by standard enzyme-linked immunosorbent assay (ELISA) using the DuoSet ELISA kit (cat. no. DY364; R&D Systems, Inc.). Immunohistochemistry staining for CCL17 was performed on the BOND-III fully automated staining system (Leica, IL, USA) using CCL17 rabbit polyclonal antibody (ProteinTech group, Chicago IL, USA). Significance of *CCL17* expression between *NOTCH1*-rearranged samples and the rest of the T-LBL RNAseq cohort was determined using DESeq2 v1.36.0.

## Results

### Patient characteristics

The pediatric T-LBL patients included in this study (n=49) represented an unselected complete cohort diagnosed in the Princess Máxima Center for Pediatric Oncology between 2018-2023. T-LBL diagnoses were based on histopathological classification according to the revised world health organization for hematological malignancies and/or flow cytometry according to the criteria of the European Group for Immunophenotyping of Leukemias^8,18^. Six out of 49 patients (12%) had a relapse. Informed consent was obtained for 45/49 T-LBL patients and these were therefore used for further analysis. The median age at diagnosis for these 45 patients was ten years, and our cohort contained more males than females (59%), in line with previously described large T-LBL cohorts^3^. Eighty-eight percent of the patients presented with a large mediastinal mass, which was in 56% of these cases accompanied by pleural effusion. RNAseq was performed for 29/43 patients and their clinical characteristics are described in **Supplementary Table 1**.

### Detection of NOTCH1 rearrangements

Transcriptome sequencing was performed at diagnosis from 2019 onwards for all samples with sufficient good quality material available (n=29). This technique allows for the identification and quantification of fusion transcripts and gene expression levels. We identified 12 fusions transcripts in total (12/29, 41%), of which six were *NOTCH1* gene fusions. Fusion partner of *NOTCH1* werethe microRNA gene *miR142HG* on chromosome 17q22 (TLBL042 and TLBL058), *IKZF2* on chromosome 2q34 (TLBL050) and *TRBJ* on chromosome 7q34 (TLBL033, TLBL049, TLBL050) (**Supplementary Table 2**). The *TRBJ::NOTCH1* fusions were missed by fusion detection algorithms, but were detected because of pronounced expression differences between exons in the 5’ and 3’ part of the gene (see **Methods**). The fusion transcripts with *miR142HG* and *IKZF2* demonstrated correct splicing to exon 27 or 28 of *NOTCH1* and genomic breakpoints were identified using available whole genome sequencing (WGS) data (**Supplementary Table 2**). *TRBJ::NOTCH1* fusion genes were previously shown to express a truncated, membrane-bound form of NOTCH1^19^ (**Figure 1A**). None of the six samples with a *NOTCH1* gene fusion exhibited mutations in *NOTCH1*/*FBXW7*, demonstrating the mutual exclusivity of *NOTCH1* mutations and *NOTCH1*-rearrangements. Furthermore, apart from homozygous loss of the *CDKN2A/2B* locus in two cases (TLBL042 and TLBL050), no other driver events were found in these *NOTCH1*-rearranged T-LBLs. *NOTCH1* gene fusions are almost never found in T-ALL, but considering the difficulties in detecting TR-rearranged fusions with conventional pipelines, we also reanalyzed exon-specific *NOTCH1* expression in 39 T-ALL samples that were diagnosed in the Princess Máxima Center for Pediatric Oncology. In line with previous studies^20,21^, none of the T-ALL samples carried gene fusions involving *NOTCH1*, thus suggesting that the frequent occurrence of these fusions represents an important molecular genetic discriminator between T-LBL and T-ALL.

**Figure 1:**
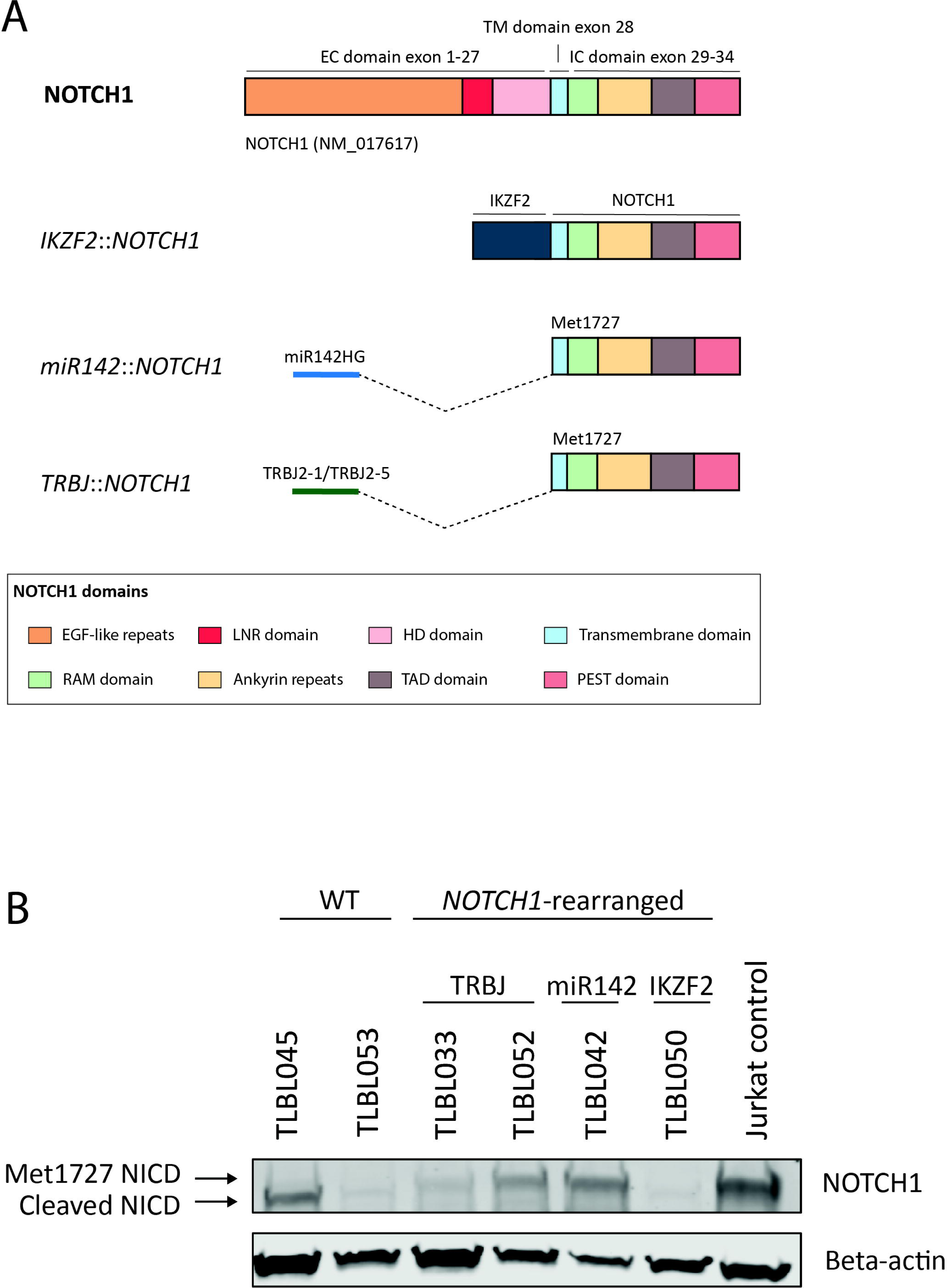
NOTCH1-rearrangements in T-LBL. A) Schematic representation of three different *NOTCH1* fusions with different fusion partners. The in-frame *IKZF2*::*NOTCH1* fusion generates a chimeric protein in which the N-terminal DNA binding domain of IKZF2 is fused to the C-terminal intracellular domains of NOTCH1. Fusions transcripts with *miR142HG* and *TRBJ* use an alternative translation start site in exon 28 (Met1727). B) Western blot analysis using Val1744 antibody (Cell Signaling Inc.) shows that *miR142HG::NOTCH1* and the *TRBJ::NOTCH1* fusions lead to a larger NICD protein, likely representing uncleaved NICD with translation initiation at Met1727. Simultaneous Beta-actin staining was performed for loading comparisons.

### All NOTCH1 gene fusions result in expression of intracellular NOTCH1

NOTCH1 is a transmembrane protein that is activated through ligand-receptor interaction, which induces a conformational change that results in dissociation of the heterodimerization (HD) subunits. This is followed by exposure of a cleavage site in the C-terminal part of the HD domain, resulting in the release of the intracellular domain of NOTCH1 (NICD). NICD subsequently translocates to the nucleus where it acts as a transcriptional regulator^22^.

To determine whether a truncated C-terminal version of NOTCH1 was indeed expressed in the T-LBL samples with a *NOTCH1*-rearrangement, we performed Western blotting using protein lysates of four *NOTCH1*-rearranged cases and two *NOTCH1* wildtype (*NOTCH1*-WT) cases. All *NOTCH1*-rearrangements lead to expression of NICD. The *IKZF2::NOTCH1*-positive T-LBL (TLBL050) showed expression of an NICD protein co-migrating with the ∼130-kDa wildtype NICD, suggesting cleavage of the chimeric protein at the 1-secretase cleavage side (Val1744). In contrast, the *TRBJ::NOTCH1*-positive (TLBL033 and TLBL052) and *miR142HG::NOTCH1*-positive (TLBL042) cases expressed a slightly larger NICD protein, which is in line with translation initiation at methionine residue 1727 (Met1727) encoded in exon 28 of *NOTCH1*, as previously reported^19,23^, and lack of 1-secretase cleavage (**Figure 1B**).

To explore the downstream consequences of T-LBL cases with a *NOTCH1* fusion in our T-LBL cohort, we performed unsupervised clustering analysis with the 200 most variable genes in the dataset (**Supplementary Table 3**), which showed that *NOTCH1*-rearranged samples mostly clustered together (**Supplementary Figure 1**). These data indicate that *NOTCH1*-rearranged samples display similar expression profiles among each other. Next, we performed differential expression analysis between *NOTCH1*-rearranged and *NOTCH1*-WT T-LBLs, to determine expression differences. A total of 1,288 genes were found to be significantly differentially expressed, compared to only 101 genes in a comparison between *NOTCH1*-mutated and *NOTCH1*-WT T-LBLs (**Figure 2A,B** and **Supplementary Tables 4 and 5**). These data indicate that *NOTCH1*-mutated and *NOTCH1*-WT cases exhibit comparable expression profiles, whereas *NOTCH1*-rearranged cases differ substantially from the rest of the cohort. Subsequently, we selected the 200 most significantly differentially expressed genes between *NOTCH1*-rearranged and *NOTCH1*-WT T-LBLs for supervised clustering. This analysis revealed that *NOTCH1*-rearranged samples formed a separate cluster, whereas the NOTCH1 mutated and *NOTCH1*-WT cases are mixed in a second cluster (**Figure 2C**), confirming the unique characteristics of the *NOTCH1* fusion samples.

**Figure 2:**
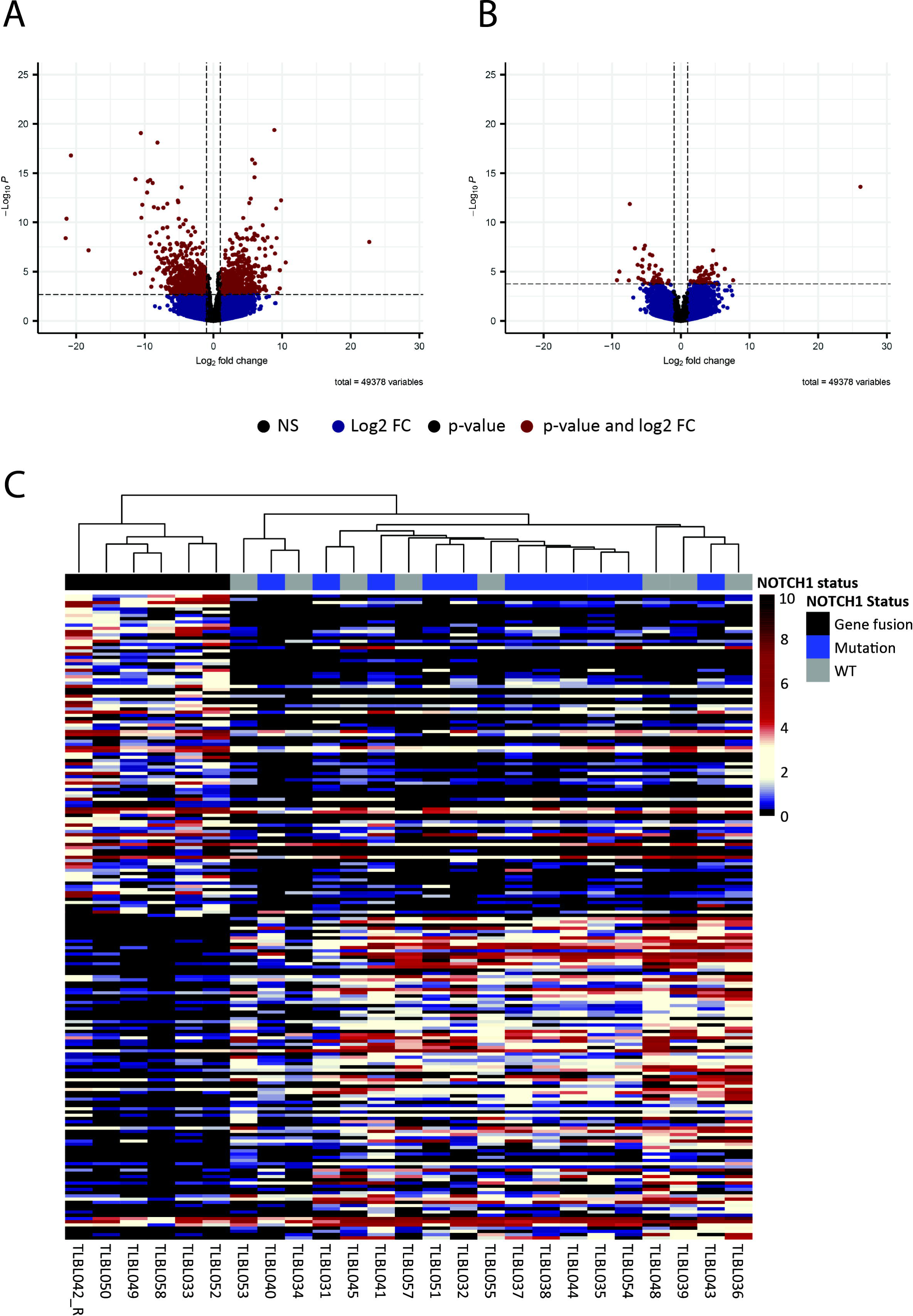
Gene expression differences in NOTCH1-rearranged, mutated and wildtype T-LBL. A+B) Volcano plots showing differentially expressed genes between *NOTCH1*-rearranged and *NOTCH1*-WT samples (n=1,288; panel A) and between *NOTCH1*-mutated and *NOTCH1* WT samples (n=101; panel B) C) Expression analysis of the 200 most significantly up- and downregulated genes (from a total of 1,288 genes) in *NOTCH1*-rearranged compared to *NOTCH1*-WT samples, revealed that *NOTCH1*-rearranged samples cluster separately from NOTCH1 WT and NOTCH1-mutated samples using Euclidean distance as a measure of similarity. The relapse sample of TLBL042 was used because of better quality. Range of 0-10 showing the log2-transformed TPM values. Significance was determined using false discovery rate (FDR)-adjusted p-values.

To explore downstream characteristics of *NOTCH1*-rearranged cases, we performed GSEA, to determine enriched KEGG pathways (**Supplementary Figure 2A**). Among others, the NOTCH signaling pathway was significantly activated in *NOTCH1*-rearranged samples compared to *NOTCH1*-WT samples, with an enrichment score of 0.695 (adj. p-value=0.0003 (**Figure 3A,B**). Enrichment of KEGG pathways was to a lesser extent observed in *NOTCH1*-mutated cases compared to *NOTCH1*-WT samples, including no significant enrichment of the NOTCH signaling pathway (**Supplementary Figure 2B**). These findings suggest that the downstream characteristics and mechanisms of action of the *NOTCH1*-rearranged samples are different from both wildtype and mutant T-LBL samples.

**Figure 3:**
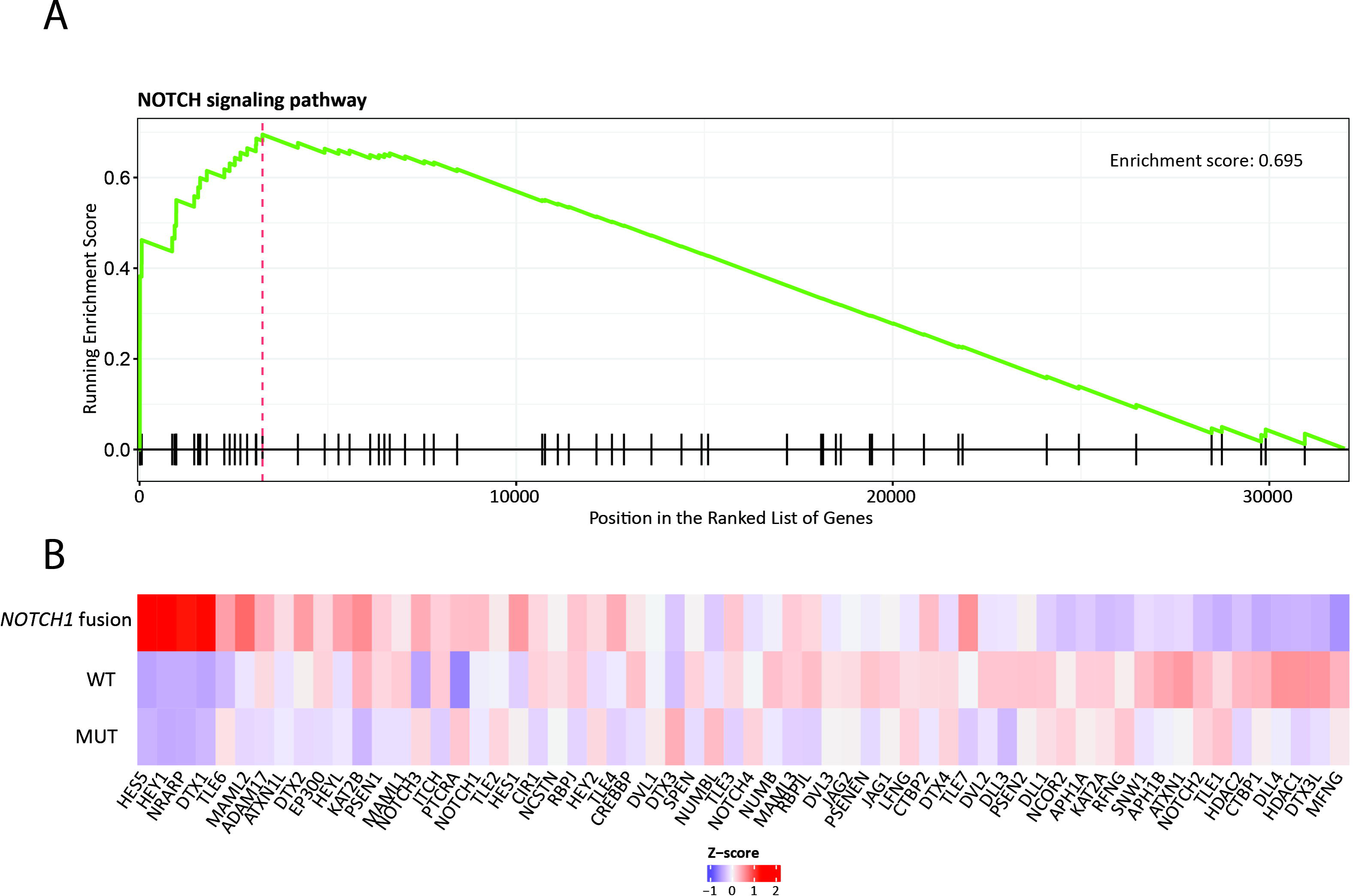
GSEA results of the NOTCH signaling pathway. A) Enrichment plot for the NOTCH signaling pathway in the *NOTCH1*-rearranged versus WT samples, showing the profile of the running enrichment score and positions of genes associated with this pathway on the rank-ordered gene list. B) Z-scores of log2-transformed TPM values of genes associated with the NOTCH signaling pathway within the complete T-LBL cohort. Mean z-scores are depicted for *NOTCH1*-rearranged, *NOTCH1*-mutated and WT samples. Genes are ranked based on their position in the rank-ordered gene list used for GSEA between *NOTCH1*-rearranged and WT samples.

### Clinical presentation of patients with NOTCH1 gene fusions

All six patients with *NOTCH1* gene fusions presented with a massively enlarged mediastinum, combined with pleural/pericardial effusion. Moreover, all *NOTCH1* gene fusion-positive patients were bone marrow negative, peripheral blood negative and cerebral spinal fluid negative. These patients therefore had a uniform clinical presentation of disease, although not differentiating between this group and the rest of the T-LBL cohort. Flowcytometry performed at diagnosis revealed positivity for cytoplasmatic CD3 (cyCD3) in all cases, as well as positivity for other T-cell markers. Precursor-marker Terminal Deoxynucleotidyl Transferase (TdT) was expressed in 50% of the *NOTCH1*-rearranged cases, which was lower than expected (∼90%)^24^ (**Supplementary Table 6**). Next, we analyzed blood CCL17 levels, which were highly elevated in all patients with a *NOTCH1* gene fusion (range from 2345 to >10000 pg/ml), compared to 31-638 pg/ml in 16 patients who did not have a *NOTCH1* gene fusion (p<0.0001, t-test) (**Figure 4A**). Follow-up CCL17 levels during first remission were available for 3/6 patients and revealed normalized values (range 57-152 pg/ml) (**Figure 4B**). Three of the patients with a clinical relapse (TLBL042, TLBL050 and TLBL058) also showed substantially elevated CCL17 levels at relapse (TLBL042:4613 pg/ml, TLBL050:8654 pg/ml, TLBL058:1662 pg/ml), which could be an indication that CCL17 levels in blood might also increase upon progression of relapse. One patient, whose relapse was discovered with routine imaging, presented with relatively little tumor load and low LDH levels, and did not have increased CCL17 levels (TLBL033) at time of establishing the relapse. CCL17 levels decreased again in second remission (range 69-1331 pg/ml) (**Figure 4C**). Although immunohistochemistry did not reveal positivity of the T-LBL cells for CCL17 (**Figure 4D**), based on gene expression, *CCL17* was significantly upregulated in the *NOTCH1*-rearranged cases compared to the rest of the T-LBL cohort (FDR-adjusted p-value=0.019). Together, our data strongly indicate that CCL17 can serve as a high-risk biomarker at diagnosis.

**Figure 4:**
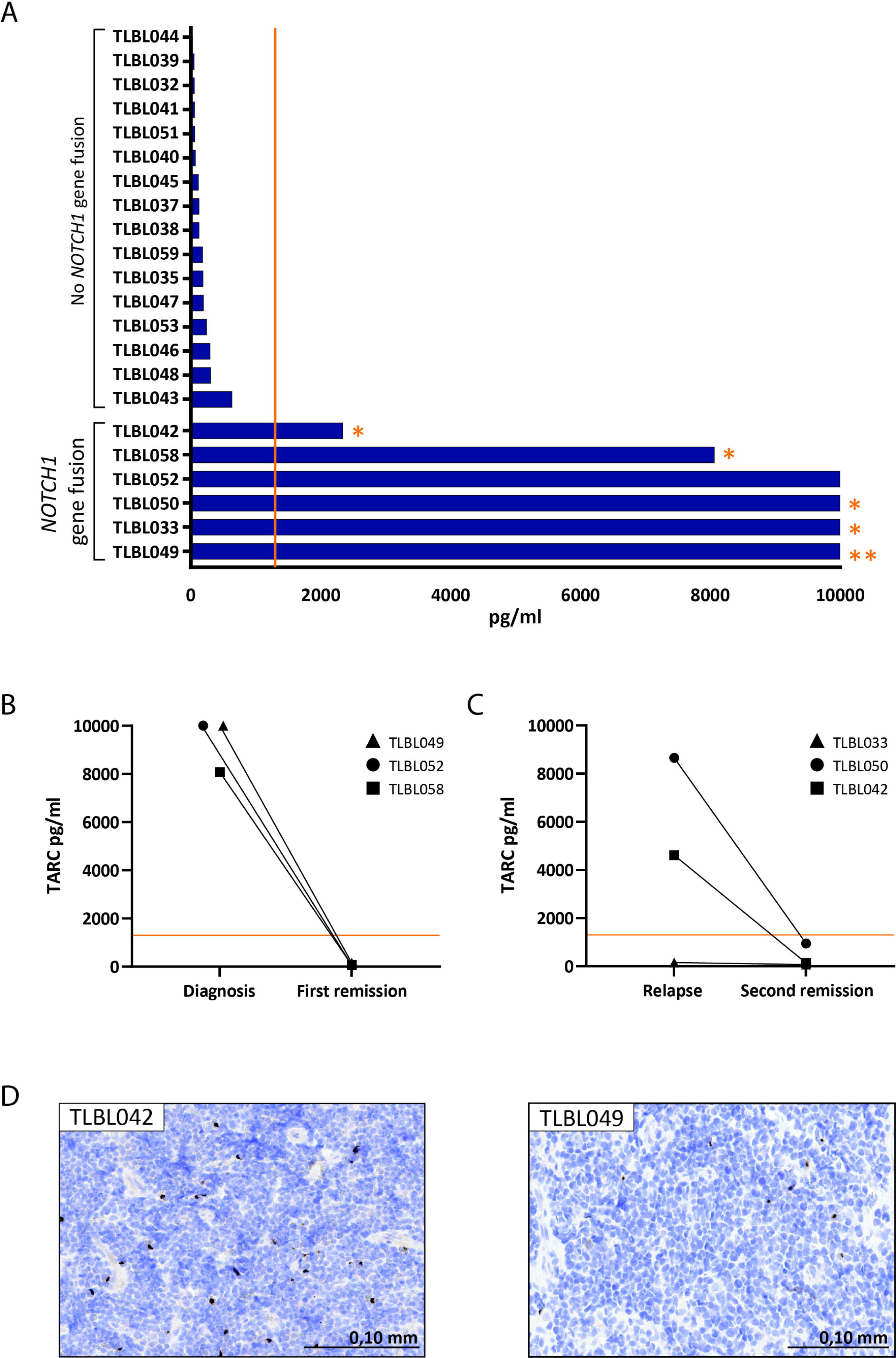
CCL17 (TARC) in *NOTCH1*-rearranged patients. A) CCL17 levels in pg/ml per patient, showing highly elevated CCL17 in blood of *NOTCH1*-rearranged T-LBL patients but none of the other patients. 10000 pg/ml is the maximum measurable CCL17 level with used assay. Orange line in figure A, B and C represents maximum normal CCL17 level (1300 pg/ml) based on what has been described in Hodgkin lymphoma^9^. Patients that had a relapse are indicated with an asterisk. Patient TLBL049 developed a therapy-related AML (double asterisk). B) For three *NOTCH1*-rearranged patients, blood CCL17 levels could be determined for a time point of remission after diagnosis, revealing normalized CCL17 levels in all three cases. C) For four *NOTCH1*-rearranged patients blood CCL17 levels were determined at time point of relapse and remission after relapse (second remission), revealing increased levels in three relapses that again normalized in second remission. D) Staining for TARC using anti-CCL17 antibody for four *NOTCH1*-rearranged patients showing that T-LBL cells do not express high levels of CCL17 based on immunohistochemistry.

### NOTCH1 gene fusions as poor prognostic marker in T-LBL

Finally, we aimed to determine the prognostic relevance of *NOTCH1* gene fusions in T-LBL. We found that five out of six *NOTCH1* fusion-positive patients had an event. Four patients had a relapse during therapy (TLBL033, TLBL042, TLBL050, TLBL058), one of them is still under treatment, the other three could not be rescued. Additionally, one patient had a therapy-related acute myeloid leukemia (t-AML) during maintenance therapy of T-LBL (TLBL049), leaving just one *NOTCH1*-fusion positive patient without an event. The t-AML carried the typical *KMT2A*::*MLLT3* fusion. This patient was rescued with AML induction chemotherapy followed by allogeneic stem cell transplantation. The sixth *NOTCH1*-fusion positive patient did not have an event, yet this patient is still under maintenance treatment. In the rest of the cohort, one event occurred, which was death due to pancreatitis complicated by a septic shock (**Figure 5**). Our data therefore shows a significant difference in cumulative incidence of events between the *NOTCH1*-fusion group and the rest of the cohort (p<0.001)

**Figure 5:**
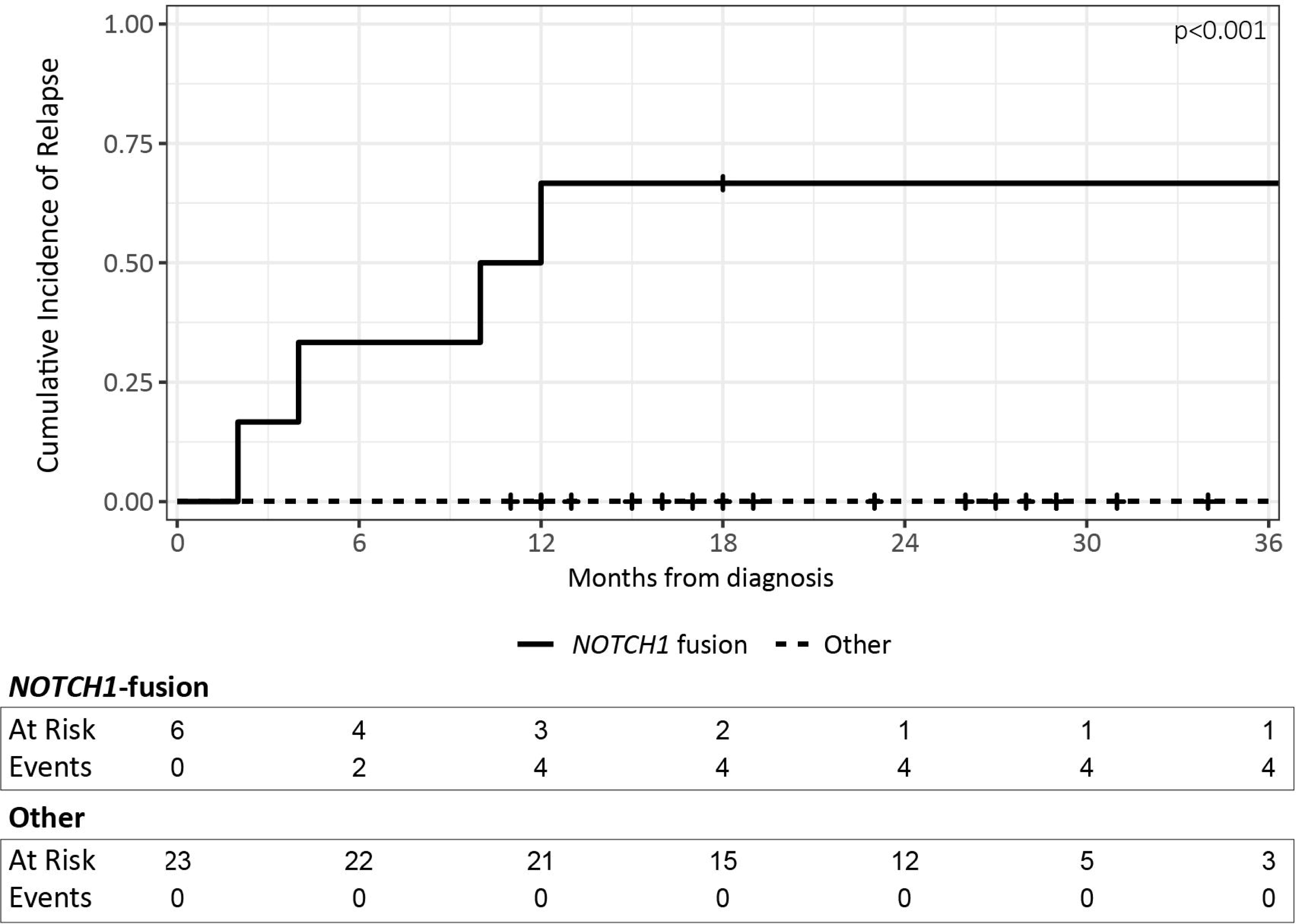
Cumulative incidence plot of events reveals a significant higher cumulative incidence of relapse in *NOTCH1*-rearranged cases compared to the rest of the T-LBL cohort (3 years) (p<0.001). The p-value is estimated using Gray’s test. Four relapses occurred in the *NOTCH1*-rearranged cohort. No relapses occurred in the cohort. In both cohorts, one other event occurred, which was a therapy related-AML in the *NOTCH1*-rearranged cohort and death due to pancreatitis complicated by a septic shock in the other cohort.

The unselected 5-year T-LBL cohort (n=49) contained six patients who relapsed, of whom four had a *NOTCH1* gene fusion. For the other two patients, only RNAseq data at relapse was available, revealing a *DDX3X*::*MLLT10* fusion and a *JAKMIP2*::*PDGFRB* fusion at relapse, respectively, established high-risk ALL aberrations^20,25^. Thus, *NOTCH1*-rearranged T-LBL caused the majority of relapses in our cohort, suggesting that they cause an aggressive T-LBL phenotype, with a significantly higher cumulative incidence of relapse (p<0.001, Gray’s test) compared to the *NOTCH1*-fusion negative patients.

## Discussion

To date, mainly clinically applicable low-risk genetics have been described for T-LBL. This implicates that molecular genetic high-risk patients who need intensified or alternative treatment strategies are undetected. Moreover, patients with unknown low-risk molecular genetic profiles could also be overtreated with current treatment strategies.

We discovered a biological high-risk subgroup of T-LBL, characterized by *NOTCH1* gene fusions. This subgroup represents 21% (6/29 patients) of our T-LBL cohort. All patients had a similar although not unique presentation of disease predominantly consisting of a large mediastinal mass, pleural/pericardial effusion and highly elevated CCL17 (TARC) levels in blood. Moreover, four out of six patients with a *NOTCH1* gene fusion had a relapse and did not survive, indicating that *NOTCH1* fusions lead to an aggressive T-LBL phenotype. Fifty percent of the *NOTCH1*-rearranged cases exhibited expression of TdT, which is lower than the expected 90%. It has been described before that the TdT-negative subset often represent diagnostically challenging cases with phenotypic features that are consistent with a late cortical subtype, coinciding with what we found in our cohort^24^.

The presence and frequency of *NOTCH1* gene fusions can currently be regarded as a major molecular genetic difference between T-LBL and T-ALL, since fusions involving *NOTCH1* are only extremely rarely described in T-ALL (<0·1%)^21,26–31^. The uniform absence of BM and PB involvement in T-LBL patients with a *NOTCH1* gene fusion coincides with the fact that these rearrangements have almost never been detected in T-ALL. *NOTCH1* gene fusions have been described in T-LBL before^28^, but given the small number of samples in these studies, as well as difficulties in detecting TR-rearranged fusions with conventional pipelines, these fusions might have been missed explaining the lower contribution of *NOTCH1* gene fusions in these studies.

*NOTCH1* gene fusions appear to have more impact on T-LBL cells compared to *NOTCH1* mutations, with more and larger changes in the downstream gene expression and NOTCH1 activity, independent of the type of *NOTCH1*-fusion. Furthermore, whereas patients with *NOTCH1* and/or *FBXW7* mutations are considered low risk and have a better outcome compared to *NOTCH1* and/or *FBXW7* WT patients^11,12^, we demonstrate that the outcome of these recurrent *NOTCH1*-rearranged T-LBLs is poor. It has recently been described that *NOTCH1* intronic single nucleotide variants (SNVs) and *NOTCH1* intragenic losses were also associated with an inferior event-free and overall survival^31^, further substantiating that distinct genetic aberrations in *NOTCH1* have a different impact on outcome. The T-LBL patients with high-risk *NOTCH1* aberrations will probably need intensified or alternative treatment strategies. A second consequence of our findings may be that when *NOTCH1* gene fusions are recognized as a separate high-risk group, the survival characteristics of the remaining group of T-LBL patients with unknown molecular genetics improves and may benefit from less intensified treatment.

The highly elevated blood CCL17 levels were exclusively observed in all T-LBL patients with a *NOTCH1*-rearrangement, even though almost all T-LBL patients had an enlarged mediastinum. Elevated CCL17 levels may therefore serve as an important biomarker to assist in the diagnosis of this high-risk group at diagnosis. Moreover, there might be a possibility that CCL17 levels could be used during follow-up as well, but these findings need to be validated in larger cohorts. In classical Hodgkin lymphoma, It has been suggested that inhibiting CCL17, produced by the Reed-Sternberg cells, may have therapeutic consequences in classical Hodgkin lymphoma as inhibiting CCL17 can decrease the recruitment of T-cells, thereby affecting the supporting microenvironment^32^. Our data suggest that CCL17 protein levels in the tumor cells of *NOTCH1*-rearranged tumor cells is only slightly increased and rapidly secreted based on immunohistochemistry, while *CLL17* gene expression and blood levels are highly increased. This likely points towards active crosstalk between the tumor cells and the microenvironment^33^. It is therefore intriguing to further explore whether *NOTCH1*-rearranged T-LBLs are dependent on CCL17 expression and whether this would provide opportunities for targeted treatment in a potentially high-risk subgroup of T-LBL. Measuring blood CCL17 levels could also serve as an easily applicable technique to identify high-risk T-LBL patients in low- and middle-income countries with restricted access to next generation sequencing techniques.

In conclusion, we discovered that, in contrast to T-ALL, *NOTCH1* gene fusions are common in T-LBL and represent a high-risk subtype with an easily applicable biomarker. The discovery of this clinically relevant high-risk T-LBL subgroup offers opportunities to develop intensified and targeted treatment strategies for this subgroup and decrease overtreatment in the remaining group of T-LBL patients.

## Data Availability

Exome and transcriptome data are submitted to the European Genome-Phenome Archive (EGA) for controlled-access data sharing (EGAS00001007703)

## Data sharing agreement

Exome and transcriptome data will be submitted to the European Genome-Phenome Archive (EGA) for controlled-access data sharing (EGAS00001007703).

## Author contributions

RK, JLCL, AB designed the study. EK wrote the manuscript and analyzed the data. MMK and LAK performed the bioinformatic analyses. JGCB performed the wet lab analyses. ES provided laboratory supervision. MSV performed the histological analyses. MMH, FAGM, MAV were involved in data curation. JPPM acquired the funding.

## References

1. Minard-Colin V, Brugieres L, Reiter A, et al. Non-Hodgkin Lymphoma in Children and Adolescents: Progress Through Effective Collaboration, Current Knowledge, and Challenges Ahead. J Clin Oncol 2015;33(27):2963–74. DOI: 10.1200/JCO.2014.59.5827.

2. Burkhardt B, Reiter A, Landmann E, et al. Poor outcome for children and adolescents with progressive disease or relapse of lymphoblastic lymphoma: a report from the berlin-frankfurt-muenster group. J Clin Oncol 2009;27(20):3363–9. DOI: 10.1200/JCO.2008.19.3367.

3. Landmann E, Burkhardt B, Zimmermann M, et al. Results and conclusions of the European Intergroup EURO-LB02 trial in children and adolescents with lymphoblastic lymphoma. Haematologica 2017;102(12):2086–2096. DOI: 10.3324/haematol.2015.139162.

4. Swerdlow SH, Campo E, Lee Harris N, et al. WHO Classification of Tumours of Haematopoietic and Lymphoid Tissues (Revised 4th edition). World Health Organization Classification of Tumours. Lyon, France: International Agency for Research on Cancer (IARC); 2017.

5. Rosolen A, Perkins SL, Pinkerton CR, et al. Revised International Pediatric Non-Hodgkin Lymphoma Staging System. J Clin Oncol 2015;33(18):2112–8. DOI: 10.1200/JCO.2014.59.7203.

6. Khanam T, Sandmann S, Seggewiss J, et al. Integrative genomic analysis of pediatric T-cell lymphoblastic lymphoma reveals candidates of clinical significance. Blood 2020. DOI: 10.1182/blood.2020005381.

7. Bontoux C, Simonin M, Garnier N, et al. Oncogenetic landscape of T-cell lymphoblastic lymphomas compared to T-cell acute lymphoblastic leukemia. Mod Pathol 2022;35(9):1227–1235. DOI: 10.1038/s41379-022-01085-9.

8. Alaggio R, Amador C, Anagnostopoulos I, et al. The 5th edition of the World Health Organization Classification of Haematolymphoid Tumours: Lymphoid Neoplasms. Leukemia 2022;36(7):1720–1748. DOI: 10.1038/s41375-022-01620-2.

9. Zijtregtop EAM, Diez C, Zwaan CM, Veening MA, Beishuizen A, Meyer-Wentrup FAG. Thymus and activation-regulated chemokine (TARC) as treatment response marker for paediatric Hodgkin lymphoma: A pilot study. Br J Haematol 2023;200(1):70–78. DOI: 10.1111/bjh.18473.

10. Jundt F, Anagnostopoulos I, Forster R, Mathas S, Stein H, Dorken B. Activated Notch1 signaling promotes tumor cell proliferation and survival in Hodgkin and anaplastic large cell lymphoma. Blood 2002;99(9):3398–403. DOI: 10.1182/blood.v99.9.3398.

11. Bonn BR, Rohde M, Zimmermann M, et al. Incidence and prognostic relevance of genetic variations in T-cell lymphoblastic lymphoma in childhood and adolescence. Blood 2013;121(16):3153–60. DOI: 10.1182/blood-2012-12-474148.

12. Callens C, Baleydier F, Lengline E, et al. Clinical impact of NOTCH1 and/or FBXW7 mutations, FLASH deletion, and TCR status in pediatric T-cell lymphoblastic lymphoma. J Clin Oncol 2012;30(16):1966–73. DOI: 10.1200/JCO.2011.39.7661.

13. Hehir-Kwa JY, Koudijs MJ, Verwiel ETP, et al. Improved Gene Fusion Detection in Childhood Cancer Diagnostics Using RNA Sequencing. JCO Precis Oncol 2022;6:e2000504. DOI: 10.1200/PO.20.00504.

14. Haas BJ, Dobin A, Stransky N, et al. STAR-Fusion: Fast and Accurate Fusion Transcript Detection from RNA-Seq. BioRxiv 2017.

15. Love MI, Huber W, Anders S. Moderated estimation of fold change and dispersion for RNA-seq data with DESeq2. Genome Biol 2014;15(12):550. DOI: 10.1186/s13059-014-0550-8.

16. Yu G, Wang LG, Han Y, He QY. clusterProfiler: an R package for comparing biological themes among gene clusters. OMICS 2012;16(5):284–7. DOI: 10.1089/omi.2011.0118.

17. Subramanian A, Tamayo P, Mootha VK, et al. Gene set enrichment analysis: a knowledge-based approach for interpreting genome-wide expression profiles. Proc Natl Acad Sci U S A 2005;102(43):15545–50. DOI: 10.1073/pnas.0506580102.

18. Bene MC, Castoldi G, Knapp W, et al. Proposals for the immunological classification of acute leukemias. European Group for the Immunological Characterization of Leukemias (EGIL). Leukemia 1995;9(10):1783–6. (https://www.ncbi.nlm.nih.gov/pubmed/7564526).

19. Palomero T, Barnes KC, Real PJ, et al. CUTLL1, a novel human T-cell lymphoma cell line with t(7;9) rearrangement, aberrant NOTCH1 activation and high sensitivity to gamma-secretase inhibitors. Leukemia 2006;20(7):1279–87. DOI: 10.1038/sj.leu.2404258.

20. Steimle T, Dourthe ME, Alcantara M, et al. Clinico-biological features of T-cell acute lymphoblastic leukemia with fusion proteins. Blood Cancer J 2022;12(1):14. DOI: 10.1038/s41408-022-00613-9.

21. Brady SW, Roberts KG, Gu Z, et al. The genomic landscape of pediatric acute lymphoblastic leukemia. Nat Genet 2022;54(9):1376–1389. DOI: 10.1038/s41588-022-01159-z.

22. Kopan R, Ilagan MX. The canonical Notch signaling pathway: unfolding the activation mechanism. Cell 2009;137(2):216–33. DOI: 10.1016/j.cell.2009.03.045.

23. Ashworth TD, Pear WS, Chiang MY, et al. Deletion-based mechanisms of Notch1 activation in T-ALL: key roles for RAG recombinase and a conserved internal translational start site in Notch1. Blood 2010;116(25):5455–64. DOI: 10.1182/blood-2010-05-286328.

24. Patel JL, Smith LM, Anderson J, et al. The immunophenotype of T-lymphoblastic lymphoma in children and adolescents: a Children’s Oncology Group report. Br J Haematol 2012;159(4):454–61. DOI: 10.1111/bjh.12042.

25. Tasian SK, Loh ML, Hunger SP. Philadelphia chromosome-like acute lymphoblastic leukemia. Blood 2017;130(19):2064–2072. DOI: 10.1182/blood-2017-06-743252.

26. Ellisen LW, Bird J, West DC, et al. TAN-1, the human homolog of the Drosophila notch gene, is broken by chromosomal translocations in T lymphoblastic neoplasms. Cell 1991;66(4):649–61. DOI: 10.1016/0092-8674(91)90111-b.

27. Speleman F, Cauwelier B, Dastugue N, et al. A new recurrent inversion, inv(7)(p15q34), leads to transcriptional activation of HOXA10 and HOXA11 in a subset of T-cell acute lymphoblastic leukemias. Leukemia 2005;19(3):358–66. DOI: 10.1038/sj.leu.2403657.

28. Salmeron-Villalobos J, Ramis-Zaldivar JE, Balague O, et al. Diverse mutations and structural variations contribute to Notch signaling deregulation in paediatric T-cell lymphoblastic lymphoma. Pediatr Blood Cancer 2022;69(11):e29926. DOI: 10.1002/pbc.29926.

29. Yamamoto K, Nakamachi Y, Yakushijin K, et al. A novel TRB@/NOTCH1 fusion gene in T-cell lymphoblastic lymphoma with t(7;9)(q34;q34). Eur J Haematol 2013;90(1):68–75. DOI: 10.1111/ejh.12019.

30. Liu Y, Easton J, Shao Y, et al. The genomic landscape of pediatric and young adult T-lineage acute lymphoblastic leukemia. Nat Genet 2017;49(8):1211–1218. DOI: 10.1038/ng.3909.

31. Mullighan C, Pölönen P, Giacomo DD, et al. [preprint] The genomic basis of childhood T-lineage acute lymphoblastic leukemia. Research Square.

32. Zijtregtop EAM, van der Strate I, Beishuizen A, et al. Biology and Clinical Applicability of Plasma Thymus and Activation-Regulated Chemokine (TARC) in Classical Hodgkin Lymphoma. Cancers (Basel) 2021;13(4). DOI: 10.3390/cancers13040884.

33. Skinnider BF, Mak TW. The role of cytokines in classical Hodgkin lymphoma. Blood 2002;99(12):4283–97. DOI: 10.1182/blood-2002-01-0099.

