## Supplementary Appendix for "NOTCH1 fusions in pediatric T-cell lymphoblastic lymphoma: a high-risk subgroup with CCL17 (TARC) levels as diagnostic biomarker"

**JLCL and RPK are considered co-last authors*

**Table of Contents**

**Supplementary Figure 1:** Differences in gene expression.

**Supplementary Figure 2:** KEGG pathway enrichment analysis.

**Supplementary Table 1:** Demographics and clinical information about the RNAseq T-LBL cohort (n=29).

**Supplementary Table 2:** Detected fusions in T-LBL cohort.

**Supplementary Table 3:** 200 most variable genes in the complete T-LBL dataset.

**Supplementary Table 4:** Differentially expressed genes in *NOTCH1*-rearranged T-LBL versus *NOTCH1*-WT T-LBL (n=1,288).

**Supplementary Table 5:** Differentially expressed genes in *NOTCH1* mutated T-LBL versus *NOTCH1*-WT T-LBL (n=101).

**Supplementary Table 6:** Clinical information about the *NOTCH1*-rearranged patients.

**Supplementary Figures**

**Supplementary Figure 1: Differences in gene expression.** A) Unsupervised clustering with the 200 most variable genes in the dataset (based on the standard deviation) shows that most of the NOTCH1-rearranged samples cluster separate from the rest of the T-LBL samples.

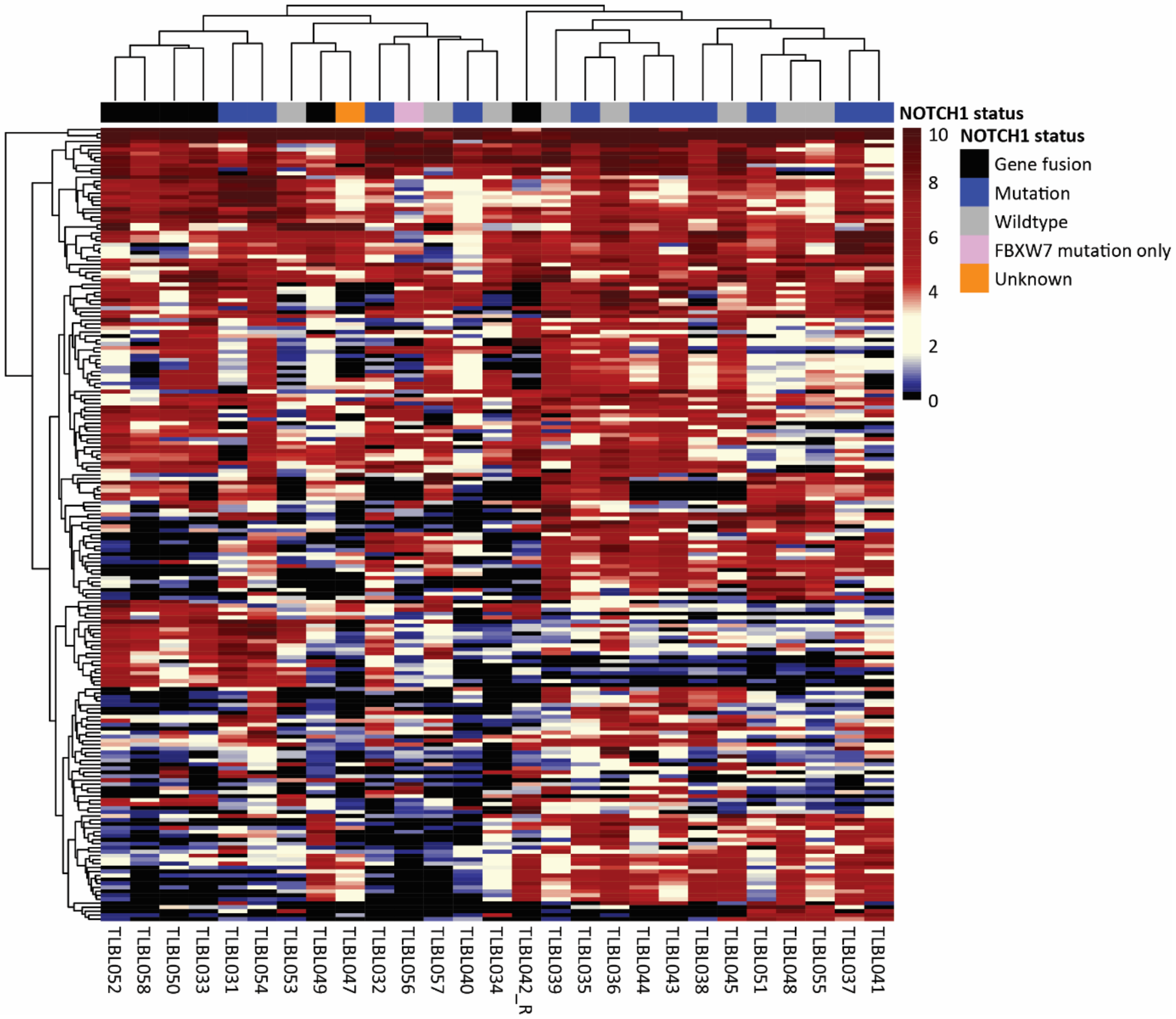

**Supplementary Figure 2: KEGG pathway enrichment analysis.** A) KEGG pathway enrichment analysis of NOTCH1-rearranged and NOTCH1-WT samples shows a large number of significantly enriched KEGG pathways in NOTCH1-rearranged T-LBL (FDR-adjusted p-value <0.05). B) KEGG pathway enrichment analysis of NOTCH1-mutated and NOTCH1-WT samples. Significantly enriched pathways are depicted (FDR-adjusted p-value < 0.05).

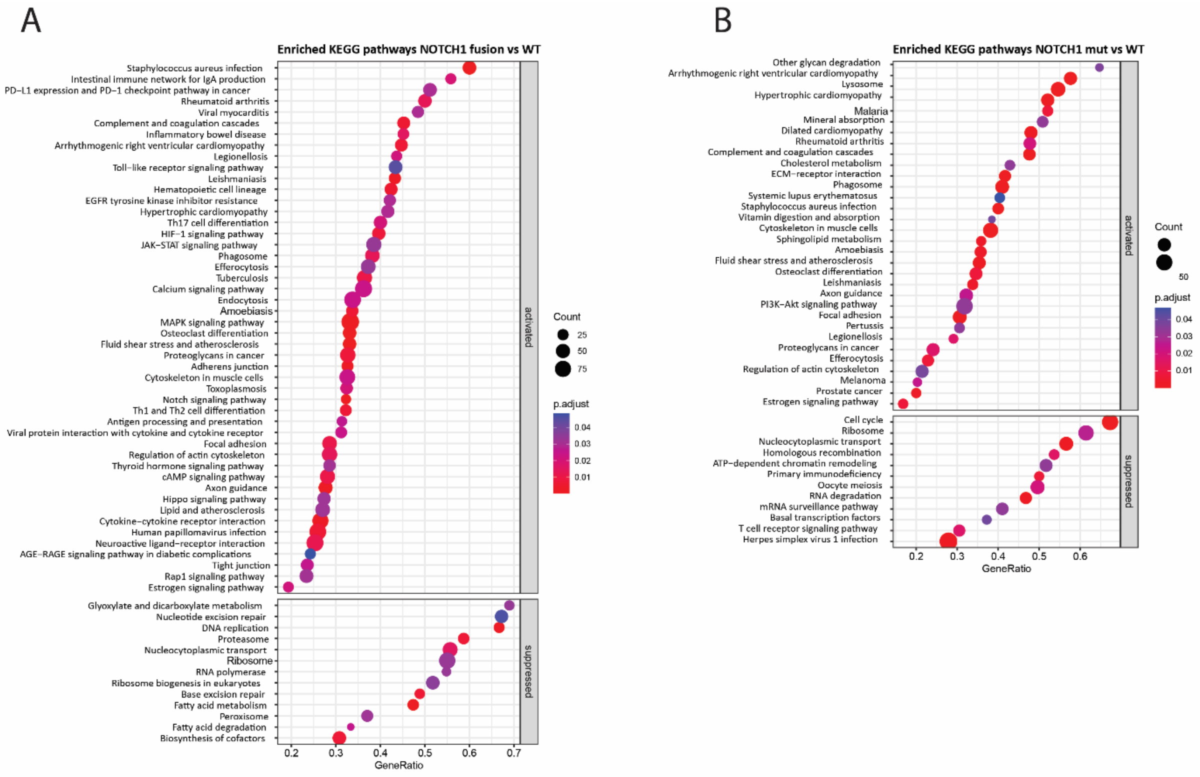

### Supplementary Tables

**Supplementary Table 1:** Demographics and clinical information about the RNAseq T-LBL cohort (n=29).

| Patient ID | Sex | Age (cat) | Mediastinal enlargement | Pleural effusion | Stage | Bone marrow (% blasts)* | Peripheral blood (% blasts)* | CNS | Material Analysis ** | Relapse | TARC measurement |
|---|---|---|---|---|---|---|---|---|---|---|---|
| **TLBL049** | male | 8-12 | + | + | III | - | - | CNS1 | LN | - | performed |
| TLBL046 | male | 13-18 | + | + | III | - | - | CNS1 | LN | - | performed |
| TLBL045 | female | 8-12 | + | - | III | - | - | CNS1 | LN | - | performed |
| TLBL047 | male | 13-18 | + | - | III | - | - | CNS1 | LN | - | performed |
| **TLBL033** | female | 13-18 | + | + | III | - | + | CNS1 | PLE | + | performed |
| TLBL044 | female | 13-18 | - | - | III | - | - | CNS2 | LN | - | performed |
| TLBL031 | male | 2-7 | + | + | III | - | - | CNS1 | PLE | - | |
| TLBL043 | female | 2-7 | + | - | III | + | + | CNS1 | LN | - | performed |
| TLBL032 | female | 13-18 | - | - | III | - | - | CNS1 | LN | - | performed |
| TLBL035 | male | 2-7 | + | - | III | - | - | CNS1 | LN | - | performed |
| TLBL034 | female | 2-7 | + | + | III | - | - | CNS1 | LN | - | |
| TLBL038 | female | 2-7 | + | - | IV | + | - | CNS2 | LN | - | performed |
| TLBL036 | male | 8-12 | + | - | III | + | - | CNS2 | LN | - | |
| TLBL037 | male | 8-12 | + | - | III | - | - | CNS1 | LN | - | performed |
| TLBL041 | male | 2-7 | + | - | III | - | - | CNS2 | LN | - | performed |
| **TLBL042** | female | 13-17 | + | + | III | - | - | CNS1 | LN | + | performed |
| TLBL040 | male | 13-17 | + | - | III | - | - | CNS1 | LN | - | performed |
| **TLBL050** | male | 8-12 | + | + | III | - | - | CNS1 | PLE | + | performed |
| TLBL051 | male | 8-12 | + | - | IV | - | - | CNS3 | LN | - | performed |
| **TLBL052** | male | 8-12 | + | + | III | - | - | CNS1 | PLE | - | performed |
| TLBL053 | female | 13-17 | + | - | III | - | + | CNS1 | PE | - | performed |
| TLBL048 | female | 2-7 | + | + | III | - | - | CNS1 | LN | - | performed |
| TLBL054 | male | 2-7 | + | + | III | - | - | CNS1 | PLE | - | |
| TLBL055 | male | 2-7 | - | - | IV | + | | CNS2 | LN | + | |
| TLBL056 | female | 2-7 | + | + | III | - | - | CNS1 | PLE | - | |
| TLBL059 | male | 8-12 | + | - | III | - | - | CNS1 | LN | - | performed |
| TLBL039 | male | 8-12 | + | - | IV | + | + | CNS1 | LN | - | performed |
| TLBL057 | male | 8-12 | + | + | IV | + | + | CNS2 | PLE | - | |
| **TLBL058** | male | 13-18 | + | + | III | - | - | CNS1 | LN | + | performed |

Patients with NOTCH1 fusions are **marked**.

*Flow positive =>1%

**LN=lymph node, PLE=pleural effusion, PE=pericardial effusion

**Supplementary Table 2:** Detected fusions in T-LBL cohort.

| Patient ID | Fusion | 5' gene | 5' breakpoint | 3' gene | 3' breakpoint |
|---|---|---|---|---|---|
| TLBL032 | *TRBC2::HOXA9* | *TRBC2* | not determined | *HOXA9* | not determined |
| TLBL040 | *NUP153::ABL1* | *NUP153* | not determined | *ABL1* | not determined |
| TLBL059 | *NUP214::ABL1* | *NUP214* | not determined | *ABL1* | not determined |
| TLBL053 | *MYB::PLAGL1* | *MYB* | not determined | *PLAGL1* | not determined |
| TLBL042 | *miR142::NOTCH1* | *miR142* | chr17:58,332,103 | *NOTCH1* | chr9:136,502,813 |
| TLBL058 | *miR142::NOTCH1* | *miR142* | chr17:58,331,375 | *NOTCH1* | chr9:136,504,127 |
| TLBL049 | *TRBJ::NOTCH1* | *TRBJ2-5* | chr7:142,797,124 | *NOTCH1* | chr9:136,505,623 |
| TLBL033 | *TRBJ::NOTCH1* | *TRBJ2-1* | chr7:142,796,366 | *NOTCH1* | chr9:136,505,820 |
| TLBL050 | *IKZF2::NOTCH1* | *IKZF2* | chr2:213,031,619 | *NOTCH1* | chr9:136,502,385 |
| TLBL052 | *TRBJ::NOTCH1* | *TRBJ2-1* | chr7:142,796,366 | *NOTCH1* | chr9:136,506,725 |
| TLBL034 | *TRBC1::MYC* | *TRBC1* | not determined | *MYC* | not determined |
| TLBL057 | *TRB::LMO1* | *TRB* | not determined | *LMO1* | not determined |

**Supplementary Table 3:** 200 most variable genes in the complete T-LBL dataset. The mean gene expressions and corresponding standard deviations are calculated from log2-transformed TPM values.

| Gene | Mean | Standard deviation |
|---|---|---|
| A2M | 5,282787629 | 2,164664973 |
| AC002454 | 3,937060273 | 2,881782226 |
| AC016735 | 1,552004062 | 2,451532851 |
| AC096631 | 4,156838597 | 2,210890315 |
| AC103833 | 3,464226897 | 2,156235497 |
| AC132807 | 3,038323594 | 2,711458948 |
| AC244205 | 2,384477286 | 2,21115946 |
| AC247036 | 3,564356394 | 2,771813906 |
| ADGRG1 | 2,98128235 | 2,369912253 |
| AL138899 | 4,679918986 | 2,718166912 |
| AL163932 | 3,985002852 | 2,863517933 |
| AL365440 | 3,067468483 | 2,375430629 |
| AL928646 | 1,313997144 | 2,248333146 |
| ALDH1A2 | 3,380087329 | 2,465559799 |
| ANKRD36BP2 | 3,961750911 | 2,536840756 |
| AP005273 | 2,871557707 | 2,370634433 |
| ARMH1 | 3,742437921 | 2,306135203 |
| BAHCC1 | 3,188419057 | 2,229633867 |
| BGN | 3,74225526 | 2,188797666 |
| BLK | 2,849452831 | 2,14444567 |
| BX571818 | 2,985979295 | 2,435525408 |
| C1QA | 5,010887187 | 2,33704289 |
| C1QB | 5,087074878 | 2,442529672 |
| C1QC | 5,745767121 | 2,488760628 |
| C7 | 2,868285978 | 2,816290968 |

| | | |
|---|---|---|
| CCL18 | 4,380325588 | 2,365554039 |
| CCL19 | 3,160929432 | 3,058457814 |
| CCL21 | 3,07446387 | 3,741530155 |
| CD163 | 4,571506835 | 2,403672132 |
| CD1A | 4,456241217 | 2,767631235 |
| CD1B | 6,066819082 | 3,164504322 |
| CD1E | 5,850478532 | 3,191460017 |
| CD40LG | 2,652781915 | 2,267611325 |
| CD8A | 5,203757847 | 2,293000535 |
| CD8B | 3,187578376 | 2,09971093 |
| CEMIP | 3,160989275 | 2,25808181 |
| CHI3L2 | 5,559782535 | 2,1749623 |
| CLDN1 | 1,954061622 | 2,307508607 |
| COL15A1 | 2,557909048 | 2,269887672 |
| COL1A1 | 5,856315209 | 2,468481178 |
| COL1A2 | 5,498795549 | 2,338232684 |
| COL3A1 | 6,61167353 | 2,435431646 |
| COL4A1 | 3,061529348 | 2,713465843 |
| COL4A2 | 2,635406978 | 2,337415804 |
| COL6A3 | 3,521654448 | 2,190777388 |
| CR2 | 4,954615278 | 2,457991768 |
| CR392039 | 7,545050404 | 2,256692392 |
| CSMD1 | 1,712870005 | 2,378376531 |
| CTSL | 5,788361431 | 2,279968758 |
| CXCL13 | 2,620789294 | 2,622499454 |
| CXCL8 | 1,924028809 | 2,131642891 |
| DDX3Y | 2,925999109 | 2,543078805 |
| DNTT | 6,860332542 | 2,94006236 |
| DPP4 | 3,010599031 | 2,108938004 |
| DTX1 | 3,374912684 | 2,240097055 |
| EGFL6 | 1,884685414 | 2,210031068 |
| EIF1AY | 2,512959681 | 2,272201559 |
| ELOVL4 | 3,571479873 | 2,105361813 |
| FABP4 | 2,90633099 | 2,241572462 |
| FCGR3A | 3,116518306 | 2,218742557 |
| FDCSP | 3,373470788 | 3,265782395 |
| FN1 | 6,031477218 | 2,567291569 |
| FP236383 | 12,73500998 | 3,291168765 |
| FP671120 | 12,37170804 | 3,110624318 |
| GOLGA8O | 2,865215647 | 2,130802431 |
| GTSF1 | 2,014099267 | 2,182714237 |
| GZMA | 2,791139533 | 2,11836449 |
| HBA1 | 4,888876898 | 2,756353117 |
| HBA2 | 5,056111251 | 2,719902774 |
| HBB | 4,319940438 | 2,713204904 |
| HES4 | 3,823358626 | 2,385075685 |

| | | |
|---|---|---|
| HIST1H1A | 2,056568394 | 2,19803037 |
| HIST1H2BB | 7,74395767 | 2,23725758 |
| HIST1H2BM | 7,247268104 | 2,329720229 |
| HIST1H3J | 6,577101503 | 2,306563285 |
| HIST1H4F | 8,92402628 | 2,797624155 |
| HIST1H4L | 7,736816169 | 2,102473469 |
| HIST3H2A | 3,465910617 | 2,543722782 |
| HIST3H2BB | 2,777769831 | 2,229813328 |
| HSPG2 | 3,321925395 | 2,195688362 |
| IGFBP7 | 4,629239506 | 2,575866425 |
| IGHGP | 2,809473515 | 2,713447123 |
| IGHJ1P | 1,663562284 | 2,332567317 |
| IGHJ3P | 2,700411918 | 2,103524624 |
| IGHV1-14 | 2,052638014 | 2,438139949 |
| IGHV3-19 | 3,2506312 | 2,774185145 |
| IGHVII-40-1 | 2,300167407 | 2,29454162 |
| IGLC6 | 1,855064731 | 2,250288001 |
| IGLL1 | 2,552306608 | 2,878235434 |
| IGLL5 | 3,198589126 | 2,728507189 |
| IL32 | 5,804208806 | 2,161702435 |
| ITLN1 | 1,791037614 | 2,474513992 |
| JCHAIN | 3,528644309 | 2,613707665 |
| KDM5D | 2,492567038 | 2,155999414 |
| KRT19 | 1,865168619 | 2,308620509 |
| LINC01221 | 4,460769115 | 2,525210354 |
| LINC01222 | 3,964760195 | 2,234868086 |
| LINC01835 | 1,819880212 | 2,984087742 |
| LUM | 2,804060588 | 2,13380597 |
| LYZ | 5,687659544 | 2,145230629 |
| MAL | 6,413768049 | 2,57808308 |
| MARCO | 3,446664001 | 2,866111123 |
| MMP9 | 4,13396752 | 2,432560681 |
| MT-RNR1 | 5,34208316 | 3,457504332 |
| MT-RNR2 | 5,135913947 | 3,384774106 |
| MT-TC | 10,72693147 | 2,099159117 |
| MT1E | 3,364149411 | 2,147744474 |
| MT1G | 3,155681631 | 2,667859796 |
| MT1H | 1,598781313 | 2,445067652 |
| MYO18B | 1,559304748 | 2,623923008 |
| MYO7B | 4,393001179 | 2,871467884 |
| NDST3 | 2,528622304 | 2,254697173 |
| NRN1 | 3,560197363 | 2,461401441 |
| PGBD4P1 | 1,771369689 | 2,528375453 |
| PLA2G2A | 2,795527353 | 2,367864163 |
| PLA2G2D | 2,534765421 | 2,171654098 |
| PLTP | 5,234328885 | 2,141434124 |

| | | |
|---|---|---|
| PLVAP | 2,482499884 | 2,36885274 |
| POSTN | 2,712224532 | 2,481781744 |
| PRG4 | 2,152284904 | 2,360873407 |
| PRSS57 | 1,231638481 | 2,399147847 |
| PTCRA | 3,820203699 | 2,437013731 |
| RAG1 | 3,460274976 | 2,279016853 |
| RCBTB2 | 4,734440297 | 2,146626731 |
| RF00612 | 2,235988549 | 2,142383932 |
| RNASE1 | 4,836766052 | 2,379174785 |
| RNU5B-1 | 3,843923065 | 2,203103713 |
| RPS3AP5 | 3,066709878 | 2,333264686 |
| RPS4Y1 | 4,462665662 | 3,809684925 |
| S100A8 | 2,956114493 | 2,621788398 |
| S100A9 | 4,897742632 | 2,509503226 |
| SAA1 | 2,030588976 | 2,345346507 |
| SCGB3A1 | 2,38348183 | 2,480673342 |
| SELL | 5,873244589 | 2,280080055 |
| SERPINB2 | 1,482976425 | 2,097012239 |
| SIRPG | 2,770013028 | 2,105718164 |
| SLPI | 3,682693264 | 3,791450091 |
| SPARCL1 | 3,434718394 | 2,682888686 |
| SPINK2 | 2,191268209 | 2,189281062 |
| SPP1 | 5,644929423 | 3,530911249 |
| TMSB15A | 3,101164823 | 2,156004336 |
| TNC | 2,6192779 | 2,160664281 |
| TRAJ21 | 3,763076839 | 2,116607261 |
| TRAJ22 | 3,303174019 | 2,142318503 |
| TRAJ27 | 3,366667378 | 2,239469889 |
| TRAJ29 | 3,394468233 | 2,142602138 |
| TRAJ30 | 2,95024205 | 2,164456609 |
| TRAJ31 | 3,024673787 | 2,199676654 |
| TRAJ32 | 3,189703657 | 2,427979468 |
| TRAJ33 | 3,233756424 | 2,512799791 |
| TRAJ34 | 2,769110261 | 2,110287172 |
| TRAJ36 | 2,485624437 | 2,096517324 |
| TRAJ37 | 2,661327181 | 2,182286398 |
| TRAJ56 | 1,800861851 | 2,209771367 |
| TRAJ57 | 1,754301575 | 2,122108113 |
| TRAJ61 | 1,471253866 | 2,105293159 |
| TRAV1-1 | 2,387455683 | 3,370875553 |
| TRBD1 | 4,567080224 | 2,926888403 |
| TRBJ1-1 | 3,802152722 | 3,557290303 |
| TRBJ1-2 | 4,573471173 | 3,270528393 |
| TRBJ1-3 | 4,799172117 | 3,202709572 |
| TRBJ1-4 | 5,010967019 | 3,167362743 |
| TRBJ1-5 | 4,83809246 | 3,016335624 |

| | | |
|---|---|---|
| TRBJ1-6 | 5,322437318 | 2,980192802 |
| TRBJ2-1 | 5,344460118 | 2,77805447 |
| TRBV10-3 | 2,07292069 | 2,601528345 |
| TRBV14 | 1,681134215 | 2,176423144 |
| TRBV18 | 1,752922068 | 2,324223067 |
| TRBV20-1 | 2,823124083 | 2,271016984 |
| TRBV21-1 | 1,927141066 | 2,256612075 |
| TRBV24-1 | 1,260158335 | 2,13553068 |
| TRBV28 | 4,132752754 | 3,545904825 |
| TRBV30 | 3,338739682 | 2,137776635 |
| TRBV4-2 | 1,590426634 | 2,150251243 |
| TRBV5-6 | 2,208711647 | 2,519023269 |
| TRBV6-1 | 1,286368821 | 2,269643225 |
| TRBV6-2 | 1,377597783 | 2,264809766 |
| TRBV6-6 | 1,666604 | 2,227744451 |
| TRBV7-9 | 2,903552792 | 2,726992408 |
| TRDC | 4,393480467 | 3,598562131 |
| TRDD3 | 1,399177428 | 2,667097502 |
| TRDJ1 | 3,201662291 | 3,209681218 |
| TRDJ2 | 2,517902704 | 2,325083674 |
| TRDJ3 | 3,153665051 | 2,99848127 |
| TRDJ4 | 2,090036427 | 2,298199649 |
| TRDV1 | 2,405986909 | 2,590468779 |
| TRGC1 | 3,200389399 | 2,473499936 |
| TRGC2 | 5,960323119 | 2,509953117 |
| TRGJ1 | 6,568404058 | 2,277649469 |
| TRGJ2 | 4,960344875 | 2,321163276 |
| TRGJP1 | 2,778987319 | 3,073511614 |
| TRGJP2 | 3,366602498 | 3,601429507 |
| TRGV4 | 3,220254095 | 2,240900444 |
| TSPAN7 | 4,963957912 | 2,113848108 |
| UGT3A2 | 2,376156373 | 2,248709956 |
| UTY | 2,507652653 | 2,16221888 |
| VSIG4 | 3,233007421 | 2,489419044 |
| VWF | 3,047171166 | 2,847176932 |
| XG | 2,232692758 | 2,833459152 |
| XIST | 1,738560588 | 2,476133428 |

**Supplementary Table 4:** Differentially expressed genes in *NOTCH1*-rearranged T-LBL versus *NOTCH1*-WT T-LBL (n=1,288). Genes with an FDR-adjusted p-value ≤ 0.05 and an absolute log2-transformed log fold change of at least 1 were considered significantly differentially expressed.

| Gene | log2FoldChange | P-value | FDR-adjusted p-value | Expression |
|---|---|---|---|---|
| AC000078 | 1,923490265 | 0,000239421 | 0,01226962 | Upregulated |
| AC004039 | 2,267746802 | 0,000108382 | 0,007035436 | Upregulated |
| AC005077 | 3,726605114 | 0,001338422 | 0,037690993 | Upregulated |
| AC005593 | 1,552628443 | 0,002037263 | 0,049023733 | Upregulated |
| AC006042 | 1,428321701 | 0,001334623 | 0,037669225 | Upregulated |
| AC007422 | 4,87542061 | 7,43042E-05 | 0,005302246 | Upregulated |
| AC007601 | 1,486111807 | 0,001357246 | 0,037999338 | Upregulated |
| AC007780 | 2,081636594 | 7,84503E-05 | 0,005491006 | Upregulated |
| AC008534 | 2,420538393 | 0,000383053 | 0,016802675 | Upregulated |
| AC008608 | 1,899288094 | 1,94991E-05 | 0,001961912 | Upregulated |
| AC008868 | 2,907678683 | 0,000174871 | 0,009826041 | Upregulated |
| AC009271 | 5,320007224 | 0,000908648 | 0,029255735 | Upregulated |
| AC011595 | 3,589794276 | 0,001112319 | 0,033564522 | Upregulated |
| AC015967 | 1,332231475 | 0,00068368 | 0,024677641 | Upregulated |
| AC016598 | 4,206154595 | 0,000348306 | 0,015791085 | Upregulated |
| AC016738 | 2,584901664 | 0,000413015 | 0,017613018 | Upregulated |
| AC016987 | 7,506598251 | 0,000525679 | 0,02067155 | Upregulated |
| AC017002 | 2,828179452 | 1,40716E-06 | 0,000281501 | Upregulated |
| AC017028 | 2,120883921 | 0,001453239 | 0,039787372 | Upregulated |
| AC023787 | 3,938991423 | 9,65996E-07 | 0,000211579 | Upregulated |
| AC025178 | 1,296479101 | 0,001581693 | 0,042052669 | Upregulated |
| AC025186 | 2,428451989 | 0,00050229 | 0,019945235 | Upregulated |
| AC025437 | 7,204182196 | 4,79879E-09 | 2,71064E-06 | Upregulated |
| AC026310 | 2,354646844 | 0,001837725 | 0,046066437 | Upregulated |
| AC027288 | 2,801253758 | 0,000553593 | 0,021500632 | Upregulated |
| AC027329 | 5,188349871 | 6,93196E-05 | 0,005072461 | Upregulated |
| AC055874 | 6,656505465 | 0,000510412 | 0,020164077 | Upregulated |
| AC063976 | 2,872120312 | 0,000274325 | 0,01352594 | Upregulated |
| AC066616 | 3,2655999 | 0,002049886 | 0,049107284 | Upregulated |
| AC069120 | 3,283886138 | 0,000206767 | 0,011011161 | Upregulated |
| AC073488 | 5,637954609 | 0,000677206 | 0,024535852 | Upregulated |
| AC074143 | 2,509546388 | 8,09173E-06 | 0,001042118 | Upregulated |
| AC074389 | 3,50879631 | 0,000136383 | 0,008253996 | Upregulated |
| AC078850 | 3,42201041 | 0,001310623 | 0,037376565 | Upregulated |
| AC079331 | 1,276634055 | 0,00012154 | 0,00761327 | Upregulated |
| AC087564 | 5,98565254 | 0,000166711 | 0,009619318 | Upregulated |
| AC090515 | 1,133604618 | 0,000361196 | 0,01624222 | Upregulated |
| AC090518 | 4,565693508 | 6,92365E-08 | 2,397E-05 | Upregulated |
| AC090673 | 8,46560686 | 3,18101E-08 | 1,31306E-05 | Upregulated |
| AC090826 | 2,25814542 | 5,02953E-07 | 0,000123615 | Upregulated |
| AC090877 | 2,712709663 | 1,41638E-06 | 0,000281501 | Upregulated |

| | | | | |
|---|---|---|---|---|
| AC091027 | 5,279450501 | 4,17099E-05 | 0,003488138 | Upregulated |
| AC091100 | 4,335689399 | 1,12514E-05 | 0,001331847 | Upregulated |
| AC091979 | 5,560370162 | 2,23948E-05 | 0,002218604 | Upregulated |
| AC092078 | 6,106332063 | 1,26754E-05 | 0,001431965 | Upregulated |
| AC092168 | 3,783239561 | 4,98307E-06 | 0,000760379 | Upregulated |
| AC092448 | 9,852843388 | 5,68061E-13 | 1,21932E-09 | Upregulated |
| AC092902 | 2,650314627 | 0,000206906 | 0,011011161 | Upregulated |
| AC092903 | 2,481830206 | 0,000615429 | 0,023121304 | Upregulated |
| AC093155 | 1,538227373 | 0,000222605 | 0,011634685 | Upregulated |
| AC093166 | 1,873605818 | 0,000124104 | 0,007710386 | Upregulated |
| AC093270 | 3,223034927 | 0,001440628 | 0,039509273 | Upregulated |
| AC093281 | 4,012386183 | 0,001550512 | 0,041566901 | Upregulated |
| AC096921 | 1,73274619 | 0,001208715 | 0,035571166 | Upregulated |
| AC097639 | 2,160124806 | 0,000723966 | 0,025558696 | Upregulated |
| AC103843 | 4,035215098 | 1,7643E-05 | 0,001832424 | Upregulated |
| AC104078 | 2,105847363 | 0,001327163 | 0,037629821 | Upregulated |
| AC105046 | 2,686988419 | 9,39845E-05 | 0,006278046 | Upregulated |
| AC106798 | 7,675206669 | 5,17264E-06 | 0,000774621 | Upregulated |
| AC107021 | 2,911114337 | 4,38306E-05 | 0,003618497 | Upregulated |
| AC107074 | 5,743117495 | 0,000672926 | 0,024495734 | Upregulated |
| AC107308 | 9,67827823 | 7,34095E-06 | 0,000986295 | Upregulated |
| AC108058 | 1,949738843 | 0,000104729 | 0,006903253 | Upregulated |
| AC110619 | 4,2133748 | 8,17522E-05 | 0,005672795 | Upregulated |
| AC110772 | 6,556603123 | 2,87809E-05 | 0,002632549 | Upregulated |
| AC112206 | 4,827735676 | 0,001157217 | 0,034498984 | Upregulated |
| AC112721 | 2,9964051 | 0,000463887 | 0,018924703 | Upregulated |
| AC117488 | 4,637469598 | 0,000454771 | 0,018798733 | Upregulated |
| AC117528 | 1,669220887 | 0,000860606 | 0,028332243 | Upregulated |
| AC121251 | 2,194720124 | 0,001187741 | 0,035213359 | Upregulated |
| AC124068 | 2,799082238 | 0,000684514 | 0,024680076 | Upregulated |
| AC127526 | 3,925633903 | 1,49079E-05 | 0,001609267 | Upregulated |
| AC131160 | 2,20696803 | 0,000178564 | 0,009964008 | Upregulated |
| AC135893 | 2,281403148 | 0,000753955 | 0,026271734 | Upregulated |
| AC145676 | 2,56070469 | 0,001543653 | 0,041556009 | Upregulated |
| AC243830 | 1,119388317 | 0,001196822 | 0,03541736 | Upregulated |
| ACVR2A | 2,01809439 | 0,000105059 | 0,006903253 | Upregulated |
| ADAM8 | 2,092886955 | 0,000380517 | 0,016759913 | Upregulated |
| ADAMTS6 | 2,769504693 | 1,23545E-08 | 6,11966E-06 | Upregulated |
| ADGRV1 | 2,531506576 | 8,24528E-05 | 0,005687879 | Upregulated |
| ADORA2B | 2,231223811 | 2,02809E-05 | 0,002034219 | Upregulated |
| AF165147 | 3,056747449 | 1,92996E-07 | 5,67827E-05 | Upregulated |
| AGAP3 | 1,849491809 | 0,000190997 | 0,010489444 | Upregulated |
| AGPAT3 | 1,840607478 | 5,44624E-05 | 0,00425825 | Upregulated |
| AGRN | 2,463848424 | 0,000119882 | 0,007525115 | Upregulated |
| AHI1 | 1,374164562 | 0,000177395 | 0,009933208 | Upregulated |
| AL008707 | 4,603934041 | 0,000819034 | 0,027442834 | Upregulated |

| | | | | |
|---|---|---|---|---|
| AL049629 | 1,942875819 | 0,00077578 | 0,026714228 | Upregulated |
| AL079301 | 3,548613825 | 0,000958511 | 0,030196838 | Upregulated |
| AL109955 | 1,862422802 | 0,002028144 | 0,0488408 | Upregulated |
| AL117342 | 2,685922593 | 0,000223519 | 0,011645063 | Upregulated |
| AL121782 | 2,369341195 | 0,001097063 | 0,033225415 | Upregulated |
| AL121820 | 2,580233999 | 1,36798E-05 | 0,001524043 | Upregulated |
| AL133268 | 1,627999596 | 0,000666637 | 0,024335278 | Upregulated |
| AL133492 | 2,394576034 | 0,000761757 | 0,026400741 | Upregulated |
| AL133493 | 2,783399945 | 0,001216173 | 0,035629783 | Upregulated |
| AL136369 | 4,973219446 | 5,67177E-09 | 3,09515E-06 | Upregulated |
| AL136961 | 3,759618703 | 0,000555492 | 0,021538914 | Upregulated |
| AL136968 | 2,036230344 | 0,001548919 | 0,04155878 | Upregulated |
| AL137024 | 6,116778947 | 2,20823E-06 | 0,000408612 | Upregulated |
| AL137783 | 3,550672384 | 0,001085258 | 0,033057745 | Upregulated |
| AL138479 | 4,87082614 | 4,13789E-09 | 2,4857E-06 | Upregulated |
| AL157395 | 1,758834106 | 0,000229667 | 0,011907568 | Upregulated |
| AL158168 | 4,445531056 | 0,001936027 | 0,047547101 | Upregulated |
| AL160254 | 3,605823611 | 0,000489329 | 0,019620086 | Upregulated |
| AL162408 | 3,276925718 | 0,000160189 | 0,009411709 | Upregulated |
| AL162464 | 7,222140391 | 0,000600245 | 0,022763374 | Upregulated |
| AL162615 | 1,393449161 | 0,000560472 | 0,021663279 | Upregulated |
| AL162727 | 2,394620164 | 0,000269269 | 0,013375123 | Upregulated |
| AL163952 | 4,333371997 | 1,66724E-05 | 0,001760008 | Upregulated |
| AL163953 | 4,426906985 | 3,07164E-06 | 0,000520772 | Upregulated |
| AL354890 | 2,710242583 | 0,001215407 | 0,035629783 | Upregulated |
| AL355377 | 1,518115187 | 0,001070501 | 0,032732131 | Upregulated |
| AL355773 | 3,885771653 | 8,64663E-05 | 0,005894084 | Upregulated |
| AL355881 | 4,93454981 | 1,78961E-05 | 0,001852732 | Upregulated |
| AL357075 | 2,829334342 | 0,001101116 | 0,033288842 | Upregulated |
| AL357874 | 1,551547959 | 6,71117E-05 | 0,004967343 | Upregulated |
| AL359813 | 2,236650084 | 0,000333743 | 0,015394742 | Upregulated |
| AL359962 | 1,564558131 | 0,000525825 | 0,02067155 | Upregulated |
| AL360182 | 2,128472199 | 0,000482872 | 0,019433772 | Upregulated |
| AL390726 | 8,137744735 | 3,38265E-06 | 0,000561398 | Upregulated |
| AL390760 | 4,701613603 | 0,001926386 | 0,047394696 | Upregulated |
| AL445471 | 2,456258539 | 0,001803617 | 0,045635071 | Upregulated |
| AL450332 | 5,138147572 | 0,00164678 | 0,043036831 | Upregulated |
| AL451047 | 6,005709804 | 3,07316E-06 | 0,000520772 | Upregulated |
| AL512622 | 5,211293576 | 9,13998E-07 | 0,000201562 | Upregulated |
| AL590807 | 2,582019665 | 0,001815698 | 0,045765925 | Upregulated |
| AL591721 | 2,123545306 | 0,000534074 | 0,020916949 | Upregulated |
| AL627308 | 4,942002089 | 9,16793E-06 | 0,001162125 | Upregulated |
| AL645608 | 2,899971644 | 0,000601364 | 0,022778976 | Upregulated |
| AL691420 | 7,014845591 | 3,23329E-05 | 0,002896588 | Upregulated |
| ALDH8A1 | 2,158412632 | 0,000377315 | 0,016664492 | Upregulated |
| ALK | 1,321557383 | 0,001722176 | 0,044323653 | Upregulated |

| | | | | |
|---|---|---|---|---|
| AMDHD1 | 1,430431765 | 0,001584246 | 0,042085782 | Upregulated |
| ANAPC1P1 | 2,345050386 | 0,000316841 | 0,014892445 | Upregulated |
| ANGPTL4 | 3,112283394 | 0,000469297 | 0,019054174 | Upregulated |
| ANGPTL6 | 2,116393966 | 0,000629999 | 0,02346311 | Upregulated |
| ANGPTL7 | 2,235825976 | 5,2371E-05 | 0,004153179 | Upregulated |
| ANKRD18DP | 3,590185382 | 0,000295841 | 0,014259286 | Upregulated |
| ANKRD34B | 4,11490527 | 0,001815781 | 0,045765925 | Upregulated |
| AP000722 | 5,532338784 | 0,00059078 | 0,022563865 | Upregulated |
| AP000864 | 3,393239918 | 2,46652E-05 | 0,002376465 | Upregulated |
| AP000870 | 3,37691435 | 0,001590479 | 0,042140991 | Upregulated |
| AP002813 | 2,089011934 | 0,000557716 | 0,021582682 | Upregulated |
| AP002954 | 2,852620761 | 2,5197E-05 | 0,002400201 | Upregulated |
| AP003063 | 3,731716864 | 1,39114E-05 | 0,001533926 | Upregulated |
| AP003100 | 5,95530859 | 1,16666E-05 | 0,001360655 | Upregulated |
| AP003718 | 5,154874495 | 0,000819859 | 0,027442834 | Upregulated |
| AP004608 | 5,240377377 | 2,06249E-05 | 0,00205591 | Upregulated |
| AP004609 | 1,250623387 | 0,001301863 | 0,037159637 | Upregulated |
| AP005209 | 5,013062568 | 1,20085E-05 | 0,001380849 | Upregulated |
| APCDD1 | 5,98516772 | 2,62473E-15 | 1,20726E-11 | Upregulated |
| APLP1 | 2,839695653 | 0,000250152 | 0,012663759 | Upregulated |
| APOBEC3A | 3,904172172 | 4,33075E-05 | 0,003603031 | Upregulated |
| AR | 2,619022302 | 0,001327681 | 0,037629821 | Upregulated |
| ARC | 3,81704307 | 0,0005376 | 0,020980726 | Upregulated |
| ARHGEF4 | 2,435045023 | 0,000284605 | 0,013905031 | Upregulated |
| ARSG | 2,777958291 | 4,02723E-08 | 1,62081E-05 | Upregulated |
| ASTN1 | 3,472562714 | 0,00064522 | 0,023827449 | Upregulated |
| ATF5 | 2,358012225 | 0,00137241 | 0,038289537 | Upregulated |
| ATP2B1 | 1,299075158 | 0,001546061 | 0,04155878 | Upregulated |
| AVPI1 | 1,780557952 | 6,32062E-05 | 0,004782966 | Upregulated |
| B3GNT4 | 1,132660242 | 0,001817923 | 0,045765925 | Upregulated |
| B4GALT1 | 2,083551058 | 9,55394E-06 | 0,001194458 | Upregulated |
| BAIAP2 | 1,820282563 | 0,001623863 | 0,042645608 | Upregulated |
| BBOX1-AS1 | 4,349969811 | 0,001626049 | 0,042668211 | Upregulated |
| BCHE | 4,175247379 | 0,000878542 | 0,028688058 | Upregulated |
| BCL6 | 2,154806982 | 2,27727E-05 | 0,002242238 | Upregulated |
| BICDL2 | 2,825578998 | 0,000110578 | 0,007120573 | Upregulated |
| BMP4 | 6,046065465 | 1,02144E-16 | 5,48122E-13 | Upregulated |
| BRAFP1 | 2,500878747 | 0,001480477 | 0,040361491 | Upregulated |
| C11orf91 | 2,397694046 | 0,001603988 | 0,042293965 | Upregulated |
| C11orf96 | 3,118955082 | 1,34416E-06 | 0,000272188 | Upregulated |
| C12orf29 | 2,805289355 | 2,11435E-07 | 6,07818E-05 | Upregulated |
| C12orf50 | 3,162463832 | 0,000313182 | 0,014767625 | Upregulated |
| C15orf48 | 4,30685085 | 1,10583E-05 | 0,001329354 | Upregulated |
| C17orf58 | 1,550105834 | 0,000492238 | 0,019712187 | Upregulated |
| C1orf116 | 3,316090872 | 0,000869058 | 0,028493943 | Upregulated |
| C8A | 4,723472939 | 0,001844683 | 0,046066437 | Upregulated |

| | | | | |
|---|---|---|---|---|
| C8B | 4,48556765 | 5,09421E-06 | 0,000770039 | Upregulated |
| CACNA1E | 5,994536537 | 1,5187E-09 | 1,16423E-06 | Upregulated |
| CADPS | 7,220813563 | 5,68634E-06 | 0,000825879 | Upregulated |
| CAMK2B | 3,117935423 | 0,000595506 | 0,022690555 | Upregulated |
| CAMK2N1 | 2,176443093 | 0,000625823 | 0,023348352 | Upregulated |
| CAMKK1 | 1,466396338 | 0,002006241 | 0,048548525 | Upregulated |
| CAP2 | 3,120399133 | 0,000805726 | 0,027307325 | Upregulated |
| CARTPT | 22,70300035 | 1,01545E-08 | 5,18959E-06 | Upregulated |
| CASC1 | 1,313202616 | 0,000816549 | 0,027442834 | Upregulated |
| CASKIN1 | 2,057705688 | 0,001527606 | 0,041348661 | Upregulated |
| CCDC198 | 5,217409993 | 1,07339E-12 | 1,91999E-09 | Upregulated |
| CCDC80 | 2,733164475 | 0,00201936 | 0,048702123 | Upregulated |
| CCL26 | 4,426919694 | 0,001313187 | 0,037383438 | Upregulated |
| CCNI2 | 3,528965949 | 0,000148666 | 0,008862924 | Upregulated |
| CCNJP2 | 2,27158266 | 0,001510419 | 0,041004195 | Upregulated |
| CCR9 | 2,366032594 | 0,000208299 | 0,011066991 | Upregulated |
| CD1A | 3,774838617 | 0,001201851 | 0,035533524 | Upregulated |
| CD1C | 4,261316711 | 7,93653E-08 | 2,64238E-05 | Upregulated |
| CD1D | 3,678030929 | 1,15553E-07 | 3,64752E-05 | Upregulated |
| CD72 | 2,265406901 | 0,000748494 | 0,026166396 | Upregulated |
| CDC42BPA | 3,825745318 | 8,00393E-07 | 0,000180284 | Upregulated |
| CDCP1 | 2,680808254 | 0,001739043 | 0,044579584 | Upregulated |
| CDK17 | 1,011624546 | 1,18208E-05 | 0,001365967 | Upregulated |
| CDK5R1 | 2,579450601 | 2,65256E-05 | 0,002484412 | Upregulated |
| CDKL5 | 1,595563559 | 0,000460114 | 0,018888421 | Upregulated |
| CELF2-AS2 | 3,135621882 | 0,000389266 | 0,016936751 | Upregulated |
| CHD7 | 1,891086852 | 0,002045293 | 0,04907576 | Upregulated |
| CHGA | 1,989697429 | 0,000722908 | 0,025549375 | Upregulated |
| CHI3L1 | 2,998810037 | 0,001002943 | 0,031169659 | Upregulated |
| CHIAP1 | 3,142946868 | 0,000347685 | 0,015791085 | Upregulated |
| CHIT1 | 3,090592584 | 0,000198873 | 0,01076156 | Upregulated |
| CHRNA2 | 5,646142305 | 4,25298E-17 | 2,73867E-13 | Upregulated |
| CHRNB4 | 2,756189325 | 0,000808594 | 0,027328477 | Upregulated |
| CIART | 3,82342857 | 3,90253E-06 | 0,000625122 | Upregulated |
| CKB | 2,331228118 | 7,49138E-05 | 0,005312775 | Upregulated |
| CLCN3P1 | 3,706309792 | 0,000242452 | 0,012351632 | Upregulated |
| CLDN1 | 5,481791329 | 4,9053E-05 | 0,003958297 | Upregulated |
| CLEC19A | 5,287332656 | 0,001714914 | 0,044242852 | Upregulated |
| CLIC5 | 3,87448986 | 0,00024429 | 0,012425594 | Upregulated |
| CLTCL1 | 2,027071343 | 0,000906606 | 0,029219202 | Upregulated |
| CMAHP | 1,964874325 | 0,000141531 | 0,008523835 | Upregulated |
| CMYA5 | 3,325528603 | 1,93996E-07 | 5,67827E-05 | Upregulated |
| CNKSR3 | 2,440788923 | 0,000803946 | 0,027275726 | Upregulated |
| CNMD | 2,504643484 | 0,000429157 | 0,018128734 | Upregulated |
| CNTN6 | 4,958554083 | 0,000376329 | 0,016654642 | Upregulated |
| COL6A5 | 4,041841612 | 0,001328854 | 0,037629821 | Upregulated |

| | | | | |
|---|---|---|---|---|
| COLEC12 | 2,511276312 | 0,000674837 | 0,024495734 | Upregulated |
| COPS8P2 | 3,21920332 | 0,001817155 | 0,045765925 | Upregulated |
| CPA4 | 5,663519205 | 0,000371832 | 0,016558595 | Upregulated |
| CR2 | 2,957989537 | 0,001661541 | 0,043317107 | Upregulated |
| CREBRF | 1,020063288 | 0,00153073 | 0,041381121 | Upregulated |
| CRH | 6,758974875 | 0,000904729 | 0,029187932 | Upregulated |
| CRYAB | 2,75297672 | 0,000392636 | 0,017058291 | Upregulated |
| CSMD3 | 4,960193208 | 0,002013222 | 0,048616174 | Upregulated |
| CSNK1A1P1 | 2,733768609 | 0,000272544 | 0,013477946 | Upregulated |
| CSPG5 | 2,394438312 | 0,000890417 | 0,028899964 | Upregulated |
| CST2 | 5,846643178 | 3,41781E-05 | 0,003014884 | Upregulated |
| CSTB | 2,14164812 | 0,000394333 | 0,017058291 | Upregulated |
| CTTN | 1,97942942 | 0,00082408 | 0,027466779 | Upregulated |
| CYFIP1 | 2,550906792 | 5,27533E-05 | 0,004173212 | Upregulated |
| CYP27A1 | 2,736593026 | 0,000753614 | 0,026271734 | Upregulated |
| CYP27B1 | 1,959393664 | 0,000109397 | 0,007072817 | Upregulated |
| DBH | 3,34964362 | 5,46218E-05 | 0,00425825 | Upregulated |
| DBH-AS1 | 4,088262872 | 9,40474E-06 | 0,001187468 | Upregulated |
| DCLK3 | 2,649342468 | 0,000284159 | 0,013905031 | Upregulated |
| DCSTAMP | 5,54996218 | 9,7944E-06 | 0,001212886 | Upregulated |
| DEPDC7 | 3,105620209 | 0,000255864 | 0,012912309 | Upregulated |
| DHRS2 | 3,9974905 | 0,001731608 | 0,044424356 | Upregulated |
| DIP2C | 3,409562679 | 6,76749E-05 | 0,004997543 | Upregulated |
| DIRAS3 | 3,230137334 | 0,001344127 | 0,037730482 | Upregulated |
| DNAJB1 | 3,355708018 | 3,55295E-05 | 0,003100115 | Upregulated |
| DNAJB5 | 2,009043661 | 5,13672E-05 | 0,00409128 | Upregulated |
| DNAJC5B | 2,390948377 | 0,001596386 | 0,042164749 | Upregulated |
| DOCK1 | 2,630073356 | 0,000505081 | 0,020027202 | Upregulated |
| DPF1 | 2,250469549 | 0,000270529 | 0,01340036 | Upregulated |
| DPYS | 4,508335053 | 7,79242E-06 | 0,001020416 | Upregulated |
| DPYSL4 | 3,500224952 | 1,48539E-06 | 0,000289849 | Upregulated |
| DTX1 | 4,145187608 | 1,0272E-06 | 0,000221964 | Upregulated |
| DUOX2 | 4,181338363 | 0,001842912 | 0,046066437 | Upregulated |
| EEF1GP4 | 5,184809715 | 0,000199889 | 0,010797647 | Upregulated |
| EGLN3 | 2,15143604 | 0,002070965 | 0,04946502 | Upregulated |
| EGR2 | 2,711987452 | 0,000183892 | 0,010155693 | Upregulated |
| EGR3 | 2,867673171 | 2,4731E-05 | 0,002376465 | Upregulated |
| EGR4 | 3,470243243 | 0,000222401 | 0,011634685 | Upregulated |
| EHF | 3,60929176 | 0,001399404 | 0,038741709 | Upregulated |
| ELAVL3 | 2,089796736 | 0,001281388 | 0,036896777 | Upregulated |
| ELF3 | 3,764522724 | 7,47749E-05 | 0,005312775 | Upregulated |
| ELL2P1 | 2,726986983 | 6,88987E-05 | 0,005053146 | Upregulated |
| EN1 | 5,239975691 | 4,54073E-05 | 0,003732376 | Upregulated |
| ENPP5 | 3,399156441 | 0,001682291 | 0,043646025 | Upregulated |
| ENPP7P4 | 2,238922731 | 0,000289945 | 0,014080487 | Upregulated |
| EPB41L5 | 1,76070842 | 1,43897E-05 | 0,001570527 | Upregulated |

| | | | | |
|---|---|---|---|---|
| EPHX3 | 3,819202191 | 4,78854E-09 | 2,71064E-06 | Upregulated |
| ESRP2 | 1,492473212 | 0,000158793 | 0,009346704 | Upregulated |
| EVC | 2,918571648 | 0,001011552 | 0,031346442 | Upregulated |
| EVC2 | 2,704714264 | 0,001285824 | 0,036898106 | Upregulated |
| FAM153A | 2,564990878 | 0,001174081 | 0,034904775 | Upregulated |
| FAM153CP | 2,732861171 | 6,42869E-06 | 0,000903862 | Upregulated |
| FAM155A-IT1 | 5,533314213 | 0,000131487 | 0,00800281 | Upregulated |
| FAM83A | 2,004082456 | 0,001413701 | 0,039078186 | Upregulated |
| FAT3 | 6,543170696 | 2,90041E-06 | 0,000510255 | Upregulated |
| FAXC | 4,844481669 | 0,000196067 | 0,01069961 | Upregulated |
| FBP1 | 3,43717405 | 0,001500032 | 0,040756557 | Upregulated |
| FBXO10 | 1,227998317 | 0,000164241 | 0,009580262 | Upregulated |
| FCGR2B | 3,103001456 | 0,001855147 | 0,046066437 | Upregulated |
| FCRLB | 3,273534884 | 8,22733E-05 | 0,005687879 | Upregulated |
| FFAR3 | 3,285771832 | 0,0015468 | 0,04155878 | Upregulated |
| FGFR2 | 3,275625505 | 0,001793646 | 0,045474512 | Upregulated |
| FGR | 2,430431871 | 0,00050735 | 0,020080929 | Upregulated |
| FHAD1 | 3,566702583 | 3,39569E-05 | 0,003003596 | Upregulated |
| FHL2 | 1,996738932 | 0,000109868 | 0,007088996 | Upregulated |
| FMN1 | 2,345564082 | 0,001464986 | 0,040074896 | Upregulated |
| FNDC7 | 2,813279515 | 6,41218E-05 | 0,004816271 | Upregulated |
| FOSB | 2,980884912 | 0,000590733 | 0,022563865 | Upregulated |
| FST | 3,652170715 | 3,9874E-05 | 0,003369614 | Upregulated |
| FYB2 | 7,122766692 | 1,40375E-05 | 0,001542549 | Upregulated |
| FYN | 1,325352401 | 4,56756E-05 | 0,003742032 | Upregulated |
| FZD3 | 3,628543293 | 3,55016E-06 | 0,000580834 | Upregulated |
| GACAT2 | 4,600414991 | 1,22236E-05 | 0,001395615 | Upregulated |
| GADD45G | 3,595323754 | 8,0819E-06 | 0,001042118 | Upregulated |
| GBX1 | 3,092721751 | 0,000645715 | 0,023827449 | Upregulated |
| GCM1 | 2,800506554 | 0,000303333 | 0,014468768 | Upregulated |
| GCSAM | 2,542708869 | 0,000395095 | 0,017058291 | Upregulated |
| GFOD1 | 2,424702437 | 6,20826E-09 | 3,27684E-06 | Upregulated |
| GFOD1-AS1 | 2,17513775 | 8,67719E-05 | 0,005894084 | Upregulated |
| GGT7 | 1,916130391 | 0,000267678 | 0,01334124 | Upregulated |
| GHRH | 3,864662359 | 0,000122008 | 0,007627776 | Upregulated |
| GJA1 | 3,256572086 | 0,000673675 | 0,024495734 | Upregulated |
| GJB7 | 1,825814403 | 3,34664E-05 | 0,002968372 | Upregulated |
| GLCCI1 | 1,306314494 | 0,001295587 | 0,037013328 | Upregulated |
| GOLGA6D | 2,321623881 | 0,000810739 | 0,027362029 | Upregulated |
| GOLM1 | 1,994786759 | 0,000677466 | 0,024535852 | Upregulated |
| GPC1 | 2,954748588 | 4,97717E-05 | 0,00400625 | Upregulated |
| GPHA2 | 1,875581682 | 0,000299871 | 0,01438583 | Upregulated |
| GPNMB | 2,228834906 | 0,001367269 | 0,038180363 | Upregulated |
| GPR153 | 1,968625026 | 0,000197557 | 0,010744503 | Upregulated |
| GPR157 | 3,309678671 | 1,55922E-09 | 1,16749E-06 | Upregulated |
| GPRC5A | 3,605447374 | 0,001849379 | 0,046066437 | Upregulated |

| | | | | |
|---|---|---|---|---|
| GPRIN1 | 2,23453123 | 0,000416688 | 0,017746157 | Upregulated |
| GRM5 | 5,838087303 | 0,00043594 | 0,018323695 | Upregulated |
| GTF2IP7 | 5,100927611 | 0,000707535 | 0,02517181 | Upregulated |
| GULOP | 2,923554674 | 0,001182545 | 0,035091621 | Upregulated |
| HBEGF | 2,399086337 | 0,001339209 | 0,037690993 | Upregulated |
| HCAR2 | 2,756933457 | 0,000292316 | 0,0141317 | Upregulated |
| HES4 | 4,630746417 | 1,50757E-06 | 0,000292404 | Upregulated |
| HES5 | 7,197262675 | 2,03393E-10 | 2,11247E-07 | Upregulated |
| HEY1 | 5,402580128 | 3,6724E-10 | 3,37829E-07 | Upregulated |
| HIPK2 | 2,213089033 | 1,08311E-06 | 0,000229427 | Upregulated |
| HLA-DQA1 | 2,035101824 | 0,000951277 | 0,030057169 | Upregulated |
| HMGA2 | 7,557798271 | 2,45301E-05 | 0,002376465 | Upregulated |
| HMGA2-AS1 | 7,897627375 | 3,50423E-09 | 2,24786E-06 | Upregulated |
| HNRNPA1P71 | 3,88894494 | 7,00558E-07 | 0,000165852 | Upregulated |
| HOMER1 | 2,792238871 | 0,000863688 | 0,028404652 | Upregulated |
| HPYR1 | 4,68934982 | 0,001818484 | 0,045765925 | Upregulated |
| HSPA1A | 3,385632719 | 0,000204085 | 0,010915154 | Upregulated |
| HSPA1B | 3,612648571 | 7,4436E-05 | 0,005302246 | Upregulated |
| HSPA6 | 4,858695236 | 5,66845E-05 | 0,004387187 | Upregulated |
| HSPA7 | 3,224512829 | 0,000948048 | 0,029984584 | Upregulated |
| HSPD1P10 | 1,861429636 | 0,00069297 | 0,024901284 | Upregulated |
| ICAM5 | 3,975204562 | 2,8512E-07 | 7,7143E-05 | Upregulated |
| IGF1R | 1,47898704 | 0,001276787 | 0,036835768 | Upregulated |
| IGFBPL1 | 7,564436804 | 2,53037E-07 | 6,90427E-05 | Upregulated |
| IKZF3 | 2,596200688 | 1,9904E-06 | 0,000372587 | Upregulated |
| IL13 | 4,763684379 | 1,27299E-06 | 0,000264428 | Upregulated |
| IL13RA2 | 4,764863608 | 0,001957435 | 0,047918621 | Upregulated |
| IL18BP | 1,586793845 | 0,000171049 | 0,009725125 | Upregulated |
| IL1RN | 3,900680775 | 0,000941302 | 0,029936712 | Upregulated |
| IL20RA | 6,004109517 | 0,000308606 | 0,014655154 | Upregulated |
| IL4I1 | 3,316911871 | 0,000145899 | 0,008731444 | Upregulated |
| IMPDH1P5 | 2,381944652 | 0,000413004 | 0,017613018 | Upregulated |
| INSYN2A | 6,101368402 | 1,96434E-09 | 1,4374E-06 | Upregulated |
| ISL2 | 3,595659908 | 0,00033114 | 0,015308898 | Upregulated |
| ISM1-AS1 | 2,519264164 | 0,000619002 | 0,023217803 | Upregulated |
| ITGB8 | 4,454342033 | 2,03614E-05 | 0,002035947 | Upregulated |
| JAKMIP3 | 3,505045407 | 3,10964E-09 | 2,08586E-06 | Upregulated |
| KCNA1 | 5,160139617 | 3,82528E-05 | 0,003275601 | Upregulated |
| KCNC1 | 1,080762415 | 0,000926645 | 0,029657236 | Upregulated |
| KCNH5 | 6,31654651 | 0,000418434 | 0,017796986 | Upregulated |
| KCNJ2 | 3,721735333 | 3,23596E-07 | 8,6106E-05 | Upregulated |
| KCNJ2-AS1 | 3,474057261 | 7,35889E-07 | 0,000171691 | Upregulated |
| KDELC1P1 | 1,594545655 | 0,000494321 | 0,019738004 | Upregulated |
| KDM6B | 1,240174964 | 0,001258598 | 0,036507279 | Upregulated |
| KDM7A | 1,655558101 | 6,06486E-06 | 0,000875263 | Upregulated |
| KIF21A | 3,46204578 | 0,000342845 | 0,015635388 | Upregulated |

| | | | | |
|---|---|---|---|---|
| KIF3A | 2,233724743 | 3,84646E-05 | 0,003284997 | Upregulated |
| KIF5C | 2,985100585 | 0,000109336 | 0,007072817 | Upregulated |
| KIR2DL1 | 4,426994607 | 0,001228586 | 0,035877535 | Upregulated |
| KIR2DL3 | 3,511331286 | 0,001488981 | 0,040557217 | Upregulated |
| KLF4 | 2,729586068 | 0,000389179 | 0,016936751 | Upregulated |
| KLK5 | 6,644170386 | 0,000365305 | 0,016335739 | Upregulated |
| KRT12 | 4,320233127 | 2,82066E-05 | 0,002587369 | Upregulated |
| KRT18P15 | 2,890892917 | 1,78977E-06 | 0,000340978 | Upregulated |
| KRT7 | 4,2657477 | 0,0007666 | 0,02648306 | Upregulated |
| KRT8P34 | 2,680091993 | 3,78369E-05 | 0,003266047 | Upregulated |
| KRT8P42 | 5,548921388 | 6,81964E-05 | 0,005024533 | Upregulated |
| KRTAP5-AS1 | 5,041353346 | 1,36392E-05 | 0,001524043 | Upregulated |
| L1TD1 | 3,933384208 | 0,000198736 | 0,01076156 | Upregulated |
| LAMA1 | 2,544339027 | 0,001528245 | 0,041348661 | Upregulated |
| LAMB3 | 3,782743157 | 3,04458E-06 | 0,000520772 | Upregulated |
| LDHAP2 | 2,131124211 | 0,001073545 | 0,032794061 | Upregulated |
| LDLRAD4-AS1 | 1,594300445 | 0,000960893 | 0,030212779 | Upregulated |
| LGALSL-DT | 2,296412671 | 0,000356015 | 0,016099157 | Upregulated |
| LILRA6 | 2,505025591 | 0,001963061 | 0,047918621 | Upregulated |
| LINC00161 | 2,7530752 | 0,000233468 | 0,012065782 | Upregulated |
| LINC00377 | 3,442377399 | 1,88783E-06 | 0,000355453 | Upregulated |
| LINC00390 | 1,264881461 | 0,001038241 | 0,03192766 | Upregulated |
| LINC00461 | 6,506950835 | 0,001632133 | 0,042758156 | Upregulated |
| LINC00520 | 4,268076566 | 4,24615E-09 | 2,4857E-06 | Upregulated |
| LINC00607 | 3,028380312 | 0,000116597 | 0,007405794 | Upregulated |
| LINC00689 | 4,321304994 | 2,45792E-05 | 0,002376465 | Upregulated |
| LINC00881 | 1,389625539 | 0,000944675 | 0,029936712 | Upregulated |
| LINC01010 | 3,945797763 | 0,00038213 | 0,016789032 | Upregulated |
| LINC01031 | 1,723845032 | 0,000174598 | 0,009826041 | Upregulated |
| LINC01033 | 2,648825214 | 0,00100942 | 0,031310494 | Upregulated |
| LINC01127 | 3,802976725 | 0,001680096 | 0,043624243 | Upregulated |
| LINC01133 | 3,597990094 | 0,000649378 | 0,023894889 | Upregulated |
| LINC01181 | 3,302358876 | 0,001206469 | 0,035571166 | Upregulated |
| LINC01220 | 2,308301156 | 0,000699526 | 0,024969672 | Upregulated |
| LINC01281 | 3,327150997 | 7,18386E-05 | 0,005197724 | Upregulated |
| LINC01307 | 2,8978352 | 0,000692916 | 0,024901284 | Upregulated |
| LINC01348 | 3,038479923 | 0,001388161 | 0,038496645 | Upregulated |
| LINC01353 | 2,336658557 | 0,000730564 | 0,025707061 | Upregulated |
| LINC01485 | 5,62996734 | 4,88796E-05 | 0,003954216 | Upregulated |
| LINC01607 | 4,195739724 | 2,56475E-05 | 0,002428741 | Upregulated |
| LINC01709 | 6,263584231 | 4,8499E-07 | 0,000121048 | Upregulated |
| LINC01727 | 3,858811781 | 0,000196728 | 0,010717498 | Upregulated |
| LINC01956 | 8,980860796 | 1,51542E-07 | 4,69154E-05 | Upregulated |
| LINC02154 | 5,524699977 | 3,55388E-06 | 0,000580834 | Upregulated |
| LINC02196 | 5,453559791 | 4,0559E-05 | 0,003418526 | Upregulated |
| LINC02227 | 7,525204347 | 5,14319E-08 | 1,88176E-05 | Upregulated |

| | | | | |
|---|---|---|---|---|
| LINC02331 | 8,879522669 | 4,20613E-20 | 1,35425E-15 | Upregulated |
| LINC02404 | 2,719805719 | 0,000454215 | 0,018798733 | Upregulated |
| LINC02454 | 9,312071554 | 0,001426258 | 0,039233172 | Upregulated |
| LINC02613 | 3,13043812 | 0,001996621 | 0,048415124 | Upregulated |
| LINC02614 | 2,93185839 | 5,13825E-06 | 0,000773067 | Upregulated |
| LINC02649 | 1,880330649 | 0,000819967 | 0,027442834 | Upregulated |
| LINC02656 | 2,130093464 | 0,000119899 | 0,007525115 | Upregulated |
| LINC02680 | 1,807213357 | 0,000292209 | 0,0141317 | Upregulated |
| LINGO2 | 5,721524488 | 0,001397616 | 0,038725511 | Upregulated |
| LIPC | 2,091539275 | 7,53531E-05 | 0,005324515 | Upregulated |
| LIPC-AS1 | 2,415743645 | 0,00037136 | 0,016558595 | Upregulated |
| LNCSRLR | 2,418243554 | 0,001571465 | 0,04188448 | Upregulated |
| LOX | 2,739815666 | 0,001278557 | 0,036853797 | Upregulated |
| LPO | 3,922401552 | 0,000429612 | 0,018128734 | Upregulated |
| LRGUK | 2,035924324 | 0,000166477 | 0,009619318 | Upregulated |
| LRMDA | 2,137284891 | 0,000481783 | 0,019414223 | Upregulated |
| LRMP | 1,507744529 | 0,000893655 | 0,02894849 | Upregulated |
| LRRTM3 | 7,467692231 | 7,59965E-09 | 3,94655E-06 | Upregulated |
| LRRTM4 | 5,789055869 | 0,001289043 | 0,036919948 | Upregulated |
| LYPD3 | 2,91773322 | 1,79548E-05 | 0,001852858 | Upregulated |
| LYST | 2,63998774 | 0,000495335 | 0,019738004 | Upregulated |
| MAFA | 4,28914913 | 0,001636789 | 0,042845286 | Upregulated |
| MAP6D1 | 4,274118696 | 3,44216E-07 | 9,0842E-05 | Upregulated |
| MCC | 2,015462871 | 0,000486943 | 0,019548777 | Upregulated |
| MCTP1 | 1,755445795 | 0,000756726 | 0,026273172 | Upregulated |
| MCTP1-AS1 | 2,201418765 | 0,000695392 | 0,024933985 | Upregulated |
| MDGA2 | 4,874364674 | 0,001588856 | 0,042138717 | Upregulated |
| MGARP | 3,420341012 | 0,000325502 | 0,015166714 | Upregulated |
| MIAT | 1,794696165 | 0,001152209 | 0,03439069 | Upregulated |
| MIR1252 | 4,410498907 | 0,000653046 | 0,024002409 | Upregulated |
| MIR1267 | 4,930585687 | 0,000210579 | 0,011161078 | Upregulated |
| MIR138-1 | 4,086155586 | 0,001473904 | 0,040263333 | Upregulated |
| MIR210 | 3,635725503 | 0,001789043 | 0,045427295 | Upregulated |
| MIR4322 | 3,77241133 | 0,000336768 | 0,015467777 | Upregulated |
| MIR4425 | 3,970689139 | 0,000755946 | 0,026273172 | Upregulated |
| MIR4427 | 3,308233725 | 0,000304794 | 0,014516938 | Upregulated |
| MIR4714 | 2,069730885 | 0,0020632 | 0,049316141 | Upregulated |
| MIR5580 | 6,028570168 | 0,000468246 | 0,019035496 | Upregulated |
| MIR635 | 1,43238251 | 0,001926877 | 0,047394696 | Upregulated |
| MIR7845 | 2,432423193 | 0,000656042 | 0,024085029 | Upregulated |
| MITF | 3,55719486 | 8,06352E-06 | 0,001042118 | Upregulated |
| MKLN1-AS | 1,251831132 | 5,69448E-06 | 0,000825879 | Upregulated |
| MMP19 | 2,879306778 | 0,00017202 | 0,0097338 | Upregulated |
| MNS1 | 3,550374763 | 1,90626E-05 | 0,001934009 | Upregulated |
| MTCL1 | 2,977248 | 1,30232E-05 | 0,001461 | Upregulated |
| MTCYBP23 | 4,162173337 | 0,000171565 | 0,009725125 | Upregulated |

| | | | | |
|---|---|---|---|---|
| MTND5P32 | 3,607128282 | 0,001001593 | 0,031157773 | Upregulated |
| MTSS1 | 2,795362823 | 0,000285103 | 0,013908257 | Upregulated |
| MXI1 | 2,044549438 | 0,000173963 | 0,009809279 | Upregulated |
| MYBPH | 4,09684557 | 0,001322839 | 0,037591735 | Upregulated |
| MYOG | 3,511438194 | 0,000205608 | 0,010978366 | Upregulated |
| NAB2 | 2,000028578 | 1,51938E-05 | 0,001630646 | Upregulated |
| NCS1 | 2,265230418 | 0,000887554 | 0,028854716 | Upregulated |
| NEBL | 3,445090417 | 0,000625487 | 0,023348352 | Upregulated |
| NECTIN4 | 4,741119422 | 1,14689E-05 | 0,001347677 | Upregulated |
| NIM1K | 1,776108473 | 7,33867E-08 | 2,48719E-05 | Upregulated |
| NINJ1 | 1,70826152 | 0,001265877 | 0,03665237 | Upregulated |
| NIPA1 | 1,238786319 | 0,001273749 | 0,036781083 | Upregulated |
| NKD2 | 2,155058754 | 0,000813031 | 0,027410653 | Upregulated |
| NKG7 | 2,652553909 | 0,000838973 | 0,027705031 | Upregulated |
| NKX2-5 | 9,146881417 | 3,93269E-12 | 5,27586E-09 | Upregulated |
| NPAS2 | 3,138672889 | 6,87892E-05 | 0,005053146 | Upregulated |
| NPAS4 | 5,364259323 | 0,000272933 | 0,013477946 | Upregulated |
| NPM1P30 | 5,191522496 | 0,000903601 | 0,029180787 | Upregulated |
| NR1D1 | 2,159421398 | 0,000278044 | 0,013688335 | Upregulated |
| NR4A1 | 2,369311237 | 0,000300884 | 0,014394609 | Upregulated |
| NR4A3 | 2,210600044 | 0,001519077 | 0,041204482 | Upregulated |
| NRARP | 4,821354418 | 5,47323E-10 | 4,76275E-07 | Upregulated |
| NUAK1 | 3,030210995 | 0,001265839 | 0,03665237 | Upregulated |
| NUPR1 | 2,841533326 | 7,25453E-06 | 0,000985545 | Upregulated |
| OLFM3 | 5,26320781 | 0,000695429 | 0,024933985 | Upregulated |
| OLR1 | 3,500550547 | 0,001144798 | 0,034223838 | Upregulated |
| OPRD1 | 2,451151702 | 7,26176E-05 | 0,005210074 | Upregulated |
| OR7D2 | 3,749276015 | 0,001342328 | 0,037730482 | Upregulated |
| OTOG | 5,933760392 | 0,001490176 | 0,040557217 | Upregulated |
| OTOR | 4,781838071 | 0,000160642 | 0,009421096 | Upregulated |
| P2RY11 | 2,362644222 | 0,000122874 | 0,007666998 | Upregulated |
| P4HA2 | 2,279542865 | 0,000727316 | 0,025627505 | Upregulated |
| PADI6 | 3,9327943 | 0,001291172 | 0,036919948 | Upregulated |
| PAFAH2 | 1,658072502 | 0,000221302 | 0,011623595 | Upregulated |
| PAPPA | 4,780785258 | 6,73737E-08 | 2,35786E-05 | Upregulated |
| PAPPA-AS2 | 6,015983918 | 2,43834E-06 | 0,000438589 | Upregulated |
| PARD6G-AS1 | 1,918127272 | 3,08902E-05 | 0,002801608 | Upregulated |
| PCA3 | 4,986114443 | 7,8266E-07 | 0,000179519 | Upregulated |
| PCAT14 | 4,659628776 | 7,45603E-06 | 0,000996106 | Upregulated |
| PCBP3 | 2,071712479 | 0,000713673 | 0,025306327 | Upregulated |
| PCDH9 | 3,672687277 | 1,61165E-05 | 0,001712549 | Upregulated |
| PCDH9-AS2 | 4,790815029 | 6,6704E-06 | 0,000921746 | Upregulated |
| PCDH9-AS3 | 4,231331641 | 0,00015146 | 0,008963958 | Upregulated |
| PCDH9-AS4 | 3,601753699 | 0,000165971 | 0,009619318 | Upregulated |
| PDE10A | 2,776463205 | 0,001173891 | 0,034904775 | Upregulated |
| PDE3A | 3,796749771 | 0,000395444 | 0,017058291 | Upregulated |

| | | | | |
|---|---|---|---|---|
| PDE4A | 2,203494976 | 0,000451983 | 0,018777401 | Upregulated |
| PDE4D | 2,356278071 | 0,000423468 | 0,017939997 | Upregulated |
| PDIA2 | 2,76814225 | 0,001413985 | 0,039078186 | Upregulated |
| PERM1 | 4,475792936 | 5,33996E-08 | 1,9318E-05 | Upregulated |
| PERP | 3,434505251 | 0,000399991 | 0,01719429 | Upregulated |
| PFKFB3 | 2,206206952 | 0,000182822 | 0,01011642 | Upregulated |
| PGBD5 | 3,479808124 | 7,33072E-05 | 0,005245045 | Upregulated |
| PGPEP1L | 3,29724729 | 0,00016642 | 0,009619318 | Upregulated |
| PHACTR3 | 4,511670406 | 0,000630357 | 0,02346311 | Upregulated |
| PHF21B | 3,501911742 | 0,002099358 | 0,049998732 | Upregulated |
| PHKA2-AS1 | 1,745238623 | 4,2592E-07 | 0,000108836 | Upregulated |
| PHYHIP | 2,83658928 | 0,000802946 | 0,027270526 | Upregulated |
| PITPNC1 | 1,273324301 | 0,0001401 | 0,008463067 | Upregulated |
| PLA2G2C | 3,980663389 | 2,05289E-08 | 8,93202E-06 | Upregulated |
| PLA2G2F | 5,807575534 | 3,56061E-09 | 2,24786E-06 | Upregulated |
| PLAT | 4,285281462 | 6,69395E-05 | 0,004966017 | Upregulated |
| PLCD3 | 1,714446164 | 0,002047046 | 0,04907576 | Upregulated |
| PLCL1 | 2,330906798 | 0,000868417 | 0,028493943 | Upregulated |
| PLEKHN1 | 2,630515149 | 0,00076574 | 0,02648306 | Upregulated |
| PLEKHS1 | 2,991589655 | 0,001938434 | 0,047569943 | Upregulated |
| PLOD2 | 2,728576417 | 0,001580106 | 0,042045183 | Upregulated |
| PLXNA2 | 3,187299203 | 3,81579E-05 | 0,003275601 | Upregulated |
| POMT1 | 1,347330656 | 1,83989E-05 | 0,001882419 | Upregulated |
| PPAN-P2RY11 | 3,539418937 | 0,001669465 | 0,043479495 | Upregulated |
| PPEF1 | 2,857558766 | 1,44847E-05 | 0,001575555 | Upregulated |
| PPL | 2,798715351 | 0,000448327 | 0,018697901 | Upregulated |
| PPP1R12BP1 | 10,52286391 | 1,23298E-06 | 0,000259466 | Upregulated |
| PPP1R12BP2 | 9,660033906 | 0,00050155 | 0,019945235 | Upregulated |
| PPP1R16B | 1,718851593 | 1,99073E-07 | 5,77437E-05 | Upregulated |
| PPP1R3C | 2,924038925 | 2,39389E-05 | 0,002335643 | Upregulated |
| PRKAG3 | 3,851531322 | 0,00083521 | 0,027637461 | Upregulated |
| PRKAR2B | 2,019904913 | 0,001444201 | 0,039573552 | Upregulated |
| PRKCZ-AS1 | 1,432072272 | 0,001081056 | 0,032992198 | Upregulated |
| PRLR | 3,177209761 | 0,000925072 | 0,029657236 | Upregulated |
| PRR15 | 3,978616494 | 6,24326E-05 | 0,004757566 | Upregulated |
| PRUNE2 | 2,602756906 | 0,000323751 | 0,015155739 | Upregulated |
| PSMC1P7 | 6,093225485 | 0,000995347 | 0,031023422 | Upregulated |
| PTGIS | 3,641279574 | 0,000733178 | 0,025742772 | Upregulated |
| PUDP | 1,686376031 | 0,000646586 | 0,023827449 | Upregulated |
| PWWP2AP1 | 2,817078526 | 0,000340056 | 0,015574368 | Upregulated |
| PYDC1 | 4,108147535 | 0,001091627 | 0,033123107 | Upregulated |
| RAPGEF1 | 1,166053779 | 0,000609122 | 0,022964759 | Upregulated |
| RAPGEF5 | 1,924763945 | 0,00165281 | 0,043159379 | Upregulated |
| RASAL1 | 2,679496577 | 0,002090042 | 0,04988368 | Upregulated |
| RASGRF2 | 1,978549616 | 0,000393536 | 0,017058291 | Upregulated |
| RASSF8 | 2,202264516 | 0,001961715 | 0,047918621 | Upregulated |

| | | | | |
|---|---|---|---|---|
| RASSF8-AS1 | 2,749878116 | 0,000251952 | 0,012734847 | Upregulated |
| RBM43P1 | 3,877088671 | 0,001209714 | 0,035571166 | Upregulated |
| RD3L | 4,72701411 | 0,001134966 | 0,034042775 | Upregulated |
| REM2 | 2,008158842 | 0,000555916 | 0,021538914 | Upregulated |
| RF00002 | 2,169878538 | 0,000386791 | 0,016920515 | Upregulated |
| RF00591 | 3,422216411 | 0,001237793 | 0,036033665 | Upregulated |
| RGS3 | 2,265876067 | 3,4371E-05 | 0,003023614 | Upregulated |
| RGS4 | 3,485647647 | 6,83709E-06 | 0,000940743 | Upregulated |
| RHBDD1 | 1,591920556 | 0,00043972 | 0,018434474 | Upregulated |
| RHCG | 5,388343781 | 0,000431613 | 0,018189336 | Upregulated |
| RHPN2 | 2,655193192 | 0,000283869 | 0,013905031 | Upregulated |
| RIN1 | 1,509022743 | 0,000939087 | 0,029936431 | Upregulated |
| RN7SKP110 | 2,243331015 | 0,001498014 | 0,040736113 | Upregulated |
| RN7SL116P | 1,858387759 | 0,000534666 | 0,020916949 | Upregulated |
| RN7SL11P | 3,928674803 | 0,001677162 | 0,043583195 | Upregulated |
| RNA5-8SN1 | 1,983954294 | 0,000960368 | 0,030212779 | Upregulated |
| RNF128 | 4,818996337 | 0,000603344 | 0,022827114 | Upregulated |
| ROR2 | 2,768565106 | 0,000473032 | 0,019157484 | Upregulated |
| RPEP1 | 3,040767676 | 0,000336538 | 0,015467777 | Upregulated |
| RPL13AP3 | 3,135204015 | 0,001538426 | 0,041484673 | Upregulated |
| RPL7L1P12 | 2,582423891 | 0,001228426 | 0,035877535 | Upregulated |
| RPS23P6 | 2,158360891 | 0,000180852 | 0,010056817 | Upregulated |
| RPS29P7 | 4,124130663 | 0,001750418 | 0,044774975 | Upregulated |
| RPS3AP14 | 4,129809105 | 0,000166267 | 0,009619318 | Upregulated |
| RPS3AP46 | 3,316452451 | 0,001857112 | 0,046066437 | Upregulated |
| RPSAP52 | 7,675103706 | 0,000801561 | 0,027270526 | Upregulated |
| RRAD | 2,774233537 | 0,000186974 | 0,010290599 | Upregulated |
| RUNX3 | 2,268956346 | 0,000148374 | 0,008862924 | Upregulated |
| RYR3 | 4,348235629 | 3,72138E-11 | 4,60836E-08 | Upregulated |
| SALL1 | 9,212046486 | 4,07628E-09 | 2,4857E-06 | Upregulated |
| SARDH | 3,329329457 | 2,50298E-05 | 0,002391352 | Upregulated |
| SCG2 | 5,982732908 | 1,4315E-09 | 1,12415E-06 | Upregulated |
| SEMA3B | 3,129542479 | 0,001712044 | 0,044204245 | Upregulated |
| SEMA3E | 4,763996238 | 1,22789E-05 | 0,00139697 | Upregulated |
| SEMA7A | 2,247898136 | 2,50001E-06 | 0,000444712 | Upregulated |
| SERGEF | 1,355324355 | 0,001008461 | 0,031310494 | Upregulated |
| SERP2 | 3,019832526 | 1,17061E-05 | 0,001360655 | Upregulated |
| SERPINB10 | 3,276070714 | 0,002046438 | 0,04907576 | Upregulated |
| SERPINB12 | 8,191421348 | 5,37778E-06 | 0,000797918 | Upregulated |
| SERPINC1 | 1,848602107 | 6,52686E-05 | 0,004875759 | Upregulated |
| SERTAD4 | 5,589834156 | 5,65417E-09 | 3,09515E-06 | Upregulated |
| SERTAD4-AS1 | 6,002336871 | 6,07127E-09 | 3,25795E-06 | Upregulated |
| SGK1 | 3,051190431 | 3,14614E-05 | 0,002837429 | Upregulated |
| SGSH | 1,844107149 | 0,000858102 | 0,028278717 | Upregulated |
| SH2B2 | 1,800115123 | 8,24996E-05 | 0,005687879 | Upregulated |
| SHISA9 | 5,447814351 | 3,83037E-13 | 8,80904E-10 | Upregulated |

| | | | | |
|---|---|---|---|---|
| SHROOM2 | 3,894609705 | 7,24157E-06 | 0,000985545 | Upregulated |
| SIRPB1 | 2,13697178 | 0,000925736 | 0,029657236 | Upregulated |
| SIRPG | 3,032737369 | 0,000746433 | 0,026151141 | Upregulated |
| SLC12A8 | 2,463435925 | 0,000170143 | 0,009712958 | Upregulated |
| SLC16A12 | 4,169001377 | 3,00903E-06 | 0,000520772 | Upregulated |
| SLC16A6 | 2,51998834 | 0,00044755 | 0,018697901 | Upregulated |
| SLC26A11 | 1,870713568 | 0,001850602 | 0,046066437 | Upregulated |
| SLC28A3 | 4,07350036 | 0,00045872 | 0,018862582 | Upregulated |
| SLC34A1 | 3,480638885 | 0,001334925 | 0,037669225 | Upregulated |
| SLC35F4 | 2,676351714 | 5,90274E-05 | 0,004535811 | Upregulated |
| SLC36A3 | 2,49155224 | 1,53846E-05 | 0,001645637 | Upregulated |
| SLC6A8 | 2,840393106 | 0,000756092 | 0,026273172 | Upregulated |
| SLC9A8 | 1,088609004 | 3,46785E-05 | 0,003042352 | Upregulated |
| SMC1B | 3,575840928 | 4,97228E-06 | 0,000760379 | Upregulated |
| SMG1P1 | 1,430554809 | 8,95681E-05 | 0,006020508 | Upregulated |
| SMG1P3 | 1,415035154 | 0,000133629 | 0,008117822 | Upregulated |
| SMG1P4 | 1,498233981 | 0,000107231 | 0,006988872 | Upregulated |
| SMG1P6 | 1,181507286 | 0,001767383 | 0,045090672 | Upregulated |
| SMIM10L2A | 2,169826692 | 0,001782107 | 0,045322666 | Upregulated |
| SNX29 | 1,373425573 | 0,00047502 | 0,019213851 | Upregulated |
| SPAG4 | 1,865108866 | 0,001817707 | 0,045765925 | Upregulated |
| SPINK1 | 7,460900869 | 0,000313268 | 0,014767625 | Upregulated |
| SPRY4-AS1 | 5,141003207 | 2,35217E-06 | 0,0004303 | Upregulated |
| SPSB1 | 1,31918223 | 0,001743435 | 0,044656613 | Upregulated |
| SPTSSB | 5,124942487 | 4,39002E-06 | 0,000686142 | Upregulated |
| SRGAP2D | 1,268083693 | 0,000457188 | 0,0188566 | Upregulated |
| ST14 | 3,233444492 | 0,00109172 | 0,033123107 | Upregulated |
| STAC2 | 3,397239128 | 0,001640171 | 0,042898941 | Upregulated |
| STC1 | 2,537286986 | 0,00197109 | 0,048041773 | Upregulated |
| SYNGR3 | 1,61666133 | 0,002042941 | 0,04907576 | Upregulated |
| SYT14 | 5,679190813 | 0,000327295 | 0,015206237 | Upregulated |
| SYTL2 | 1,869098493 | 0,000502394 | 0,019945235 | Upregulated |
| TBL1X | 1,72867997 | 0,000173385 | 0,009793826 | Upregulated |
| TBR1 | 2,957233569 | 0,000605477 | 0,022854082 | Upregulated |
| TBX21 | 2,712924331 | 0,000435786 | 0,018323695 | Upregulated |
| TCEAL2 | 5,020914999 | 0,000396298 | 0,017058291 | Upregulated |
| TDRD9 | 3,586046425 | 0,001030433 | 0,03183959 | Upregulated |
| TENM4 | 3,025208789 | 0,00113557 | 0,034042775 | Upregulated |
| TGFB2 | 2,315722825 | 0,001834909 | 0,046066437 | Upregulated |
| TGM1 | 3,171373867 | 0,00117685 | 0,034954834 | Upregulated |
| TIAM1 | 1,279739363 | 0,001091756 | 0,033123107 | Upregulated |
| TMEM108-AS1 | 3,94398492 | 0,000438312 | 0,018399394 | Upregulated |
| TMEM151B | 4,805370254 | 0,000565171 | 0,021740515 | Upregulated |
| TMEM155 | 3,48113891 | 0,0016862 | 0,043689153 | Upregulated |
| TMEM243 | 1,033876928 | 0,000309743 | 0,014665862 | Upregulated |
| TMEM45A | 3,047053595 | 0,00022445 | 0,011674671 | Upregulated |

| | | | | |
|---|---|---|---|---|
| TMEM51-AS1 | 3,188326675 | 8,60595E-05 | 0,005882924 | Upregulated |
| TNFAIP3 | 1,657050088 | 8,14961E-05 | 0,005672795 | Upregulated |
| TNNT2 | 4,752337471 | 0,001387913 | 0,038496645 | Upregulated |
| TRAJ32 | 3,239640089 | 0,001750837 | 0,044774975 | Upregulated |
| TRAV1-1 | 5,21429946 | 0,000901755 | 0,029150418 | Upregulated |
| TRAV13-1 | 3,260342837 | 0,000893711 | 0,02894849 | Upregulated |
| TRAV8-2 | 5,153423942 | 4,44176E-07 | 0,000111728 | Upregulated |
| TRAV8-3 | 3,06314113 | 0,000373574 | 0,0166132 | Upregulated |
| TRAV8-4 | 5,48511637 | 5,46398E-06 | 0,000806989 | Upregulated |
| TRBV21-1 | 5,496077754 | 7,77322E-05 | 0,005461043 | Upregulated |
| TRBV7-9 | 6,436044748 | 2,32386E-09 | 1,62655E-06 | Upregulated |
| TREM2 | 3,556757413 | 0,002099521 | 0,049998732 | Upregulated |
| TRPM5 | 2,376596249 | 0,000921054 | 0,029566482 | Upregulated |
| TRPM8 | 5,807878155 | 0,000141636 | 0,008523835 | Upregulated |
| TSPOAP1 | 2,641575138 | 4,91184E-08 | 1,83891E-05 | Upregulated |
| TWIST2 | 2,943295114 | 0,001594177 | 0,042140991 | Upregulated |
| U91319 | 7,075982283 | 3,23872E-05 | 0,002896588 | Upregulated |
| UBXN10 | 3,865268846 | 5,87068E-07 | 0,000141059 | Upregulated |
| UBXN10-AS1 | 4,529865831 | 5,55E-07 | 0,000134455 | Upregulated |
| ULBP1 | 6,692694367 | 4,75898E-08 | 1,80518E-05 | Upregulated |
| ULBP3 | 2,900643917 | 7,54101E-05 | 0,005324515 | Upregulated |
| UPK1B | 3,650222841 | 0,00162987 | 0,042733653 | Upregulated |
| VAV2 | 1,331865929 | 0,001192865 | 0,035332724 | Upregulated |
| VEGFA | 2,865236268 | 0,000261706 | 0,013165879 | Upregulated |
| VOPP1 | 1,700756404 | 2,48002E-05 | 0,002376465 | Upregulated |
| VPS37D | 1,880046425 | 8,79667E-05 | 0,005950133 | Upregulated |
| VSIG10L | 1,668793782 | 0,002039271 | 0,049035396 | Upregulated |
| VWC2 | 6,693200277 | 0,000103008 | 0,006810149 | Upregulated |
| WDR63 | 3,564121722 | 0,000342193 | 0,015627788 | Upregulated |
| WFDC2 | 4,477169977 | 0,001285613 | 0,036898106 | Upregulated |
| WNK2 | 4,181294718 | 0,000212121 | 0,011196171 | Upregulated |
| WWTR1 | 2,157433345 | 0,000954381 | 0,030096184 | Upregulated |
| XIRP1 | 3,840023216 | 0,00011931 | 0,007525115 | Upregulated |
| XRCC6P5 | 6,243195173 | 0,000325186 | 0,015166714 | Upregulated |
| YEATS2-AS1 | 1,333976676 | 0,000884546 | 0,028796499 | Upregulated |
| Z98257 | 3,670365406 | 9,49066E-06 | 0,001193635 | Upregulated |
| ZDHHC11B | 2,688755401 | 0,001034635 | 0,031847177 | Upregulated |
| ZFPM1 | 1,45160642 | 0,000248832 | 0,012616748 | Upregulated |
| ZNF365 | 5,439178297 | 0,000227577 | 0,011818234 | Upregulated |
| ZNF438 | 1,057775085 | 0,00048004 | 0,019368214 | Upregulated |
| ZNF474 | 1,82524203 | 0,001381647 | 0,038448471 | Upregulated |
| ZNF503 | 3,29938303 | 1,90844E-05 | 0,001934009 | Upregulated |
| ZNF503-AS1 | 3,987107389 | 0,000787783 | 0,026983244 | Upregulated |
| ZNF503-AS2 | 4,355547178 | 2,26556E-07 | 6,39861E-05 | Upregulated |
| ZNF821 | 1,682509298 | 0,000463206 | 0,018924703 | Upregulated |
| ABHD12B | -5,409960848 | 2,67459E-05 | 0,002484412 | Downregulated |

| | | | | |
|---|---|---|---|---|
| AC002407 | -3,862885434 | 0,000127921 | 0,007884236 | Downregulated |
| AC002454 | -6,723454226 | 1,27591E-12 | 2,16213E-09 | Downregulated |
| AC002464 | -6,617382737 | 3,47825E-09 | 2,24786E-06 | Downregulated |
| AC004522 | -3,072112607 | 4,34625E-05 | 0,003606607 | Downregulated |
| AC004540 | -1,96904748 | 0,001864171 | 0,046169774 | Downregulated |
| AC004817 | -4,785018927 | 5,01807E-06 | 0,000762108 | Downregulated |
| AC005336 | -8,143301663 | 7,93347E-19 | 8,51446E-15 | Downregulated |
| AC005551 | -4,745056975 | 7,79645E-06 | 0,001020416 | Downregulated |
| AC006130 | -2,598647231 | 0,001055792 | 0,032343791 | Downregulated |
| AC006504 | -1,108974659 | 0,000788645 | 0,026984079 | Downregulated |
| AC007100 | -5,251824325 | 3,4003E-06 | 0,000561432 | Downregulated |
| AC007132 | -4,650472149 | 9,87405E-06 | 0,001216677 | Downregulated |
| AC007179 | -4,808459373 | 1,13621E-05 | 0,001340021 | Downregulated |
| AC008539 | -8,055697077 | 3,52047E-07 | 9,21532E-05 | Downregulated |
| AC008591 | -5,752416606 | 0,000169354 | 0,009689896 | Downregulated |
| AC008878 | -1,398556466 | 0,001044731 | 0,032096565 | Downregulated |
| AC009152 | -2,928861858 | 0,001535369 | 0,041436946 | Downregulated |
| AC009560 | -2,691135013 | 0,000825985 | 0,027473377 | Downregulated |
| AC009570 | -3,680192298 | 7,16253E-05 | 0,005193962 | Downregulated |
| AC010615 | -1,538605311 | 0,001804312 | 0,045635071 | Downregulated |
| AC010719 | -4,527438758 | 2,79927E-05 | 0,002575088 | Downregulated |
| AC011509 | -3,246071505 | 5,35264E-06 | 0,000797865 | Downregulated |
| AC012499 | -5,131095997 | 2,66807E-05 | 0,002484412 | Downregulated |
| AC013564 | -3,165850657 | 0,001418127 | 0,039125467 | Downregulated |
| AC013565 | -3,255947996 | 0,000357069 | 0,016124198 | Downregulated |
| AC015802 | -1,325133288 | 0,001609634 | 0,042375635 | Downregulated |
| AC015908 | -3,938154296 | 0,000358305 | 0,016157353 | Downregulated |
| AC016735 | -7,152453915 | 7,48766E-06 | 0,000996199 | Downregulated |
| AC017100 | -4,631945361 | 2,73417E-14 | 7,336E-11 | Downregulated |
| AC018359 | -6,327517803 | 0,000808518 | 0,027328477 | Downregulated |
| AC018552 | -3,859214437 | 2,72383E-05 | 0,002512866 | Downregulated |
| AC019069 | -2,672535105 | 9,37212E-10 | 7,73729E-07 | Downregulated |
| AC019117 | -4,778989798 | 0,000191752 | 0,010489444 | Downregulated |
| AC020595 | -4,423339457 | 4,09073E-07 | 0,000105367 | Downregulated |
| AC022239 | -3,390857017 | 0,001143863 | 0,034223838 | Downregulated |
| AC023034 | -6,571819237 | 1,02675E-06 | 0,000221964 | Downregulated |
| AC024558 | -2,677256148 | 0,000219888 | 0,011568169 | Downregulated |
| AC024563 | -4,218070606 | 0,00156948 | 0,04188448 | Downregulated |
| AC025188 | -2,900122409 | 0,000833525 | 0,027610091 | Downregulated |
| AC025370 | -3,296174313 | 1,32123E-08 | 6,3492E-06 | Downregulated |
| AC026150 | -2,709456841 | 0,000775779 | 0,026714228 | Downregulated |
| AC037471 | -6,764381114 | 4,64039E-06 | 0,000721772 | Downregulated |
| AC055717 | -18,18949463 | 7,14461E-08 | 2,44718E-05 | Downregulated |
| AC060809 | -6,622204981 | 6,35803E-06 | 0,000897849 | Downregulated |
| AC064807 | -1,366912029 | 3,35618E-06 | 0,000559892 | Downregulated |
| AC068726 | -2,389456586 | 0,00059366 | 0,022646995 | Downregulated |

| | | | | |
|---|---|---|---|---|
| AC073415 | -2,201412879 | 0,000409681 | 0,017530623 | Downregulated |
| AC079313 | -2,09414987 | 5,57586E-06 | 0,000819753 | Downregulated |
| AC087277 | -3,537421557 | 0,000611543 | 0,023029047 | Downregulated |
| AC087498 | -4,795458169 | 0,000711447 | 0,025255189 | Downregulated |
| AC090541 | -2,25238782 | 0,001387618 | 0,038496645 | Downregulated |
| AC090579 | -3,078322094 | 0,000376575 | 0,016654642 | Downregulated |
| AC091078 | -6,333718213 | 6,41874E-08 | 2,27103E-05 | Downregulated |
| AC092079 | -2,707244989 | 0,001373853 | 0,038289537 | Downregulated |
| AC092142 | -3,25919777 | 0,000561447 | 0,021674957 | Downregulated |
| AC092640 | -3,381289823 | 0,001235907 | 0,036011321 | Downregulated |
| AC092646 | -21,40067373 | 4,3995E-11 | 5,08434E-08 | Downregulated |
| AC092933 | -2,950839708 | 0,000389189 | 0,016936751 | Downregulated |
| AC093390 | -9,089635309 | 1,42705E-08 | 6,65896E-06 | Downregulated |
| AC093418 | -4,987032251 | 2,25741E-07 | 6,39861E-05 | Downregulated |
| AC093484 | -1,330296726 | 0,000115845 | 0,007400506 | Downregulated |
| AC093770 | -2,775131963 | 0,000300254 | 0,01438583 | Downregulated |
| AC093890 | -3,570136456 | 2,26055E-05 | 0,002232606 | Downregulated |
| AC095052 | -6,396598145 | 3,67229E-06 | 0,000591184 | Downregulated |
| AC096589 | -4,977762068 | 0,001033037 | 0,031847177 | Downregulated |
| AC096677 | -2,006724078 | 3,62987E-08 | 1,47938E-05 | Downregulated |
| AC096713 | -2,951082991 | 0,000182864 | 0,01011642 | Downregulated |
| AC096751 | -4,088597654 | 0,000944502 | 0,029936712 | Downregulated |
| AC097713 | -2,836792306 | 0,00082929 | 0,027526444 | Downregulated |
| AC099494 | -3,132404341 | 5,09205E-05 | 0,004088496 | Downregulated |
| AC103923 | -3,250084464 | 1,49304E-07 | 4,66713E-05 | Downregulated |
| AC104041 | -3,503137759 | 0,000309437 | 0,014665862 | Downregulated |
| AC104083 | -3,230849808 | 0,00026535 | 0,013245678 | Downregulated |
| AC104335 | -1,576641335 | 0,001825694 | 0,045887481 | Downregulated |
| AC104411 | -2,7877862 | 0,000698897 | 0,024969672 | Downregulated |
| AC104785 | -2,330931631 | 8,32358E-05 | 0,005726372 | Downregulated |
| AC104791 | -2,344525294 | 1,81774E-05 | 0,001869834 | Downregulated |
| AC105383 | -9,28136569 | 2,68146E-09 | 1,83692E-06 | Downregulated |
| AC107204 | -4,874843171 | 2,94137E-10 | 2,95948E-07 | Downregulated |
| AC107223 | -2,071877935 | 0,001032107 | 0,031847177 | Downregulated |
| AC108047 | -1,273565403 | 0,000329327 | 0,015256589 | Downregulated |
| AC108704 | -2,067988423 | 1,1242E-05 | 0,001331847 | Downregulated |
| AC110023 | -7,398414154 | 9,04891E-08 | 2,94291E-05 | Downregulated |
| AC110285 | -3,180829123 | 2,34694E-05 | 0,002296795 | Downregulated |
| AC110751 | -5,185930252 | 0,001343927 | 0,037730482 | Downregulated |
| AC112482 | -7,263082446 | 4,49381E-08 | 1,74322E-05 | Downregulated |
| AC112693 | -5,609997066 | 7,52389E-06 | 0,000996901 | Downregulated |
| AC113414 | -2,635670082 | 0,001857137 | 0,046066437 | Downregulated |
| AC115622 | -4,932931457 | 0,000374217 | 0,016618831 | Downregulated |
| AC119403 | -1,496601173 | 8,42014E-05 | 0,005780452 | Downregulated |
| AC120042 | -3,314492208 | 0,001756274 | 0,044878385 | Downregulated |
| AC125603 | -3,15127718 | 0,00200696 | 0,048548525 | Downregulated |

| | | | | |
|---|---|---|---|---|
| AC126121 | -4,169533543 | 0,000420652 | 0,017844164 | Downregulated |
| AC132216 | -7,042294928 | 1,60046E-05 | 0,001706288 | Downregulated |
| AC132807 | -10,35261494 | 1,62362E-12 | 2,61379E-09 | Downregulated |
| AC134980 | -6,488687609 | 0,000402947 | 0,017275191 | Downregulated |
| AC136475 | -1,908442919 | 0,000339728 | 0,015574368 | Downregulated |
| AC139720 | -2,937764926 | 0,000236885 | 0,012203197 | Downregulated |
| AC141424 | -4,364560715 | 1,59167E-10 | 1,70823E-07 | Downregulated |
| AC242426 | -2,604579972 | 0,000170691 | 0,009725125 | Downregulated |
| AC244502 | -4,443091823 | 1,46504E-05 | 0,001588208 | Downregulated |
| AC247036 | -6,725883711 | 3,22776E-10 | 3,14922E-07 | Downregulated |
| ACY3 | -5,417530151 | 3,63203E-06 | 0,000590608 | Downregulated |
| ADAMTS19 | -6,830440194 | 4,30906E-07 | 0,000109243 | Downregulated |
| ADAMTS19-AS1 | -6,609215233 | 7,75669E-05 | 0,005461043 | Downregulated |
| ADGRG1 | -4,447223435 | 1,15112E-05 | 0,001347734 | Downregulated |
| AF064858 | -4,014567584 | 0,000269604 | 0,013375123 | Downregulated |
| AF233439 | -1,871961198 | 0,00104588 | 0,032101241 | Downregulated |
| AGGF1P3 | -6,029150395 | 0,000879716 | 0,028697276 | Downregulated |
| AGPAT5 | -1,254165583 | 0,000822573 | 0,02746243 | Downregulated |
| AKAP2 | -1,798674622 | 2,97426E-05 | 0,002705147 | Downregulated |
| AKAP6 | -3,141925974 | 2,93186E-06 | 0,000510255 | Downregulated |
| AKR1C3 | -3,098362222 | 0,000245186 | 0,01245148 | Downregulated |
| AL021331 | -4,685732849 | 0,001085133 | 0,033057745 | Downregulated |
| AL023693 | -5,001972711 | 0,000461105 | 0,018888421 | Downregulated |
| AL023973 | -2,78480811 | 0,000471944 | 0,019137492 | Downregulated |
| AL031118 | -2,426888909 | 1,18367E-05 | 0,001365967 | Downregulated |
| AL033529 | -4,037944738 | 0,001839889 | 0,046066437 | Downregulated |
| AL034397 | -3,827531085 | 3,2465E-06 | 0,000544415 | Downregulated |
| AL034550 | -1,69263017 | 0,000182867 | 0,01011642 | Downregulated |
| AL035696 | -7,666625221 | 6,01546E-07 | 0,000143467 | Downregulated |
| AL096865 | -3,394757717 | 0,001787952 | 0,045427295 | Downregulated |
| AL122008 | -3,755022669 | 0,000348711 | 0,015791085 | Downregulated |
| AL132657 | -1,621337641 | 0,002096255 | 0,049994914 | Downregulated |
| AL132822 | -4,522408775 | 1,94017E-05 | 0,00195823 | Downregulated |
| AL136087 | -5,361148773 | 0,000910024 | 0,029270777 | Downregulated |
| AL136313 | -2,808791745 | 0,000662808 | 0,024250492 | Downregulated |
| AL138889 | -3,453844235 | 7,12002E-07 | 0,000167331 | Downregulated |
| AL139351 | -7,042053824 | 5,12719E-08 | 1,88176E-05 | Downregulated |
| AL157385 | -2,91452519 | 0,001565712 | 0,041864967 | Downregulated |
| AL161645 | -3,195080411 | 0,000836555 | 0,027653564 | Downregulated |
| AL161665 | -1,881593264 | 0,000791893 | 0,027066443 | Downregulated |
| AL354712 | -5,85154797 | 9,1326E-06 | 0,001162125 | Downregulated |
| AL356275 | -3,761278131 | 0,000507683 | 0,020080929 | Downregulated |
| AL356750 | -1,8679204 | 0,000530435 | 0,020801986 | Downregulated |
| AL357143 | -4,800633501 | 0,000888127 | 0,028854716 | Downregulated |
| AL359643 | -1,238415864 | 4,01856E-06 | 0,000640522 | Downregulated |
| AL392086 | -5,374477756 | 1,24944E-06 | 0,000261223 | Downregulated |

| | | | | |
|---|---|---|---|---|
| AL445670 | -5,426209163 | 0,00013064 | 0,007966343 | Downregulated |
| AL513166 | -3,815621021 | 0,000284464 | 0,013905031 | Downregulated |
| AL590009 | -2,110466621 | 0,000316535 | 0,014892445 | Downregulated |
| AL590428 | -3,10206702 | 1,05921E-06 | 0,000225851 | Downregulated |
| AL603783 | -4,28900438 | 0,000238297 | 0,012236769 | Downregulated |
| ALOX12P2 | -2,688146084 | 0,001524626 | 0,041320176 | Downregulated |
| ANGPT1 | -5,044301308 | 5,51664E-05 | 0,004290323 | Downregulated |
| ANTXRL | -4,119314357 | 5,14634E-05 | 0,00409128 | Downregulated |
| AP000345 | -3,533315144 | 0,000161009 | 0,009425454 | Downregulated |
| AP000662 | -2,03331462 | 0,000323459 | 0,015155739 | Downregulated |
| AP000904 | -4,740334771 | 2,41016E-06 | 0,000437277 | Downregulated |
| AP000942 | -3,679080788 | 0,000600121 | 0,022763374 | Downregulated |
| AP001092 | -5,594373224 | 4,54419E-05 | 0,003732376 | Downregulated |
| AP001542 | -2,323368702 | 0,00132774 | 0,037629821 | Downregulated |
| AP002518 | -3,20067696 | 0,000992304 | 0,031018664 | Downregulated |
| AP005273 | -9,650982023 | 9,24597E-14 | 2,28994E-10 | Downregulated |
| AP006565 | -6,191688892 | 0,00158004 | 0,042045183 | Downregulated |
| APOBEC3D | -1,228457312 | 0,00128234 | 0,036896777 | Downregulated |
| APOBEC3H | -2,789974032 | 5,85202E-05 | 0,004518409 | Downregulated |
| ASB9 | -2,705738321 | 0,001709001 | 0,04418556 | Downregulated |
| ATP2B4 | -1,511143779 | 6,08935E-06 | 0,000875263 | Downregulated |
| B4GALT6 | -4,042442811 | 6,22834E-11 | 6,91496E-08 | Downregulated |
| BAALC-AS1 | -3,262515945 | 2,12329E-09 | 1,51919E-06 | Downregulated |
| BAHCC1 | -5,197761383 | 6,06088E-13 | 1,21964E-09 | Downregulated |
| BATF | -2,129788821 | 0,000982944 | 0,030755913 | Downregulated |
| BCL11A | -2,683659802 | 0,000564217 | 0,021729767 | Downregulated |
| BEND6 | -3,814659865 | 3,93287E-05 | 0,003338267 | Downregulated |
| BFSP1 | -3,21355506 | 0,000114708 | 0,007342441 | Downregulated |
| BIN2 | -1,789214073 | 0,000454832 | 0,018798733 | Downregulated |
| BIVM | -2,593161903 | 4,19822E-08 | 1,66877E-05 | Downregulated |
| BMPR1B | -4,011229112 | 0,001165657 | 0,034718453 | Downregulated |
| BNIP3P41 | -5,082547804 | 0,00019189 | 0,010489444 | Downregulated |
| BNIP3P9 | -4,139928389 | 0,000970162 | 0,03044473 | Downregulated |
| BRI3BP | -1,203536744 | 0,001210333 | 0,035571166 | Downregulated |
| BX276092 | -3,942674497 | 0,000696366 | 0,024939812 | Downregulated |
| BX321878 | -5,128828678 | 0,000819404 | 0,027442834 | Downregulated |
| BX571818 | -5,273368499 | 3,98503E-07 | 0,000103473 | Downregulated |
| C11orf74 | -2,224664385 | 0,001139078 | 0,034116175 | Downregulated |
| C17orf113 | -3,066038378 | 1,43579E-06 | 0,000283608 | Downregulated |
| C17orf51 | -2,592413917 | 3,82104E-05 | 0,003275601 | Downregulated |
| C1QTNF4 | -5,710461585 | 1,9226E-07 | 5,67827E-05 | Downregulated |
| C20orf197 | -4,65592145 | 2,82916E-08 | 1,18299E-05 | Downregulated |
| C3orf56 | -8,260219922 | 1,1069E-09 | 8,90972E-07 | Downregulated |
| CAPG | -2,224548501 | 8,66227E-05 | 0,005894084 | Downregulated |
| CAPN14 | -3,236179023 | 0,001119553 | 0,03371961 | Downregulated |
| CASP10 | -2,066821694 | 1,33105E-06 | 0,000271239 | Downregulated |

| | | | | |
|---|---|---|---|---|
| CBX2 | -1,993087344 | 0,000876567 | 0,028652627 | Downregulated |
| CCDC179 | -2,728468198 | 0,002014286 | 0,048616174 | Downregulated |
| CCDC180 | -2,723232869 | 0,001360569 | 0,038059295 | Downregulated |
| CCDC42 | -2,856621003 | 9,07884E-05 | 0,006089819 | Downregulated |
| CCL21 | -8,217804367 | 6,33455E-06 | 0,000897849 | Downregulated |
| CD163L1 | -2,595272555 | 0,001819436 | 0,045765925 | Downregulated |
| CD320 | -1,271034304 | 0,001843976 | 0,046066437 | Downregulated |
| CD3EAP | -1,333832752 | 0,000359702 | 0,016197664 | Downregulated |
| CD47 | -1,197061256 | 0,000656934 | 0,024090329 | Downregulated |
| CD5L | -5,913653359 | 2,49217E-06 | 0,000444712 | Downregulated |
| CD70 | -3,540781015 | 0,001064634 | 0,032583668 | Downregulated |
| CHML | -2,965147592 | 3,40003E-10 | 3,21973E-07 | Downregulated |
| CHRNA9 | -2,651036366 | 0,001771436 | 0,045122572 | Downregulated |
| CHST12 | -2,324919534 | 7,47105E-07 | 0,000173054 | Downregulated |
| CLEC11A | -3,928891897 | 2,41616E-07 | 6,70632E-05 | Downregulated |
| CLEC4C | -3,035950635 | 0,000574788 | 0,022084086 | Downregulated |
| CLEC4M | -8,817035683 | 6,32836E-05 | 0,004782966 | Downregulated |
| CLGN | -4,085714136 | 9,90059E-06 | 0,001216677 | Downregulated |
| CNR2 | -2,32344108 | 0,000642041 | 0,023802177 | Downregulated |
| COMMD7 | -1,318577998 | 2,63851E-05 | 0,00248398 | Downregulated |
| CPT1A | -1,241238813 | 0,000283044 | 0,013905031 | Downregulated |
| CREB3L4 | -1,006228043 | 0,000823097 | 0,02746243 | Downregulated |
| CRHBP | -2,863483192 | 0,000749653 | 0,026178506 | Downregulated |
| CTHRC1 | -4,336955731 | 1,54064E-08 | 7,08628E-06 | Downregulated |
| CTNNBIP1 | -1,235260935 | 0,001855539 | 0,046066437 | Downregulated |
| CTSG | -3,977533356 | 1,11764E-05 | 0,001331847 | Downregulated |
| CTSW | -3,693657622 | 2,29834E-06 | 0,000422854 | Downregulated |
| CUX2 | -4,367336992 | 0,000269109 | 0,013375123 | Downregulated |
| CXorf49 | -3,726410116 | 0,000946661 | 0,02997015 | Downregulated |
| CXorf49B | -3,678932555 | 0,00110921 | 0,033502087 | Downregulated |
| CXorf58 | -3,851308631 | 2,67768E-05 | 0,002484412 | Downregulated |
| CYP4F2 | -9,201051513 | 4,9075E-15 | 1,75563E-11 | Downregulated |
| CYTL1 | -3,335258701 | 0,002021779 | 0,048723977 | Downregulated |
| DANCR | -2,267630447 | 9,57139E-06 | 0,001194458 | Downregulated |
| DCPS | -1,009606777 | 0,000704949 | 0,025107582 | Downregulated |
| DENND1B | -1,420664098 | 0,000117401 | 0,007426247 | Downregulated |
| DFFB | -1,794536107 | 0,00036386 | 0,01629375 | Downregulated |
| DGUOK-AS1 | -1,279589443 | 0,001432058 | 0,039307724 | Downregulated |
| DHCR7 | -1,890151467 | 0,001428317 | 0,039238514 | Downregulated |
| DHRS4L2 | -1,094105059 | 0,000766432 | 0,02648306 | Downregulated |
| DNAH14 | -3,28154861 | 1,62897E-05 | 0,001725259 | Downregulated |
| DNAJC3-DT | -1,057307221 | 0,001656734 | 0,043226784 | Downregulated |
| DNAJC6 | -2,722216132 | 0,001054013 | 0,032320059 | Downregulated |
| DOCK7 | -1,245899659 | 0,001208724 | 0,035571166 | Downregulated |
| DPEP1 | -3,632029532 | 0,001034021 | 0,031847177 | Downregulated |
| DUSP27 | -7,260246091 | 4,92598E-07 | 0,000122001 | Downregulated |

| | | | | |
|---|---|---|---|---|
| EBNA1BP2 | -1,091485589 | 0,000825517 | 0,027473377 | Downregulated |
| EFHC2 | -2,947325575 | 0,001995227 | 0,048415124 | Downregulated |
| ELMO1 | -1,96278941 | 6,46506E-06 | 0,000905024 | Downregulated |
| ELMO1-AS1 | -2,076721518 | 0,000361795 | 0,016246481 | Downregulated |
| ELOVL6 | -2,398772365 | 0,000634496 | 0,023589902 | Downregulated |
| ENAM | -4,975929349 | 0,000198805 | 0,01076156 | Downregulated |
| EPDR1 | -1,117705323 | 0,00126834 | 0,036690036 | Downregulated |
| ERG | -3,122178519 | 0,001935598 | 0,047547101 | Downregulated |
| EVPL | -5,107836447 | 0,000129554 | 0,007945215 | Downregulated |
| FAAH | -2,43596298 | 6,43227E-05 | 0,004816271 | Downregulated |
| FADS1 | -1,792150912 | 0,00161697 | 0,042499258 | Downregulated |
| FAIM | -1,531774708 | 0,000451125 | 0,018777401 | Downregulated |
| FAM120C | -1,091695543 | 7,01047E-05 | 0,005118282 | Downregulated |
| FAM151A | -3,676750568 | 0,000867075 | 0,028486968 | Downregulated |
| FAM180B | -3,169187622 | 0,000377974 | 0,016670733 | Downregulated |
| FAM30A | -3,526860356 | 0,001906353 | 0,047054934 | Downregulated |
| FAR2 | -1,888743791 | 8,20731E-06 | 0,001052791 | Downregulated |
| FDCSP | -4,688822177 | 0,000820805 | 0,027442834 | Downregulated |
| FH | -1,487870199 | 0,000794978 | 0,02711431 | Downregulated |
| FHIT | -3,070576916 | 0,000177906 | 0,009944528 | Downregulated |
| FKBP4P1 | -5,589354093 | 7,31964E-06 | 0,000986295 | Downregulated |
| FLI1 | -1,376660231 | 0,000264931 | 0,013245312 | Downregulated |
| FMO5 | -2,417687539 | 5,89594E-05 | 0,004535811 | Downregulated |
| FNDC1 | -3,534322539 | 0,000658211 | 0,024109682 | Downregulated |
| FZD6 | -3,055083028 | 5,76164E-08 | 2,06119E-05 | Downregulated |
| GABRR3 | -5,152074107 | 0,001967426 | 0,047988811 | Downregulated |
| GALNT6 | -2,041559455 | 7,12882E-05 | 0,00518119 | Downregulated |
| GASK1A | -2,950804799 | 0,000231462 | 0,011981296 | Downregulated |
| GCSAML | -5,397378163 | 0,000222959 | 0,011634685 | Downregulated |
| GFI1B | -5,175215797 | 2,53927E-05 | 0,002411703 | Downregulated |
| GGH | -2,602526256 | 0,000666461 | 0,024335278 | Downregulated |
| GNB4 | -1,654490606 | 1,22105E-05 | 0,001395615 | Downregulated |
| GNG2 | -2,051481624 | 1,71874E-05 | 0,001790881 | Downregulated |
| GNPTAB | -1,279615678 | 0,000169343 | 0,009689896 | Downregulated |
| GOLGA6L7 | -4,738215474 | 1,10548E-05 | 0,001329354 | Downregulated |
| GOLGA8N | -3,879016036 | 6,25043E-05 | 0,004757566 | Downregulated |
| GOLGA8O | -3,899127242 | 0,000116848 | 0,007405794 | Downregulated |
| GOLGA8Q | -3,770924933 | 0,000142734 | 0,008573887 | Downregulated |
| GOLGA8R | -3,41485916 | 0,000671839 | 0,024495734 | Downregulated |
| GPA33 | -3,96975678 | 1,31519E-06 | 0,000269715 | Downregulated |
| GPR150 | -2,408005817 | 0,000396041 | 0,017058291 | Downregulated |
| GPSM2 | -1,612669792 | 0,000295304 | 0,014254737 | Downregulated |
| GPX7 | -1,296778314 | 0,000582004 | 0,022308092 | Downregulated |
| GREM1 | -4,838411269 | 1,10652E-05 | 0,001329354 | Downregulated |
| GRIK5 | -3,362365571 | 0,001474376 | 0,040263333 | Downregulated |
| GRIN2B | -3,842772903 | 0,001285047 | 0,036898106 | Downregulated |

| | | | | |
|---|---|---|---|---|
| GSEC | -1,180636865 | 0,001671416 | 0,043479495 | Downregulated |
| GTF3A | -1,327350854 | 0,000973318 | 0,030484352 | Downregulated |
| GTSF1 | -6,332830146 | 1,67062E-08 | 7,57591E-06 | Downregulated |
| GUCY1A1 | -2,656601245 | 0,000952812 | 0,030076167 | Downregulated |
| GYPE | -3,683725614 | 1,08985E-05 | 0,001324145 | Downregulated |
| HACD1 | -3,808447699 | 4,4822E-08 | 1,74322E-05 | Downregulated |
| HCG11 | -2,420129146 | 0,001312902 | 0,037383438 | Downregulated |
| HELB | -1,295712104 | 0,000587927 | 0,022508314 | Downregulated |
| HEMGN | -5,057631037 | 4,42158E-11 | 5,08434E-08 | Downregulated |
| HHIP | -2,74539014 | 0,000240139 | 0,01226962 | Downregulated |
| HHIP-AS1 | -3,497826932 | 4,83814E-05 | 0,003933678 | Downregulated |
| HMHB1 | -3,352837324 | 0,000873878 | 0,028593762 | Downregulated |
| HOMER2P1 | -4,87430678 | 4,0867E-05 | 0,003435497 | Downregulated |
| HORMAD2 | -5,673318822 | 3,93994E-05 | 0,003338267 | Downregulated |
| HOXA9 | -4,841585528 | 0,000599683 | 0,022763374 | Downregulated |
| HOXB2 | -2,458882407 | 0,001727961 | 0,044366157 | Downregulated |
| HPDL | -2,717335711 | 9,22388E-05 | 0,006174249 | Downregulated |
| HPGDS | -5,546130579 | 6,28241E-06 | 0,000895021 | Downregulated |
| HSH2D | -3,243407398 | 0,001243445 | 0,036165491 | Downregulated |
| HSPD1P11 | -2,802096385 | 0,001374746 | 0,038289537 | Downregulated |
| HSPD1P12 | -3,976766745 | 7,86168E-07 | 0,000179519 | Downregulated |
| HYLS1 | -1,05446296 | 0,000642423 | 0,023802177 | Downregulated |
| IFITM5 | -2,920436597 | 0,001210858 | 0,035571166 | Downregulated |
| IFRD2 | -1,074275623 | 0,0014802 | 0,040361491 | Downregulated |
| IGHV1OR15-6 | -6,363581183 | 8,83336E-05 | 0,005962423 | Downregulated |
| IGHV3-19 | -5,693035585 | 8,00717E-07 | 0,000180284 | Downregulated |
| IGHV3-42 | -5,788436871 | 7,26019E-05 | 0,005210074 | Downregulated |
| IGHV3-79 | -3,956762881 | 0,000363245 | 0,016288855 | Downregulated |
| IGHV4-80 | -5,607588427 | 4,75259E-06 | 0,000735669 | Downregulated |
| IGHVII-43-1 | -6,132970999 | 7,20764E-05 | 0,005203235 | Downregulated |
| IGHVII-60-1 | -5,057669367 | 3,14164E-05 | 0,002837429 | Downregulated |
| IGHVII-78-1 | -4,377438118 | 0,000703734 | 0,025092048 | Downregulated |
| IGHVIII-11-1 | -4,441444013 | 0,000484056 | 0,019457134 | Downregulated |
| IGHVIII-38-1 | -4,08740362 | 0,001552267 | 0,041579314 | Downregulated |
| IGHVIII-44 | -4,946170167 | 4,61476E-05 | 0,003769528 | Downregulated |
| IGLL1 | -11,34708151 | 4,06856E-15 | 1,63744E-11 | Downregulated |
| IL17RE | -3,232138708 | 0,000287819 | 0,014019528 | Downregulated |
| IL36A | -2,508088101 | 0,000306703 | 0,014586281 | Downregulated |
| IMPA2 | -2,061760531 | 0,000116726 | 0,007405794 | Downregulated |
| INPP4B | -2,513576951 | 0,000191258 | 0,010489444 | Downregulated |
| IQCH-AS1 | -1,191511834 | 0,000793265 | 0,027084581 | Downregulated |
| IQGAP2 | -2,410453004 | 0,000185798 | 0,010243378 | Downregulated |
| IRGM | -2,205672866 | 0,000973059 | 0,030484352 | Downregulated |
| ITPRIPL1 | -1,58624615 | 0,001425951 | 0,039233172 | Downregulated |
| KCNK16 | -7,472827972 | 1,3017E-06 | 0,00026866 | Downregulated |
| KCNK17 | -7,875343005 | 1,79234E-08 | 7,90519E-06 | Downregulated |

| | | | | |
|---|---|---|---|---|
| KCTD19 | -2,718009354 | 0,000625164 | 0,023348352 | Downregulated |
| KLHL33 | -3,859863477 | 0,000464345 | 0,018924703 | Downregulated |
| KLRF2 | -5,834438181 | 5,14176E-05 | 0,00409128 | Downregulated |
| LAT2 | -2,733736452 | 0,001553929 | 0,041589227 | Downregulated |
| LDHAL6EP | -3,185971315 | 3,28018E-05 | 0,002925541 | Downregulated |
| LINC00051 | -4,137964157 | 8,85468E-05 | 0,005964312 | Downregulated |
| LINC002481 | -3,33330953 | 6,19307E-06 | 0,000886214 | Downregulated |
| LINC00426 | -1,78378844 | 0,000105859 | 0,006927496 | Downregulated |
| LINC00528 | -1,77666981 | 0,001122805 | 0,033754395 | Downregulated |
| LINC00539 | -2,39368378 | 4,96536E-06 | 0,000760379 | Downregulated |
| LINC00664 | -2,801246516 | 0,000104966 | 0,006903253 | Downregulated |
| LINC00692 | -4,710140497 | 0,001423975 | 0,039233172 | Downregulated |
| LINC00707 | -6,233865806 | 4,76566E-08 | 1,80518E-05 | Downregulated |
| LINC00887 | -4,213811501 | 0,000820089 | 0,027442834 | Downregulated |
| LINC00891 | -2,398625348 | 0,000783113 | 0,02690915 | Downregulated |
| LINC00987 | -2,589162064 | 0,000710047 | 0,025233304 | Downregulated |
| LINC01016 | -4,647049523 | 0,001542812 | 0,041556009 | Downregulated |
| LINC01134 | -2,247570698 | 0,000151733 | 0,008963958 | Downregulated |
| LINC01182 | -4,279776601 | 0,000149115 | 0,008862924 | Downregulated |
| LINC01215 | -1,565678152 | 5,12142E-05 | 0,00409128 | Downregulated |
| LINC01268 | -3,599420027 | 0,001203028 | 0,03553569 | Downregulated |
| LINC01342 | -6,238500355 | 1,29992E-05 | 0,001461 | Downregulated |
| LINC01378 | -6,030376408 | 0,000165641 | 0,009619318 | Downregulated |
| LINC01579 | -3,830798265 | 0,00052675 | 0,020682634 | Downregulated |
| LINC01597 | -3,199843727 | 0,001593428 | 0,042140991 | Downregulated |
| LINC01793 | -4,313708972 | 0,000420322 | 0,017844164 | Downregulated |
| LINC01825 | -3,076228843 | 0,000116757 | 0,007405794 | Downregulated |
| LINC01833 | -6,542233753 | 0,000145211 | 0,008706432 | Downregulated |
| LINC01835 | -10,59447718 | 1,24107E-05 | 0,001407001 | Downregulated |
| LINC01934 | -3,688908171 | 8,63754E-10 | 7,3185E-07 | Downregulated |
| LINC01939 | -5,751662881 | 0,000451666 | 0,018777401 | Downregulated |
| LINC01958 | -5,413008485 | 0,001766809 | 0,045090672 | Downregulated |
| LINC01971 | -5,440447957 | 5,55411E-07 | 0,000134455 | Downregulated |
| LINC02014 | -2,77128186 | 0,000129878 | 0,007949949 | Downregulated |
| LINC02036 | -7,160689049 | 1,48213E-06 | 0,000289849 | Downregulated |
| LINC02059 | -8,359836633 | 1,02678E-07 | 3,27318E-05 | Downregulated |
| LINC02203 | -7,027650194 | 7,87559E-06 | 0,0010266 | Downregulated |
| LINC02285 | -3,599373322 | 0,000130322 | 0,007962023 | Downregulated |
| LINC02405 | -6,862340982 | 0,00056418 | 0,021729767 | Downregulated |
| LINC02435 | -5,223545163 | 9,7997E-05 | 0,006519029 | Downregulated |
| LINC02470 | -5,500508359 | 0,000113526 | 0,007295802 | Downregulated |
| LOXL1-AS1 | -2,509225344 | 7,91594E-05 | 0,005528623 | Downregulated |
| LPAR6 | -2,263559901 | 3,53161E-05 | 0,003089871 | Downregulated |
| LRRC38 | -9,539311339 | 6,93235E-15 | 2,23201E-11 | Downregulated |
| LRRCC1 | -1,622359617 | 0,001846642 | 0,046066437 | Downregulated |
| LSM3P4 | -3,89152608 | 7,78526E-05 | 0,005461043 | Downregulated |

| | | | | |
|---|---|---|---|---|
| LTB | -2,112348035 | 0,001426905 | 0,039233172 | Downregulated |
| LYL1 | -2,319755794 | 0,000772721 | 0,026665914 | Downregulated |
| MALRD1 | -5,462926981 | 0,000383655 | 0,016806191 | Downregulated |
| MAP10 | -1,241215396 | 7,35195E-06 | 0,000986295 | Downregulated |
| MAPRE1P3 | -20,75471061 | 1,62444E-17 | 1,30755E-13 | Downregulated |
| MBNL1-AS1 | -3,74548043 | 3,05687E-06 | 0,000520772 | Downregulated |
| MBNL3 | -1,329649494 | 0,000461331 | 0,018888421 | Downregulated |
| MBOAT2 | -2,268721333 | 0,001605219 | 0,042293965 | Downregulated |
| MDFI | -5,246247143 | 1,70556E-08 | 7,62691E-06 | Downregulated |
| MDFIC | -2,789805631 | 6,42181E-05 | 0,004816271 | Downregulated |
| ME3 | -1,437712489 | 0,000697926 | 0,024967903 | Downregulated |
| MED12L | -2,401441566 | 0,001330194 | 0,037634662 | Downregulated |
| MFHAS1 | -2,047519024 | 0,001235688 | 0,036011321 | Downregulated |
| MIR1273H | -2,288295627 | 0,001587976 | 0,042138717 | Downregulated |
| MIR3186 | -5,591924552 | 2,48014E-07 | 6,82505E-05 | Downregulated |
| MIR4740 | -4,319317039 | 0,000942332 | 0,029936712 | Downregulated |
| MLC1 | -5,244995023 | 5,67821E-06 | 0,000825879 | Downregulated |
| MRM1 | -1,067010028 | 0,000303323 | 0,014468768 | Downregulated |
| MS4A3 | -6,947969922 | 0,000366128 | 0,016349823 | Downregulated |
| MTCO1P46 | -5,547441102 | 0,001954168 | 0,047918621 | Downregulated |
| MTND4P23 | -2,083600633 | 0,000447986 | 0,018697901 | Downregulated |
| MTND5P25 | -2,605972506 | 0,000264641 | 0,013245312 | Downregulated |
| MYCN | -4,947164527 | 0,000428507 | 0,018128734 | Downregulated |
| MYO18B | -6,184014222 | 0,000620161 | 0,023217803 | Downregulated |
| MYOT | -1,954226633 | 0,001956899 | 0,047918621 | Downregulated |
| MYOZ3 | -2,93904126 | 0,001592186 | 0,042140991 | Downregulated |
| MYT1L | -10,48695429 | 3,50867E-11 | 4,51874E-08 | Downregulated |
| NDST3 | -8,836766087 | 9,92717E-15 | 2,90568E-11 | Downregulated |
| NFE2 | -4,009809901 | 0,000124288 | 0,007710386 | Downregulated |
| NMRK2 | -3,885545203 | 0,001614258 | 0,042462642 | Downregulated |
| NRROS | -1,617550978 | 0,000599126 | 0,022763374 | Downregulated |
| NRXN2 | -3,972003655 | 1,37833E-05 | 0,00153028 | Downregulated |
| NSG1 | -3,048418125 | 0,000802522 | 0,027270526 | Downregulated |
| OPN3 | -1,92011808 | 7,05688E-06 | 0,000966853 | Downregulated |
| OR3A1 | -6,21597916 | 0,00093323 | 0,029779197 | Downregulated |
| OR3A3 | -5,565674195 | 0,000871079 | 0,028531176 | Downregulated |
| OR7E14P | -3,187158143 | 0,000235034 | 0,012127225 | Downregulated |
| ORAI1 | -2,352030157 | 0,000149789 | 0,008881716 | Downregulated |
| OSGEPL1-AS1 | -1,152007825 | 0,00122606 | 0,035877535 | Downregulated |
| OSTCP6 | -5,286456089 | 0,000884488 | 0,028796499 | Downregulated |
| PAG1 | -2,27851228 | 0,000547936 | 0,021332388 | Downregulated |
| PAN3-AS1 | -1,624211396 | 5,98283E-05 | 0,004586412 | Downregulated |
| PCDH10 | -8,667345143 | 2,78387E-12 | 4,26821E-09 | Downregulated |
| PCDH11X | -5,677339459 | 0,00134838 | 0,037816885 | Downregulated |
| PCDH15 | -6,193920938 | 1,70027E-05 | 0,001777388 | Downregulated |
| PCYT1B | -3,112480104 | 5,40641E-05 | 0,004245613 | Downregulated |

| | | | | |
|---|---|---|---|---|
| PDCD6IPP2 | -2,169780716 | 0,001727522 | 0,044366157 | Downregulated |
| PDK2 | -1,633066864 | 0,000727507 | 0,025627505 | Downregulated |
| PGBD1 | -1,644005366 | 1,64006E-07 | 5,02906E-05 | Downregulated |
| PGBD4P1 | -7,293411274 | 1,07303E-05 | 0,001308643 | Downregulated |
| PGLYRP2 | -2,17487672 | 0,000538334 | 0,020983947 | Downregulated |
| PIEZO1 | -1,308058391 | 0,001131847 | 0,033994476 | Downregulated |
| PIGW | -1,254024425 | 0,000494073 | 0,019738004 | Downregulated |
| PLCB1-IT1 | -2,926166125 | 0,001001583 | 0,031157773 | Downregulated |
| PLD4 | -3,698314762 | 0,000900438 | 0,029137074 | Downregulated |
| PLEKHG4B | -4,500571958 | 0,000114676 | 0,007342441 | Downregulated |
| PM20D2 | -1,417130604 | 0,000781935 | 0,026897382 | Downregulated |
| POGLUT2 | -2,078032022 | 0,001716904 | 0,044258744 | Downregulated |
| PRKAR1B | -1,917424894 | 3,15048E-06 | 0,000531078 | Downregulated |
| PRKRA-AS1 | -1,038632379 | 0,000808896 | 0,027328477 | Downregulated |
| PRR5L | -2,686595124 | 0,000748441 | 0,026166396 | Downregulated |
| PRSS57 | -11,4185472 | 1,69471E-05 | 0,001777353 | Downregulated |
| PRTFDC1 | -3,126358965 | 8,36496E-06 | 0,001068757 | Downregulated |
| R3HDM2P1 | -7,908439694 | 4,15277E-06 | 0,000654312 | Downregulated |
| RAB37 | -1,477563639 | 3,65926E-06 | 0,000591184 | Downregulated |
| RAP1GAP2 | -2,801532002 | 0,000785481 | 0,026933052 | Downregulated |
| RARRES2P2 | -2,861115991 | 0,001088225 | 0,033116796 | Downregulated |
| RARRES2P4 | -3,160847084 | 0,001671821 | 0,043479495 | Downregulated |
| RETN | -3,986267611 | 5,45671E-05 | 0,00425825 | Downregulated |
| RF01293 | -1,95709998 | 0,000446628 | 0,018697901 | Downregulated |
| RFESD | -2,015192645 | 0,000678958 | 0,024562277 | Downregulated |
| RFLNB | -1,93419659 | 1,91016E-05 | 0,001934009 | Downregulated |
| RGS9 | -3,070825696 | 0,001988978 | 0,048367927 | Downregulated |
| RHOBTB3 | -2,614515677 | 0,000943832 | 0,029936712 | Downregulated |
| RIMS3 | -2,373653033 | 0,001097986 | 0,033225415 | Downregulated |
| RN7SL268P | -1,745597908 | 0,000931297 | 0,029747 | Downregulated |
| RN7SL617P | -4,784458932 | 0,000240461 | 0,01226962 | Downregulated |
| RN7SL684P | -3,712822699 | 4,62454E-05 | 0,003769528 | Downregulated |
| RN7SL7P | -4,704714043 | 0,001114072 | 0,033585919 | Downregulated |
| RNA5SP94 | -4,978321557 | 0,001910883 | 0,047109276 | Downregulated |
| RNASEH1-AS1 | -1,000398571 | 5,31634E-05 | 0,004195351 | Downregulated |
| RNF125 | -1,585072825 | 8,45155E-05 | 0,005789669 | Downregulated |
| RNU6-1004P | -2,724673534 | 0,000123396 | 0,007684671 | Downregulated |
| RNU6-101P | -1,70410282 | 0,001996934 | 0,048415124 | Downregulated |
| RNU6-236P | -4,194509725 | 0,00082925 | 0,027526444 | Downregulated |
| RNU6-345P | -4,018010889 | 0,001686667 | 0,043689153 | Downregulated |
| RNU6-915P | -2,684698891 | 0,000239863 | 0,01226962 | Downregulated |
| RNU6-925P | -2,381593242 | 1,79576E-07 | 5,40355E-05 | Downregulated |
| RNU6ATAC36P | -4,960716009 | 0,000746339 | 0,026151141 | Downregulated |
| RPARP-AS1 | -1,582980181 | 1,22907E-08 | 6,11966E-06 | Downregulated |
| RPL31P33 | -4,378375843 | 0,000458218 | 0,018862582 | Downregulated |
| RPS10P7 | -1,321576607 | 0,000256545 | 0,012926416 | Downregulated |

| | | | | |
|---|---|---|---|---|
| RPS2P52 | -21,52878752 | 4,19607E-09 | 2,4857E-06 | Downregulated |
| RPS6P2 | -4,976708768 | 4,36731E-05 | 0,003614764 | Downregulated |
| RPUSD3 | -1,092291935 | 0,000216876 | 0,011428427 | Downregulated |
| RRM2P3 | -3,648983063 | 0,000457403 | 0,0188566 | Downregulated |
| RTN4R | -2,463204677 | 1,49446E-05 | 0,001609267 | Downregulated |
| RUNX1 | -1,616773108 | 0,000394677 | 0,017058291 | Downregulated |
| RUNX2 | -2,901961191 | 0,000454565 | 0,018798733 | Downregulated |
| S100Z | -5,36670496 | 1,26409E-08 | 6,16664E-06 | Downregulated |
| S1PR4 | -1,462588032 | 0,001726911 | 0,044366157 | Downregulated |
| SACS | -1,128156611 | 0,001600809 | 0,042246915 | Downregulated |
| SCGB3A1 | -5,935883601 | 1,03743E-06 | 0,000222681 | Downregulated |
| SCMH1 | -1,074166708 | 0,000942489 | 0,029936712 | Downregulated |
| SCN1A-AS1 | -4,60214694 | 1,7E-07 | 5,16368E-05 | Downregulated |
| SCN2A | -7,363485843 | 4,10334E-10 | 3,66987E-07 | Downregulated |
| SCN9A | -4,022927947 | 9,71962E-08 | 3,12943E-05 | Downregulated |
| SCUBE1 | -3,685829326 | 0,001354488 | 0,037955147 | Downregulated |
| SELL | -3,540540527 | 1,84767E-06 | 0,000349937 | Downregulated |
| SEPHS1P2 | -6,205035475 | 4,12041E-05 | 0,003454814 | Downregulated |
| SFRP4 | -5,177824155 | 2,83346E-06 | 0,000501257 | Downregulated |
| SFXN4 | -1,428828651 | 9,78689E-05 | 0,006519029 | Downregulated |
| SHD | -4,371978108 | 4,16604E-06 | 0,000654312 | Downregulated |
| SHISA3 | -3,318385001 | 0,000548782 | 0,021339526 | Downregulated |
| SLA2 | -2,314537636 | 0,000153558 | 0,009055127 | Downregulated |
| SLC15A4 | -1,19066705 | 0,000329107 | 0,015256589 | Downregulated |
| SLC18A2 | -4,935529858 | 7,73751E-06 | 0,001020416 | Downregulated |
| SLC22A16 | -5,475810748 | 3,0328E-07 | 8,13727E-05 | Downregulated |
| SLC22A20P | -3,972420226 | 1,43191E-05 | 0,00156814 | Downregulated |
| SLC22A3 | -3,622398461 | 0,001269457 | 0,036690036 | Downregulated |
| SLC27A5 | -1,239875399 | 0,001709948 | 0,04418556 | Downregulated |
| SLC35F3 | -3,473339188 | 0,000200626 | 0,010801946 | Downregulated |
| SLC38A5 | -2,644912688 | 2,91893E-06 | 0,000510255 | Downregulated |
| SLC40A1 | -2,763398518 | 3,64697E-05 | 0,003173555 | Downregulated |
| SLC44A5 | -4,521688868 | 0,000164248 | 0,009580262 | Downregulated |
| SLC9B2 | -1,209307653 | 0,000646805 | 0,023827449 | Downregulated |
| SLFN13 | -2,234975672 | 0,001338155 | 0,037690993 | Downregulated |
| SMAD6 | -2,901089758 | 0,00016909 | 0,009689896 | Downregulated |
| SMIM24 | -3,528972431 | 2,22298E-08 | 9,54308E-06 | Downregulated |
| SMIM3 | -1,932007084 | 0,001321434 | 0,037584983 | Downregulated |
| SMIM33 | -2,201342435 | 0,000201951 | 0,010855132 | Downregulated |
| SMYD2 | -1,03997434 | 0,000962118 | 0,030221758 | Downregulated |
| SNHG20 | -1,207264253 | 1,84167E-05 | 0,001882419 | Downregulated |
| SNHG4 | -1,681060112 | 0,000128314 | 0,007884236 | Downregulated |
| SNX16 | -1,545260535 | 5,66315E-05 | 0,004387187 | Downregulated |
| SOAT2 | -4,375296116 | 6,5326E-06 | 0,000906595 | Downregulated |
| SORL1 | -2,145155358 | 0,000200211 | 0,010797647 | Downregulated |
| SPATA13-AS1 | -1,570657701 | 0,001721685 | 0,044323653 | Downregulated |

| | | | | |
|---|---|---|---|---|
| SPATA16 | -5,346559884 | 0,001987018 | 0,048356771 | Downregulated |
| SPDYE4 | -5,982264034 | 1,69745E-06 | 0,000325314 | Downregulated |
| SPINK2 | -7,285669604 | 3,24769E-12 | 4,753E-09 | Downregulated |
| SPNS3 | -2,744685752 | 0,001535127 | 0,041436946 | Downregulated |
| SPSB4 | -3,844830927 | 2,41747E-06 | 0,000437277 | Downregulated |
| SRGAP3 | -2,020740451 | 0,000461695 | 0,018888421 | Downregulated |
| SRY | -9,043156725 | 0,000341723 | 0,015627788 | Downregulated |
| STAB2 | -4,767177273 | 0,000799111 | 0,02722643 | Downregulated |
| STARD4-AS1 | -1,366831734 | 0,001215877 | 0,035629783 | Downregulated |
| STN1 | -1,398473699 | 7,9607E-08 | 2,64238E-05 | Downregulated |
| STRA6LP | -3,19417335 | 0,000815635 | 0,027442834 | Downregulated |
| SV2C | -4,93371452 | 0,000298782 | 0,014379474 | Downregulated |
| SVIP | -1,90115863 | 3,71773E-05 | 0,003226405 | Downregulated |
| SYNJ2 | -1,743586693 | 3,89931E-05 | 0,003321325 | Downregulated |
| SYT16 | -4,72681861 | 4,12226E-06 | 0,000653815 | Downregulated |
| TAF4B | -1,750761454 | 0,000100125 | 0,00664688 | Downregulated |
| TBC1D4 | -2,210418383 | 0,000643697 | 0,023821973 | Downregulated |
| TBX1 | -4,30885465 | 1,11425E-05 | 0,001331847 | Downregulated |
| TBXT | -7,6132901 | 0,000310525 | 0,014681325 | Downregulated |
| TEN1-CDK3 | -2,394900898 | 0,000401659 | 0,017242935 | Downregulated |
| TESMIN | -1,842374963 | 0,000135681 | 0,008226979 | Downregulated |
| TESPA1 | -1,699026718 | 2,03606E-06 | 0,00037893 | Downregulated |
| TEX30 | -1,94318938 | 0,000604831 | 0,022854082 | Downregulated |
| TIAF1 | -1,393026915 | 0,001844279 | 0,046066437 | Downregulated |
| TIGD2 | -1,166614339 | 0,001848387 | 0,046066437 | Downregulated |
| TLX2 | -6,591052425 | 8,64999E-07 | 0,000193405 | Downregulated |
| TM7SF3 | -1,405093619 | 2,67628E-05 | 0,002484412 | Downregulated |
| TMC3-AS1 | -4,016840705 | 1,58945E-06 | 0,00030644 | Downregulated |
| TMED8 | -1,157921524 | 0,000128273 | 0,007884236 | Downregulated |
| TMEM254-AS1 | -1,616703674 | 0,001384931 | 0,038496645 | Downregulated |
| TMEM26 | -3,529322656 | 0,000175445 | 0,009841093 | Downregulated |
| TMEM72 | -3,291180555 | 7,26566E-05 | 0,005210074 | Downregulated |
| TMIGD2 | -2,538717793 | 0,00029115 | 0,014117699 | Downregulated |
| TMOD2 | -1,356641645 | 0,000348359 | 0,015791085 | Downregulated |
| TNFSF4 | -3,443240077 | 7,06392E-05 | 0,005145638 | Downregulated |
| TRAF3IP2 | -1,816756776 | 3,75372E-05 | 0,003248888 | Downregulated |
| TRAJ51 | -5,330817646 | 0,000494924 | 0,019738004 | Downregulated |
| TRAJ55 | -4,668913268 | 0,001092547 | 0,033123107 | Downregulated |
| TRAJ56 | -6,167394983 | 6,09788E-05 | 0,0046635 | Downregulated |
| TRAJ57 | -5,454518726 | 0,00033675 | 0,015467777 | Downregulated |
| TRAJ59 | -5,577743785 | 0,00125647 | 0,036478422 | Downregulated |
| TRAJ61 | -6,58788532 | 0,000127174 | 0,007874257 | Downregulated |
| TRAPPC6A | -1,317569728 | 0,00075726 | 0,026273172 | Downregulated |
| TRAV22 | -4,798662202 | 8,44134E-08 | 2,77333E-05 | Downregulated |
| TRAV30 | -5,897924576 | 9,00064E-07 | 0,000199858 | Downregulated |
| TRAV31 | -6,40175017 | 5,3963E-05 | 0,004245613 | Downregulated |

| | | | | |
|---|---|---|---|---|
| TRAV38-2DV8 | -3,790533245 | 0,000721303 | 0,025520654 | Downregulated |
| TRAV41 | -3,94279988 | 0,000171448 | 0,009725125 | Downregulated |
| TRBJ1-1 | -6,185503183 | 4,32303E-05 | 0,003603031 | Downregulated |
| TRBV4-2 | -3,195607456 | 0,000346535 | 0,015781326 | Downregulated |
| TRDC | -5,785016376 | 9,9898E-06 | 0,001222972 | Downregulated |
| TRDJ1 | -6,141758777 | 6,56215E-05 | 0,004890774 | Downregulated |
| TRDJ2 | -4,01408393 | 0,001921774 | 0,047341501 | Downregulated |
| TRDJ3 | -6,460640002 | 1,41203E-06 | 0,000281501 | Downregulated |
| TRDV3 | -4,68121887 | 2,94664E-05 | 0,002687619 | Downregulated |
| TREML2 | -2,178335738 | 0,001570679 | 0,04188448 | Downregulated |
| TRGV9 | -5,785622907 | 6,51138E-06 | 0,000906595 | Downregulated |
| TSPAN2 | -2,12416964 | 0,000993661 | 0,031021603 | Downregulated |
| TTLL2 | -6,169597589 | 2,32245E-07 | 6,50226E-05 | Downregulated |
| TXNRD3 | -2,006573896 | 0,001229087 | 0,035877535 | Downregulated |
| U91328 | -1,394229615 | 0,000375658 | 0,016654642 | Downregulated |
| UCA1 | -2,906209281 | 8,77257E-05 | 0,005946323 | Downregulated |
| UGT3A2 | -10,56852774 | 8,61323E-20 | 1,3866E-15 | Downregulated |
| UMODL1 | -4,62606423 | 1,40748E-08 | 6,65896E-06 | Downregulated |
| UMODL1-AS1 | -5,124716982 | 8,93382E-13 | 1,69201E-09 | Downregulated |
| UOX | -2,369682334 | 0,001417709 | 0,039125467 | Downregulated |
| UROC1 | -3,599717445 | 0,00046622 | 0,018977116 | Downregulated |
| USP20 | -2,056354545 | 0,000994325 | 0,031021603 | Downregulated |
| USP41 | -2,110444995 | 0,001781509 | 0,045322666 | Downregulated |
| VANGL1 | -1,663806232 | 0,00093087 | 0,029747 | Downregulated |
| VPREB1 | -8,063455527 | 3,84379E-12 | 5,27586E-09 | Downregulated |
| VPS9D1-AS1 | -2,245246281 | 0,00052016 | 0,020518382 | Downregulated |
| WDR12 | -1,214389275 | 0,00021111 | 0,011161078 | Downregulated |
| WDR64 | -3,544126356 | 0,000102003 | 0,006757621 | Downregulated |
| XG | -5,640356846 | 9,69081E-06 | 0,001204691 | Downregulated |
| XKR3 | -4,125190756 | 0,001907218 | 0,047054934 | Downregulated |
| XKR5 | -2,180921023 | 0,000237308 | 0,012205466 | Downregulated |
| YBX2 | -4,593785014 | 6,66918E-05 | 0,004959069 | Downregulated |
| Z97198 | -3,927749312 | 0,000784233 | 0,026918921 | Downregulated |
| Z97989 | -1,37864228 | 0,000264467 | 0,013245312 | Downregulated |
| ZBTB8A | -2,603288083 | 0,000105794 | 0,006927496 | Downregulated |
| ZNF114 | -3,960329013 | 2,68527E-05 | 0,002484412 | Downregulated |
| ZNF280B | -1,940648172 | 6,4249E-05 | 0,004816271 | Downregulated |
| ZNF385C | -4,146580346 | 2,47447E-08 | 1,0483E-05 | Downregulated |
| ZNF391 | -2,434170382 | 0,000409993 | 0,017530623 | Downregulated |
| ZNF485 | -1,113637182 | 0,000857703 | 0,028278717 | Downregulated |
| ZNF492 | -3,924498826 | 0,000331407 | 0,015308898 | Downregulated |
| ZNF555 | -2,275006576 | 0,000107836 | 0,007014157 | Downregulated |
| ZNF57 | -1,619015466 | 0,001018855 | 0,031512095 | Downregulated |
| ZNF595 | -3,06827557 | 2,31002E-05 | 0,002267552 | Downregulated |
| ZNF620 | -1,666243654 | 0,002012042 | 0,048616174 | Downregulated |
| ZNF660 | -1,661672256 | 0,000687174 | 0,024748247 | Downregulated |

| | | | | |
|---|---|---|---|---|
| ZNF710-AS1 | -1,20631051 | 2,19731E-05 | 0,002183546 | Downregulated |
| ZNF718 | -1,825671339 | 2,41693E-05 | 0,002350994 | Downregulated |
| ZNF74 | -1,455054237 | 0,000479743 | 0,019368214 | Downregulated |
| ZNF849P | -4,958901123 | 1,38612E-05 | 0,001533641 | Downregulated |
| ZNF861P | -5,196696831 | 0,001566832 | 0,041864967 | Downregulated |

**Supplementary Table 5:** Differentially expressed genes in *NOTCH1* mutated T-LBL versus *NOTCH1*-WT T-LBL (n=101). Genes with an FDR-adjusted p-value ≤ 0.05 and an absolute log2-transformed log fold change of at least 1 were considered significantly differentially expressed.

| Gene | log2FoldChange | P-value | FDR-adjusted p-value | Expression |
|---|---|---|---|---|
| AC004988 | 3,542231912 | 5,94847E-05 | 0,033956228 | Upregulated |
| AC025437 | 4,101489256 | 0,000142807 | 0,045956771 | Upregulated |
| AC087360 | 26,15243749 | 2,41295E-14 | 6,98862E-10 | Upregulated |
| AC112206 | 5,411250182 | 2,54132E-05 | 0,018904403 | Upregulated |
| AC118758 | 3,626935093 | 0,00015212 | 0,046815446 | Upregulated |
| ANKRD1 | 3,234114128 | 0,000141881 | 0,045956771 | Upregulated |
| BCHE | 5,162462722 | 1,77795E-06 | 0,00514949 | Upregulated |
| BNC1 | 3,692444841 | 5,9668E-05 | 0,033956228 | Upregulated |
| CCN5 | 3,712739236 | 0,000122285 | 0,044831983 | Upregulated |
| CHI3L1 | 3,321842685 | 2,291E-05 | 0,01843172 | Upregulated |
| CLIC5 | 3,639554747 | 6,23284E-05 | 0,033956228 | Upregulated |
| CMYA5 | 2,06994267 | 0,000173145 | 0,049943983 | Upregulated |
| CSDC2 | 3,666658101 | 9,30717E-06 | 0,01078254 | Upregulated |
| DPYS | 4,687835458 | 7,28647E-08 | 0,00035173 | Upregulated |
| EEF1A1P12 | 1,895892749 | 0,000119014 | 0,04419246 | Upregulated |
| FADS3 | 1,642353287 | 0,000129071 | 0,045956771 | Upregulated |
| FAM180A | 4,542932801 | 2,64666E-06 | 0,005394366 | Upregulated |
| FAT3 | 4,529528023 | 0,000169698 | 0,049645989 | Upregulated |
| FMO1 | 3,417980328 | 0,000151036 | 0,046815446 | Upregulated |
| FTH1P10 | 2,016702159 | 8,79098E-06 | 0,010608886 | Upregulated |
| GAPLINC | 3,074292778 | 3,71193E-06 | 0,006324033 | Upregulated |
| GJA1 | 3,241941593 | 8,36175E-05 | 0,03795462 | Upregulated |
| GJB2 | 2,424060575 | 9,17034E-05 | 0,039026393 | Upregulated |
| HBEGF | 2,643307577 | 4,0375E-05 | 0,028521459 | Upregulated |
| HNRNPA1P71 | 2,889236422 | 5,51895E-05 | 0,033956228 | Upregulated |
| HS3ST2 | 3,421763355 | 6,48299E-05 | 0,034139425 | Upregulated |
| ITGB5 | 2,6623336 | 6,33096E-05 | 0,033956228 | Upregulated |
| ITGB8 | 3,477252676 | 0,000111634 | 0,043692732 | Upregulated |
| KCNA1 | 4,769734154 | 1,26685E-05 | 0,012019062 | Upregulated |
| MGAT4C | 5,181779019 | 0,000174165 | 0,049943983 | Upregulated |
| MRPL49P2 | 7,618270918 | 7,49367E-05 | 0,036173179 | Upregulated |
| MSR1 | 3,547929199 | 1,16205E-05 | 0,011605643 | Upregulated |
| MT1G | 4,293008674 | 0,00013031 | 0,045956771 | Upregulated |
| MYOZ1 | 2,793235706 | 7,72905E-06 | 0,009732889 | Upregulated |
| NUPR1 | 2,509593383 | 4,16459E-06 | 0,006701051 | Upregulated |
| OSR2 | 4,123284758 | 5,03908E-05 | 0,03321378 | Upregulated |
| PDGFD | 2,880747608 | 2,43039E-05 | 0,018904403 | Upregulated |
| PGBD5 | 2,919417307 | 0,000114721 | 0,043719127 | Upregulated |
| PLAT | 3,748300177 | 5,06146E-05 | 0,03321378 | Upregulated |
| PODN | 3,144042139 | 6,13837E-05 | 0,033956228 | Upregulated |
| PRR15 | 3,269829695 | 0,000142123 | 0,045956771 | Upregulated |

| | | | | |
|---|---|---|---|---|
| RF00100 | 4,434327827 | 1,65629E-05 | 0,014536673 | Upregulated |
| RHOBTB1 | 1,433776423 | 8,76176E-05 | 0,038654512 | Upregulated |
| RNY1 | 3,884175718 | 0,000141803 | 0,045956771 | Upregulated |
| RNY3 | 6,404174713 | 5,40981E-06 | 0,007461162 | Upregulated |
| ROR2 | 2,627655634 | 0,000116803 | 0,04393461 | Upregulated |
| SDS | 2,661416484 | 0,000132093 | 0,045956771 | Upregulated |
| SERHL | 1,85732508 | 9,29745E-05 | 0,039026393 | Upregulated |
| SERHL2 | 2,377027822 | 9,96404E-06 | 0,010963927 | Upregulated |
| SERTAD4 | 3,57004094 | 1,62219E-05 | 0,014536673 | Upregulated |
| SERTAD4-AS1 | 3,466776218 | 0,000114469 | 0,043719127 | Upregulated |
| TRDV1 | 4,064634548 | 2,11757E-05 | 0,017523221 | Upregulated |
| VSIG4 | 4,548835082 | 5,20549E-06 | 0,007461162 | Upregulated |
| AC005599 | -1,776515143 | 0,000161803 | 0,048029072 | Downregulated |
| AC037471 | -4,134296712 | 0,000141218 | 0,045956771 | Downregulated |
| AC068295 | -4,751532921 | 0,000162512 | 0,048029072 | Downregulated |
| AC116565 | -7,582774202 | 7,78847E-05 | 0,036979893 | Downregulated |
| ADCYAP1 | -5,238684612 | 0,000133386 | 0,045956771 | Downregulated |
| AL392086 | -3,599999379 | 0,000148848 | 0,046815446 | Downregulated |
| AP002518 | -3,515101089 | 2,54556E-05 | 0,018904403 | Downregulated |
| AP002755 | -3,889544194 | 1,28644E-05 | 0,012019062 | Downregulated |
| C1QTNF4 | -5,238325726 | 2,40053E-08 | 0,000231755 | Downregulated |
| CCK | -9,30047656 | 7,42399E-05 | 0,036173179 | Downregulated |
| CLEC4C | -2,882446958 | 0,000130494 | 0,045956771 | Downregulated |
| CRB1 | -5,581094593 | 6,51333E-07 | 0,002096061 | Downregulated |
| DDC | -4,168550961 | 2,37657E-07 | 0,000860406 | Downregulated |
| EPHA10 | -4,108796052 | 0,00011091 | 0,043692732 | Downregulated |
| ERBB4 | -4,021773348 | 6,23455E-05 | 0,033956228 | Downregulated |
| FAM95C | -4,560869067 | 2,25751E-06 | 0,005394366 | Downregulated |
| GTSF1L | -3,783633824 | 9,22909E-05 | 0,039026393 | Downregulated |
| HMX3 | -4,178957113 | 0,000103054 | 0,041454713 | Downregulated |
| IAPP | -5,503649278 | 1,13971E-05 | 0,011605643 | Downregulated |
| KCNK9 | -3,55217144 | 8,80847E-05 | 0,038654512 | Downregulated |
| KIR3DL2 | -4,243636175 | 0,000155221 | 0,046829861 | Downregulated |
| KLHL33 | -3,903933214 | 3,81839E-05 | 0,027648002 | Downregulated |
| KSR2 | -7,443100785 | 1,34996E-12 | 1,95495E-08 | Downregulated |
| LINC002481 | -2,822022921 | 6,82868E-06 | 0,008989956 | Downregulated |
| LINC00511 | -4,041609588 | 7,38706E-05 | 0,036173179 | Downregulated |
| LINC00707 | -3,789939418 | 8,38689E-05 | 0,03795462 | Downregulated |
| LINC01833 | -5,441164916 | 7,14663E-05 | 0,036173179 | Downregulated |
| LINC01958 | -5,790398385 | 5,86919E-05 | 0,033956228 | Downregulated |
| LPAL2 | -3,180995463 | 1,83424E-05 | 0,015625013 | Downregulated |
| MIR4422HG | -3,237597673 | 5,16045E-05 | 0,03321378 | Downregulated |
| NTRK1 | -2,974408582 | 6,1334E-05 | 0,033956228 | Downregulated |
| PHF19 | -1,785028542 | 8,05991E-05 | 0,037651459 | Downregulated |
| PRSS2 | -5,652135019 | 3,16184E-06 | 0,005723515 | Downregulated |
| RAB44 | -4,639817109 | 4,95461E-06 | 0,007461162 | Downregulated |

| | | | | |
|---|---|---|---|---|
| RAPGEF4 | -2,909666319 | 6,95899E-05 | 0,035991666 | Downregulated |
| RPL7AP28 | -6,699575059 | 4,47475E-08 | 0,000296449 | Downregulated |
| SALL3 | -8,952752084 | 1,02208E-05 | 0,010963927 | Downregulated |
| SLC22A20P | -3,687731119 | 2,5837E-06 | 0,005394366 | Downregulated |
| SLC44A3 | -2,723747296 | 0,000138056 | 0,045956771 | Downregulated |
| SLCO1A2 | -5,734014464 | 2,79375E-06 | 0,005394366 | Downregulated |
| SLCO1B1 | -6,204005329 | 4,31563E-05 | 0,029760381 | Downregulated |
| SNX18P3 | -4,036696618 | 0,000153557 | 0,046815446 | Downregulated |
| SYT16 | -4,514877427 | 1,87668E-07 | 0,000776491 | Downregulated |
| TEX15 | -6,298776483 | 2,14607E-06 | 0,005394366 | Downregulated |
| TOGARAM2 | -1,975728764 | 0,00014866 | 0,046815446 | Downregulated |
| YBX2 | -5,410183635 | 5,11772E-08 | 0,000296449 | Downregulated |
| ZBTB16 | -3,700435053 | 9,66094E-05 | 0,039972846 | Downregulated |
| ZNF793-AS1 | -3,272492107 | 0,00010302 | 0,041454713 | Downregulated |

**Supplementary Table S6:** Clinical information about the *NOTCH1*-rearranged patients.

| Patient ID | TLBL049 | TLBL033 | TLBL050 | TLBL052 | TLBL058 | TLBL042 |
|---|---|---|---|---|---|---|
| Diagnosis | T-LBL | T-LBL | T-LBL | T-LBL | T-LBL | T-LBL |
| Fusion | *TRBJ*::*NOTCH1* | *TRBJ*::*NOTCH1* | *IKFZ2*::*NOTCH1* | *TRBJ*::*NOTCH1* | *miR142*::*NOTCH1* | *miR142*::*NOTCH1* |
| Age | 8 | 13 | 10 | 12 | 17 | 16 |
| Sex | Male | Female | Male | Male | Male | Female |
| *NOTCH1* status | WT[1] | WT[1] | WT | WT | WT | WT |
| *FBXW7* status | WT[1] | WT[1] | WT | WT | WT | WT |
| Other driving mutations | - | - | Homozygous loss CDKN2A/B | None | None | Homozygous loss CDKN2A/B |
| Stage | III | III | III | III | III | III |
| Event | t-AML | Relapse | Relapse | - | Relapse | Relapse |
| CNS status | CNS1 | CNS1 | CNS1 | CNS1 | CNS1 | CNS1 |
| Peripheral blood (% blasts)[2] | negative | negative | negative | negative | negative | negative |
| Bone marrow (% blasts)[2] | negative | negative | negative | negative | negative | negative |
| Diagnosis material | mediastinal mass | pleural fluid | pleural fluid | pleural fluid | pericardial fluid | mediastinal mass |
| LDH diagnosis | 1156 | 1138 | 1566 | 1497 | 1018 | 666 |
| LDH relapse | - | 336 | 1138 | - | - | 616 |
| Mediastinal mass | + | + | + | + | + | + |
| Pericardial fluid | - | + | - | + | + | - |
| Pleural fluid | + | + | + | + | + | + |
| Kidney involvement | - | + | - | - | - | + |
| TARC at diagnosis (pg/ml) | 10000 | 10000 | 10000 | 10000 | 8067 | 2345 |

[1] Mutational status determined by variant calling on RNA sequencing data.
[2] Based on gold standard cytomorphology.

| | | | | | | |
|---|---|---|---|---|---|---|
| TARC at first remission (pg/ml) | 152 | - | - | 57 | 62 | - |
| TARC at relapse (pg/ml) | 75 (t-AML) | 153 | 8654 | - | 1662 | 4613 |
| TARC at second remission (pg/ml) | - | 69 | 946 | - | 158 | 136 |
| Flowcytometry | mCD3 partially; cyCD3+; CD5+; CD10+; CD4+; CD8+; CD1a+; Tdt-; CD30- | CD1a partially, CD2+, mCD3-, cyCD3+, CD4+, CD5+, CD7+, CD8+, CD10+, CD19-, CD30-, CD45+, CD56+, cyTdT-, TCR-AB-, TCR-GD- | CD1a+, CD2+, mCD3-, cyCD3+, CD4+, CD5+, CD7+, CD8+, CD10+, CD30-, CD38+ CD45+, CD56-, TdT-, TCR-GD-, TCR-AB-, CD99-, CD45RA-, CD34-, TCRbF+ | CD1a+, CD2+, mCD3+, cyCD3+, CD4+, CD5+, CD7+, CD8+, CD10 partially, CD13-, CD19-, CD33-, CD34-, CD44-, CD45 zw, CD45-RA-, CD56-, cyCD79a-, CD99 weak, CD117-, CD123-, cyB-F1+, cyMPO-, cyTdT+, HLADR-, TCR-AB+, TCR-GD-, CD38+, CD30- | CD1a+, CD2+, mCD3+, cyCD3+, CD4+, CD5+, CD7+, CD8+, CD10-, CD13-, CD19-, CD33-, CD34-, CD38+, CD44-, CD45+, CD45-RA-, CD56-, cyCD79a-, CD99 weak, CD117-, CD123-, TCR-AB+, cyB-F1+, HLADR-, TCR-GD-, cyMPO-, cyTdT+ | CD45weak+, mCD3+, CyCD3+, CD10 partially+, CD1a+, CD5+, CD7+, CD2+, CD4 partially+, CD8+, CD30-, TdT partially+, TCRAB-, TCRGD-, CD99+, CD34- |
